# Assessing Covariate Balance with Small Sample Sizes

**DOI:** 10.1101/2024.04.23.24306230

**Authors:** George Hripcsak, Linying Zhang, Yong Chen, Kelly Li, Marc A. Suchard, Patrick B. Ryan, Martijn J. Schuemie

**Author notes:** **Corresponding author:** George Hripcsak, Department of Biomedical Informatics, Columbia University Irving Medical Center, 622 W. 168th Street, PH20, New York, 10032, NY, USA. **Ethics of approval statement:** The research was approved by the Columbia University Institutional Review Board as an OHDSI network study. **Data availability:** The Merative and Optum databases used in this real-world study are not available from the authors but are available for licensing by those data owners.

## Abstract

Propensity score adjustment addresses confounding by balancing covariates in subject treatment groups through matching, stratification, or weighting. Diagnostics test the success of adjustment. For example, if the standardized mean difference (SMD) for a relevant covariate exceeds a threshold like 0.1, the covariate is considered imbalanced and the study may be biased. Unfortunately, for studies with small or moderate numbers of subjects, the probability of identifying a study as biased because of chance imbalance can be grossly larger than a given nominal level like 0.05, yet that chance imbalance may not cause significant bias. In this paper, we illustrate that chance imbalance is operative in real-world settings even for moderate sample sizes of 2000. We identify a previously unrecognized challenge that as meta-analyses increase the precision of an effect estimate, the diagnostics must also undergo meta-analysis for a corresponding increase in precision. We propose an alternative diagnostic that checks whether the standardized mean difference statistically significantly exceeds the threshold. Through simulation and real-world data, we find that this diagnostic achieves a better trade-off of type 1 error rate and power than standard nominal threshold tests and not testing for sample sizes from 250 to 4000 and for 20 to 100,000 covariates. We confirm that in network studies, meta-analysis of effect estimates must be accompanied by meta-analysis of the diagnostics or else systematic confounding may overwhelm the estimated effect. Our procedure supports the review of large numbers of covariates, enabling more rigorous diagnostics.

## 1 INTRODUCTION

Observational research carries a mixed reputation, with concern about biased and contradictory studies, leading to distrust and limited use. ^1^ A major source of bias is the multiplicity of pathways that are commonly pursued in executing an observational research study, ^2^ leading to problems like inflated type 1 error rate and the potential for the researcher to steer the study to a desired outcome. One way to address this challenge is with a pre-specified protocol and strict diagnostics whose failure results in rejecting the study instead of iterating on the design. Such a framework has been developed ^3,4^ and used successfully to generate high-impact evidence. ^5-7^ The framework requires diagnostics ^8^ that are discriminating in the sense of identifying studies with substantial potential bias and not identifying studies with inconsequential bias. The purpose of this paper is to improve the discrimination of a diagnostic related to covariate imbalance. We frame this paper in terms of accepting and rejecting studies according to the diagnostic, which simplifies the explanation, but we recognize that diagnostics provide several pathways with varying strictness. The diagnostics can be used to blind the results without limiting iteration on the design. This discourages researchers from iterating to a desired result (because the imbalanced study results are hidden), but it still suffers from multiplicity. The researcher may choose to use the diagnostic purely as a descriptive statistic. Despite the path taken, our goal is to create a more useful diagnostic to discriminate biased studies.

Observational research carries the risk of producing a biased estimate due to confounding. Propensity score adjustment, invented 40 years ago, ^9^ is a commonly used solution for adjusting measured confounders. It is a balancing score in the sense that matching subject treatment groups based on the score tends to balance all of the covariates used to estimate the score. Applying the score to a causal analysis can be done in several ways—matching, stratification, inverse probability treatment weighting, etc.—while the goal is the same: mitigate the impact of the confounders on the causal estimate. To ensure that the adjustment has been effective, covariate imbalance is often used as a diagnostic, ^10^ with standardized mean difference (SMD) of suspected confounders across the treatment groups under comparison being a common metric. ^11^ A high SMD reflects imbalance among the treatment groups and potentially ineffective adjustment for that potential confounder. SMD identifies only the treatment link of the covariate and not the link to the outcome, so we refer to the covariates as potential confounders.

The diagnostic is typically used as a descriptive statistic, and imbalance is addressed through assertion of non-confounding, inclusion in the outcome model, switching to doubly robust methods, or modifications to the propensity model. Unfortunately, this produces the kind of multiplicity that Hoffmann et al. ^2^ cautioned about. We believe that with a sufficiently discriminating diagnostic (measured in terms of type 1 error and power), adherence to strict diagnostic criteria with rejection of studies can improve reliability and credibility.

It is known that SMD can falsely declare studies as biased when sample size is too small or too large. ^11^ With small sample sizes, chance imbalance, i.e., the probability of asserting imbalance by using a cutoff for SMD when no underlying balance exists, can cause large deviations of the standardized mean difference from zero. That is, observed imbalance may be due to systematic differences in treatment groups (that may reflect confounding) or may be due to chance sampling variations between treatment groups, and the latter increases as sample size decreases. To account for chance imbalance, Austin ^11^ suggests measuring the empirical distribution of the standardized mean difference, but this is rarely done in practice. Additional improvements such as comparing moments beyond the mean have been suggested ^11^ but are rarely used. We note that even chance imbalance can cause bias if the covariate is linked to the outcome, but we also note that most chance imbalance is accommodated by the larger confidence intervals produced with smaller sample sizes. That is, a small amount of confounding or chance bias will produce little effect on a study with a wide confidence interval.

With large sample sizes, small degrees of systematic imbalance can be detected even though imbalance at that level is unlikely to cause an appreciable change in the effect estimate. ^11^ Researchers often pick a threshold to which a nominal estimate of the standardized mean difference can be compared; 0.1 is chosen most often ^11-23^ but 0.25 has also been used. ^19,20^

The problem of chance imbalance grows as sample size decreases, either due to a small observational data source or an uncommon treatment (i.e., imbalance between two treatment groups). The problem of chance imbalance is also more pronounced with increasing number of covariates. Traditional manual selection of confounders typically investigates 5 to a few dozen covariates. High-dimensional propensity score adjustment ^24^ leads to hundreds of covariates. Large-scale propensity score adjustment ^25,26^ leads to tens of thousands of covariates. Using a formula we derived in Section 2, we estimate that with a sample size of 250, it takes only 5 covariates to reject 90% of studies by chance, and with a sample size of 1000, it takes as few as 20 covariates. These numbers are well within the number of covariates often adjusted for in traditional observational research. As demonstrated in this paper, most of these studies need not be marked as biased. We therefore require a diagnostic that can identify imbalance large enough to produce a noticeable change in the effect estimate, which we quantify in terms of type 1 error rate and power.

A further challenge arises with the growth of networks of observational databases and federated analyses across those databases. ^27-30^ Combining the effect estimates from the databases using meta-analysis can result in more precise estimates. The increasing precision of the effect estimates should be matched with increased precision of diagnostics to ensure the reproducibility of the research findings. That is, if there is systematic imbalance across the databases, then the level of imbalance will have a proportionately larger effect compared to the increased precision of the effect estimate. Therefore, any diagnostic for a meta-analytic effect estimate must go beyond independent database-level diagnostics and operate at the network level.

The covariates that are tested for imbalance are usually the covariates adjusted for. Most commonly, researchers select a small number of covariates, say 4 to 20, that are suspected to be potential confounders and adjust for them. The process is unreliable, as illustrated by a sample of hypertension studies, ^31-35^ each of which claims to have adequately addressed confounding but for which there is only moderate overlap of the confounders across studies. An alternative, which has empirical backing, is to adjust for all observed covariates—potentially tens of thousands—that are not eliminated as potential mediators, colliders, or instruments. ^25,26^ In comparisons, adjusting for all covariates has been shown to outperform manual confounder selection ^26,36,37^ and empirical confounder selection. ^25^

Even if the researcher chooses to adjust for a small number of potential confounders, the ability to diagnose imbalance on large numbers of covariates is still useful. Whether a covariate is adjusted for or not, significant imbalance carries information and should be addressed. If imbalanced covariates are not plausible instruments, then they should be adjusted for or the study should be discarded.

The rest of this paper is organized as follows. In Section 2, we propose a simple yet effective heuristic based on a hypothesis testing framework to address excessive false rejection beyond a given nominal level (e.g., 0.05) due to chance imbalance. Specifically, we develop a statistical test for exceeding the 0.1 threshold and with Bonferroni correction where appropriate. We emphasize that our goal is not to make assertions about the underlying balance distribution, but to create a heuristic that reflects the potential impact of imbalance in the current sample. Our procedure is applicable to safeguard against excessive false rejection at either a single database level or the network level. The approach is easy to understand and easy to implement. In Section 3, we evaluate the empirical performance of the proposed methods using simulation study. In Section 4, we study the operating characteristics of this diagnostic procedure on real-world data, and we report on the implications for single database studies and for meta-analyses. We conclude this paper with a discussion of the practical implications of our methods to clinical evidence generation using observational data in Section 5.

## 2 ALGORITHM

### 2.1 Standardized mean difference

For each subject *i*, let *t*_*i*_ be the binary treatment status, let *y*_*i*_ be the binary outcome status, and let there be *J* binary non-treatment covariates *x*_*i,j*_. To diagnose the potential for confounding, we seek to quantify the imbalance between the two treatment groups for each of the covariates, where high imbalance on a covariate that is a confounder can lead to biased outcome estimates. To quantify covariate imbalance, we use the standardized mean difference (SMD), *d*_*j*_, for covariates *x*_*i,j*_, defining it as follows (with *n*_*t*_ being the number in each treatment group and *s*_*t,j*_ being the sum of the covariates in each treatment group):

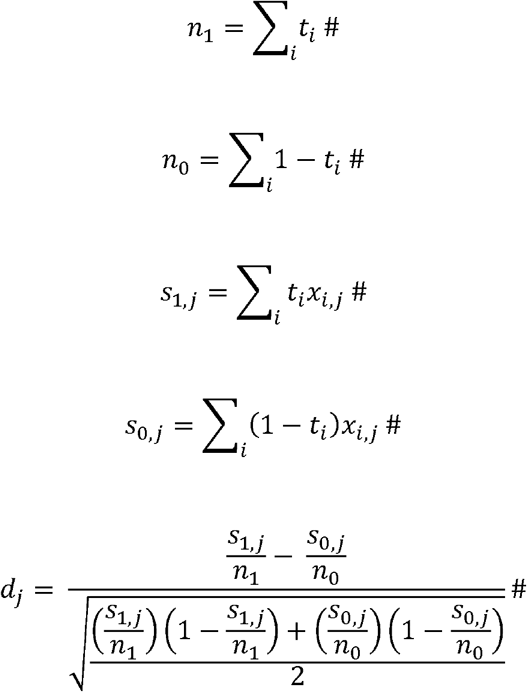

The estimated asymptotic variance of *d*_*j*_ can be calculated as follows ^38^:

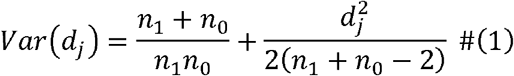

### 2.2 Statistical properties of the current diagnostic

The most common imbalance diagnostic ^11-23^ is to check whether *any covaria*te has |*d*_*j*_| > 0.1, signifying substantial imbalance that may lead to a notable shift in the effect estimate. In this paper, we refer to this as the nominal test for imbalance. The probability of falsely rejecting a study that has no actual confounding or instruments can be calculated.

To illustrate the impact of multiplicity of covariates on the false positive rejection, we consider J independent binary covariates and a diagnostic threshold of 0.1 on |*d*_j_|. We can calculate the probability of false rejection under the null hypothesis that none of the covariates is imbalanced (other than by chance). This approach is similar to that of Han and Sidell (e.g., see their Equation 3), ^39^ and is covered further in the Discussion. Specifically, the probability of incorrectly rejecting one balanced covariate is as follows:

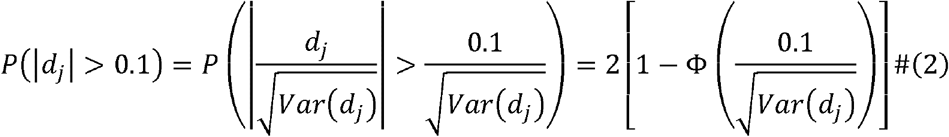

When there are totally independent balanced covariates, the probability of falsely rejecting at least one covariate is as follows:

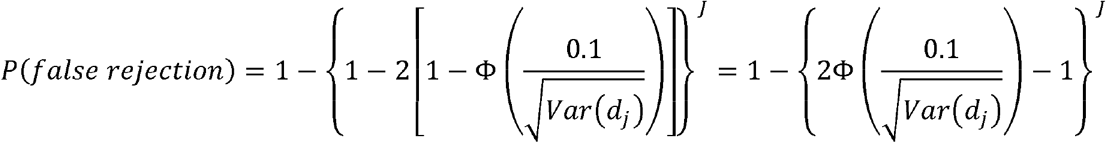

To provide a simpler expression of the above results, we consider a scenario where the covariates are all binary with prevalence 0.5, and the treatment and control of equal size *n*_1_=*n*_0_=*n*, Equation (1) can be simplified as:

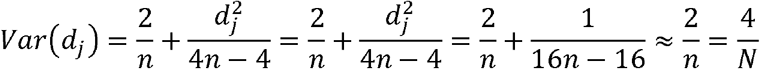

where *N*=2*n*is the total sample size.

Thus, the probability of falsely rejecting at least one covariate can be further simplified to

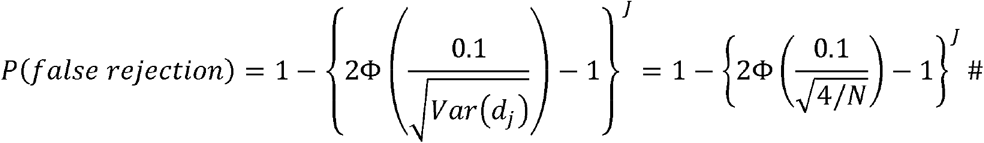

where Φ is the cumulative distribution function of the standard normal distribution and N is the total sample size, *n*_1_+*n*_0_. With N=250, it takes only 5 covariates to reject 90% of studies by chance, and with N=1000, it takes 20 covariates.

### 2.3 Our modified diagnostic

Motivated by the above calculation and the need to account for multiplicity, we propose to use Bonferroni correction for *J* covariates, which can be correlated. Specifically, we check whether *any* standardized mean difference exceeds the threshold of 0.1 beyond what would be expected by chance, in which case the study is rejected as being imbalanced. We propose the following test statistic:

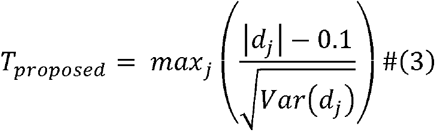

which is compared with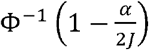. We will reject the null hypothesis that none of the covariates is imbalanced enough to cause substantial bias, if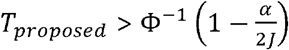 at the nominal level of α, such as 0.05. We emphasize that the null hypothesis here differs from that of Equation (2). In Equation (2), the null hypothesis is that *d*_*j*_ is 0 in order to estimate the probability of declaring a study imbalanced beyond our threshold when it is not imbalanced at all. For our new test statistic, Equation (3), we are not looking to detect all imbalance, only imbalance that is substantial as defined by the 0.1 threshold. Therefore, the null hypothesis is that *d*_*j*_ is within –0.1 to 0.1, implying that any imbalance in the study covariates is insufficient to cause substantial bias according to the widely adopted heuristic threshold of 0.1. ^11^

There are several properties that would be desirable for such a test as described by the following remarks.

Remark 1: One property is that with increasing sample size, it approximates current practice of testing for nominally exceeding a threshold. Making the simplifying assumption that *n*_1_=*n*_0_ and again defining *N* as *n*_1_+*n*_0_, Var(*d*_*j*_) varies by approximately 1/*N*. Therefore, as sample size approaches infinity, the rule approaches a nominal test of whether any *d*_*j*_ exceeds 0.1. We recognize that such a threshold is just a heuristic, and even the most cited imbalance paper by Austin ^11^ cites previous authors that picked a heuristic. ^12^ Nevertheless, over two decades, the research community has largely employed two thresholds, 0.1 and 0.25, and our test mimics the first with large sample sizes.

Remark 2: A second desirable property of the rule is that it accommodates greater imbalance (and potentially worse confounding) as the effect estimate grows less precise. That is, as the confidence interval of the effect estimate grows in size, it will take a larger amount of confounding to produce a change in the effect estimate that is significantly larger than the confidence interval. We recognize that this statement is most easy to interpret in the setting of collapsible analyses like linear regression and risk ratios, but to first order, it remains true of other designs. For example, one would expect that confounding that can shift a risk ratio by 0.2 to be very important to an effect estimate whose confidence interval is 0.1 wide but to be inconsequential to one whose confidence interval is 4.0 wide. Noting that in general and to first order, sampling variance for effect estimates will affect standard errors and therefore confidence intervals by a factor of 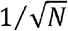, we seek a test with a similar dependence on sample size. For our test, as N falls, the new threshold is set by the standard error of *d*_*j*_, *α*, and *J*. For g iv en *α* and *J*, the rise of the effective threshold above 0.1 is approximately proportional to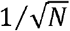. For example, with one-fourth the sample size, the effective threshold will double its level above 0.1. (Each covariate sees a slightly different threshold due to the *d*_*j*_ term in *Var* (*s m d*_*j*_), which is about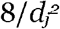 times smaller than the first term.) Therefore, our test also follows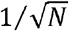, other than the shift of0.1 due to the threshold.

Remark 3: A third property is to accommodate multiple covariates. As the number of covariates rises, the probability of chance imbalance rises. We employ a Bonferroni correction, which requires the fewest assumptions (for testing whether imbalance may be by chance) but is the most conservative. It effectively raises the threshold. Use of the Bonferroni correction is discussed further below and tested in a sensitivity analysis. We assert that if our test with a Bonferroni correction can outperform current methods, then that demonstrates the benefit of a new test, and it leaves open the possibility of future work with a less conservative correction ^40^ that exploits inter-covariate dependencies.

Remark 4: A fourth property is the understandability and ease of implementation of the test. Testing for statistically significant departure from a threshold is a straightforward concept to understand, is easy to implement, and provides a framework for tailoring the test for different study designs. There is a natural way to accommodate different sample sizes, different numbers of covariates, and different study designs, such as a meta-analysis of effect estimates across independent databases.

In practice, because the threshold for imbalance is just a heuristic that depends on the study data set properties, there are endless possible tests that can be imagined. Ultimately, an empirical study of the test’s operating characteristics is needed to explore its appropriateness to the task, and that is the purpose of this paper.

### 2.4 Imbalance in network studies

By “network study,” we mean a study carried out on a set of databases where similar analyses are done separately on each database and then the results are aggregated using direct meta-analysis. This two-level process is necessary where data pooling is not possible due to privacy or logistical concerns. The question we address here is how to apply a diagnostic for covariate imbalance on a network study.

Our primary motivation is as follows. The practice of testing for imbalance using a threshold on SMD ^11^ is a heuristic that has selected thresholds like 0.1 for which imbalance is small enough that any associated confounding would have an insignificant impact on the effect estimate ^11^; for example, the confidence interval of the effect estimate may be much larger than any shift in the effect estimates due to confounding. A meta-analysis may improve the precision of an effect estimate such that even a smaller amount of confounding might have significant impact on the effect estimate. If confounding is shared among the databases such that any shift in effect estimate at the database level is preserved after the meta-analysis, then as the confidence interval of the effect estimate shrinks, any shift due to confounding will be relatively larger and potentially become impactful. We therefore seek a diagnostic that accounts for the increased precision of the effect estimate for covariates that appear to share confounding. While we cannot know the confounding structure, we have measured the imbalance. Thus, for network studies, we not only aggregate the information about the effect estimate; we also aggregate information about the covariate imbalance.

For the network-level diagnostic, we test whether any of the meta-analytic SMD estimates exceed a threshold like 0.1 beyond that expected by chance. We consider the assumptions of meta-analysis as follows. A diagnostic for imbalance on a network study is only relevant if a researcher first determines that a meta-analysis is relevant and appropriate for the primary effect estimate. Therefore, we assume that study-level requirements for meta-analysis have been met; for example, we assume that databases are independent. In addition, meta-analysis requires comparability among the effect sizes, which in this case are SMDs. This first requires that covariates be coordinated among the databases. For small numbers of covariates, the covariates can be mapped among databases manually, and for large numbers of covariates, it is best if the databases share a common data model such that the covariates are named identically across databases. The use of SMD, because it is standardized, should create comparable effect sizes across databases. We do not know if confounding and therefore imbalance are shared among databases, so the SMD estimates may not be homogeneous.

We therefore employ a random effects meta-analysis, and we incorporate both sampling variance and inter-database variance in our test for exceeding the imbalance threshold. Specifically, we use the following model:

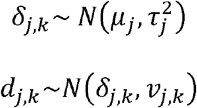

where *μ*_*j*_ is the aggregate imbalance for covariate *j* across all the network databases,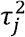is heterogeneity of that imbalance among databases, *δ*_*j,k*_ is the underlying imbalance for covariate *j* in database *k, v*_*j,k*_ is the sampling variance for estimating the SMD for covariate j in database *k*, and *d*_*j,k*_ is the estimated SMD for the *j*’th covariate for the *k*’th database. We supply R function rma with our estimated SMDs and their estimated variances for each database, and return the estimated aggregate imbalance,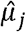 (rma’s beta), and the variance of that estimate,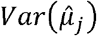(the square of rma’s se), which includes both the sampling variance (*v*_*j,k*_) and inter-database variance (i.e., heterogeneity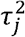). We test for imbalance using a test analogous to Equation (3):

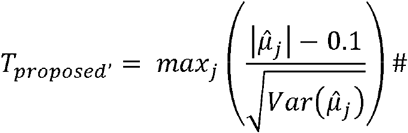

which is compared with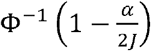for *J* total covariates. We will reject the null hypothesis that none of the covariates is imbalanced enough to cause substantial bias, if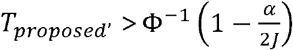at the nominal level of α, such as 0.05.

Thus,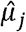, our meta-analytic estimate of the SMD for covariate *j*, represents the information we have about the imbalance of that covariate pooled across databases. Covariates with similar imbalance across databases will have less inter-database variance so that the variance,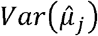, will be smaller, requiring a lower nominal level of imbalance,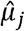, to satisfy *T*_*proposed*’_ and trigger rejection. Conversely, covariates with disparate imbalance will have more inter-database variance, higher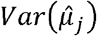, requiring a higher nominal level of imbalance,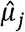, to trigger rejection.

### 2.5 Comparison of diagnostics at database level and at network level

Our diagnostic is applicable to single-database studies and to studies on networks of databases. In this paper, we compare three diagnostics for single databases: we use the shorthand of “all” for accepting all studies (i.e., no diagnostic), “**nominal**” for checking for any *d*_*j*_≥0.1 (i.e., current practice when a diagnostic is used), and “**signif**” for checking for any *d*_*j*_ being statistically significantly greater than 0.1 after Bonferroni correction (i.e., our proposed diagnostic). Below we outline the three diagnostics in the Algorithm 1.

#### Algorithm 1 Diagnostics at database-level

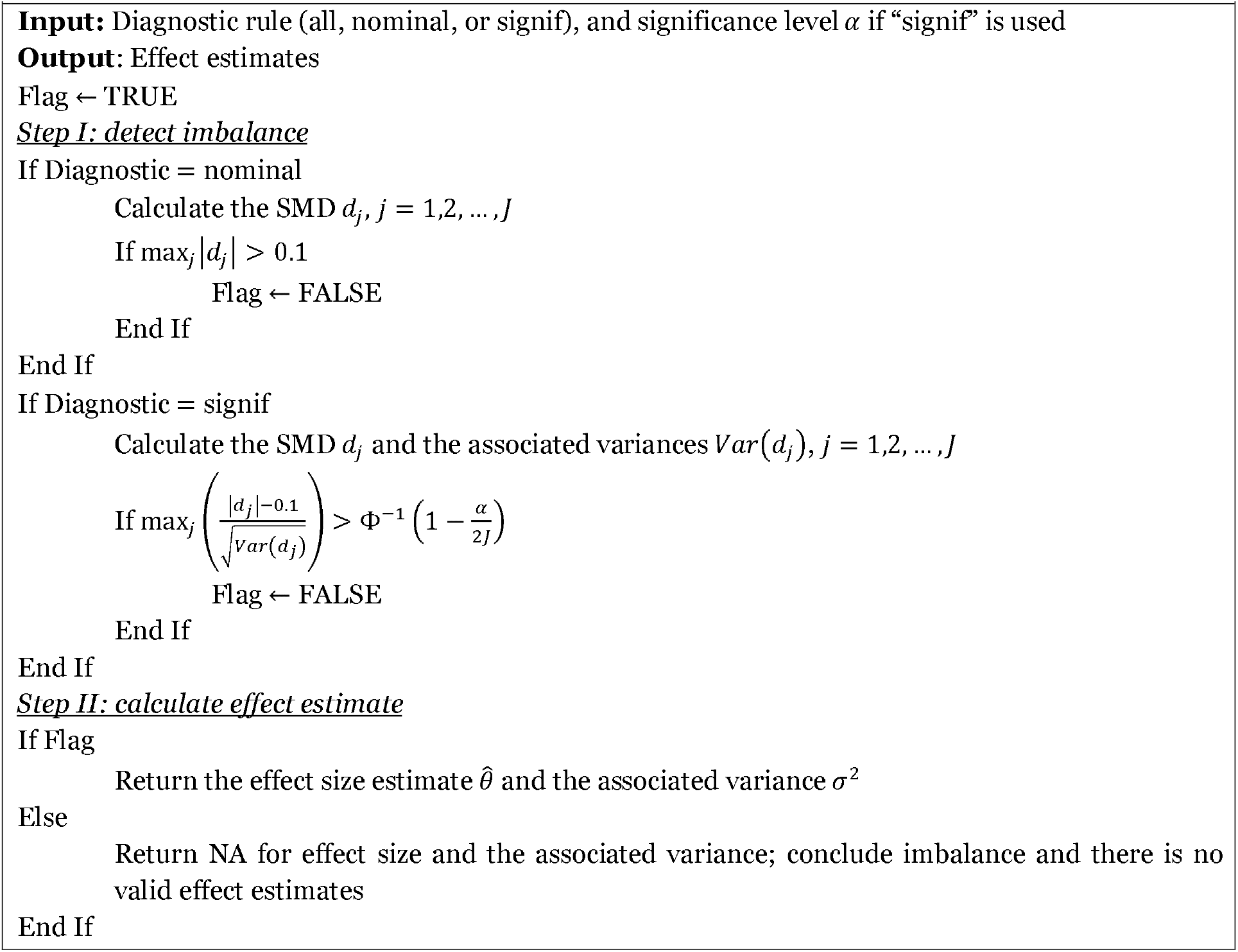

For network studies, we have two opportunities to apply our diagnostic: at the single database level before the studies are aggregated and at the network level after a meta-analysis. We employ a random effects meta-analysis using the R function rma to determine the overall standardized mean difference for each covariate. We define “overall” standardized mean difference of a covariate as the meta-analytic estimate of the SMD for that covariate.

In this paper, the three diagnostics—all, nominal, and signif—are applied at two levels, database and network, resulting in nine total rules (Table 1). With a format of “<Network diagnostic>On<Database diagnostic>,” each rule applies the database diagnostic to individual databases, rejects those databases that fail the diagnostic, runs the meta-analysis only the remaining databases, and applies the network diagnostic to the meta-analytic estimates of the covariate SMDs (the “overall” SMDs).

**Table 1.**
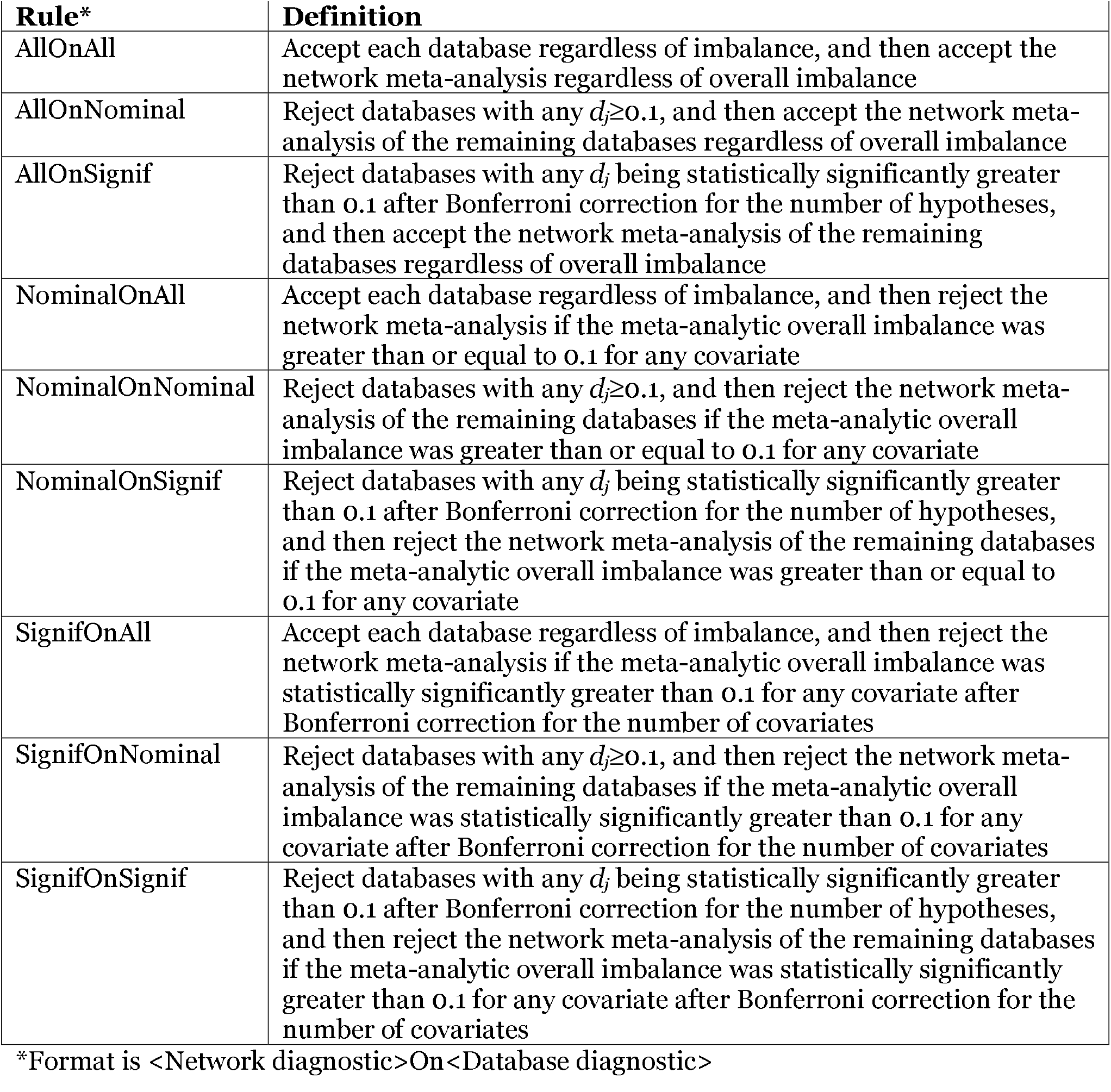
Nine diagnostic rules for network studies

Of note, rule AllOnAll ignores imbalance and accepts all studies regardless of the detection of a potential for bias. Rules NominalOnAll and SignifOnAll ignore database-level imbalance and rely solely on a network-level detection. Rules AllOnNominal and AllOnSignif ignore any network-level imbalance but exploit database-level detection. In general, database-level imbalance detection is expected to be severely affected by sample size, either missing imbalance or declaring false-positive imbalance on small databases. Network-level imbalance is expected to be more likely to detect imbalance that is shared among databases but may miss an aberrant database. Below we outline the three diagnostics in the Algorithm 2.

#### Algorithm 2

Diagnostics at network-level

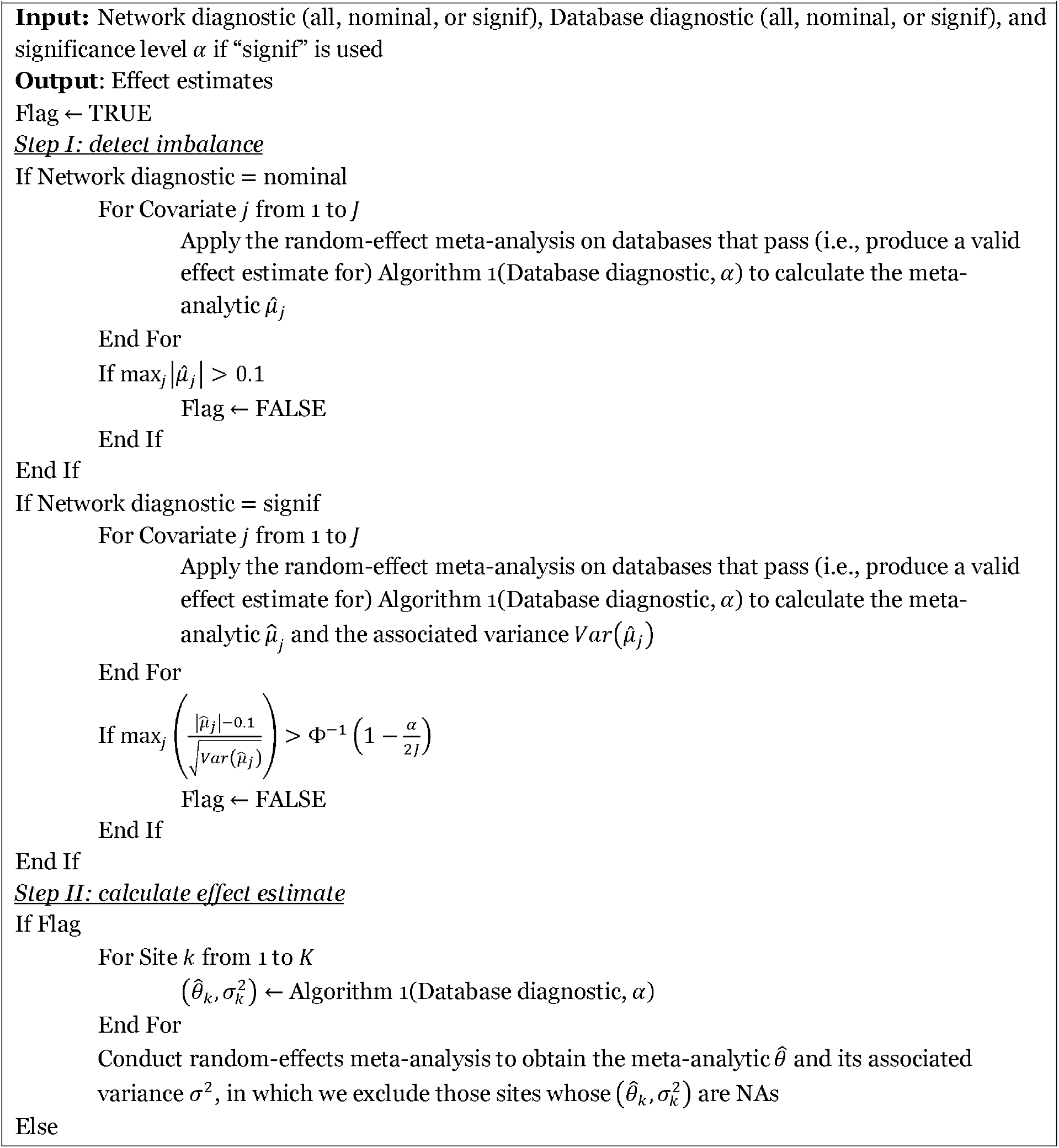
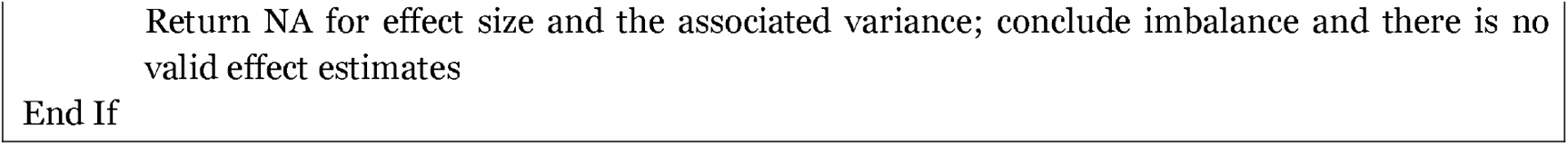

### 2.6 Performance metrics

We assess the operating characteristics of our diagnostic using type 1 error and power. Given some main hypothesis of interest and its corresponding null hypothesis, type 1 error is the probability of rejecting the null hypothesis when it is true, and power is the probability of rejecting the null hypothesis when it is false. For a test of statistical significance, our diagnostic results in three states: accept the null hypothesis, reject the null hypothesis, and reject the study as imbalanced. To best compare different diagnostics and different study scenarios, for the primary analysis, we collapse these into two states. A rejected study is treated as if it carries no information, as if the confidence intervals were infinite and thus the null hypothesis is not rejected. Therefore, we use the following estimations of type 1 error and power. Define “*positive*” as a study in which the null hypothesis is rejected, “*negative*” as a study in which the null hypothesis is not rejected, and “*invalid*” as a study that has been rejected by the imbalance diagnostic. When the null hypothesis is true, we get:

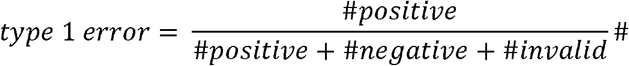

Similarly, when the null hypothesis is false, we get:

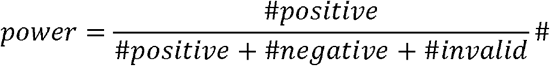

As a secondary analysis, we keep the three states separate. Here we seek to have a low proportion of positives when the null hypothesis is true, a low proportion of invalid (rejected) studies when confounding is low, and a high proportion of positives when the null hypothesis is false and confounding is low. In the discussion below, in the primary analysis, we refer to the positive rate when the null hypothesis is true as “type 1 error” whereas in the 3-state analysis we refer to it as the false positive rate even though they are numerically identical to keep clear which metric we are referring to. We do not set a target degree of confounding that should be detected by the diagnostic but instead let the false positive rate determine when too much confounding is being allowed.

## 3 SIMULATION

### 3.1 Simulation design

Our goal for the simulation was to create a data set that would mimic the balance characteristics of a data set whose treatments groups had been completely or imperfectly adjusted for confounding, for example using 1-to-1 matching based on a propensity score. We used a binary treatment and a binary outcome, with a set of binary covariates that could include a confounder or other correlations that were not sufficiently balanced by the adjustment procedure.

For the base case, we varied database sample size, effect size, and degree of confounding, holding outcome prevalence, covariate prevalence, and aggregate sample size constant and using homogeneous confounding across databases. We used an aggregate sample size of 20,000 subjects, which were apportioned among databases with 4000, 2000, 1000, 500, and 250 subjects, producing 5, 10, 20, 40, and 80 databases, respectively. Each experiment was carried out 200 times to estimate error rates. Each subject, *i*, was defined as follows:

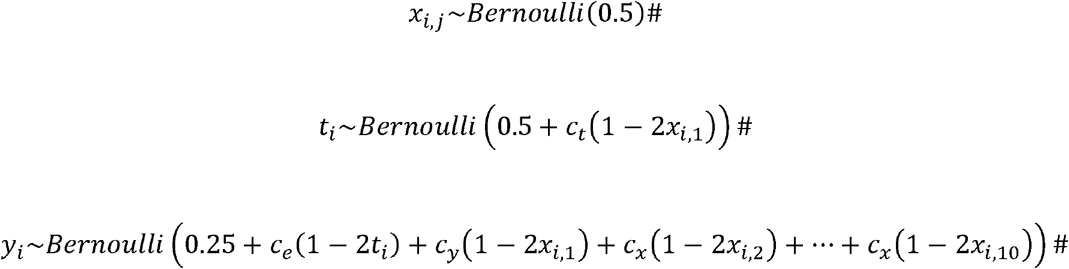

where the *x*_*i,j*_ were the non-treatment covariates for subject *i* and covariate *j, t*_*i*_ was the treatment, and *y*_*i*_ was the outcome. Index *j* varied from 1 to 1000 with *x*_*i*,1_ being a potential confounder, with *x*_*i*,2_ … *x*_*i*_,_10_ being causally linked to the outcome but not the treatment, and with *x*_*i*,11_ … *x*_*i*,1000_ not being linked to treatment or outcome. Constant *c*_*e*_ determined the treatment effect, *c*_*t*_ was the link from the confounder *x*_*i*,1_ to the treatment *t*_*i*_, *c*_*y*_ was the link from the confounder x_i,1_ to the outcome *y*_*i*_ and *c*_*x*_ was the link from covariates *x*_*i*,2_ to *x*_*i*,10_ to the outcome *y*_*i*_. We used 1000 covariates to simulate the probability of detecting imbalance among covariates by chance in an analysis with large-scale covariate adjustment such as is used in large-scale propensity score adjustment ^25,26^ or high-dimensional propensity score adjustment. ^24^ For the base case, *c*_*e*_ varied from 0 to 0.1, *c*_*t*_ varied from 0 to 0.3, *c*_*y*_ was held at 0.1, and *c*_*x*_ was held at 0.1. The parameters were chosen such that they best illustrated the range of performance, for example from an effect that was undetectable to an effect that was detectable by all studied approaches, and they were iterated upon to find weaknesses in the approaches. When *c*_*t*_ equaled zero, there was no systematic source of imbalance between the two groups and any detected imbalance was due to chance.

To estimate an effect size, we used function glm (family = binomial) in R to carry out a simple logistic regression using *t*_*i*_ to predict *y*_*i*_, ignoring *x*_*i,j*_, as one might do if one assumes a previous adjustment procedure succeeded in achieving balance. We studied our ability to detect imbalance among covariates in the treated versus untreated group, correlating those results with measured type 1 error and power based on the true effect *c*_*e*_ and the effect estimates and their variances from the model.

### 3.2 Quantifying imbalance and performance

To quantify imbalance, we used SMD as defined above. We estimated the type 1 error rate as the proportion of 200 study iterations where the effect coefficient estimate differed statistically significantly from 0 (“positive” as defined above) when *c*_*e*_=0. To test statistical significance, we compared glm’s effect coefficient to zero using glm’s estimated variance and α=0.05. We estimated the power as the proportion of the 200 iterations where the effect coefficient estimate differed statistically significantly from 0 (“positive”) when *c*_*e*_≠0.

We carried out several simulation experiments in addition to the base case. We reran the study under the conditions of low covariate prevalence (mean *x*_*ij*_ 0.1 instead of 0.5), low outcome prevalence (mean *y*_*i*_ 0.01 instead of 0.25), heterogeneous confounding (varying *c*_*t*_ from –0.3 to 0.3 for each database instead of holding it constant), and fewer databases (capped at 5 databases instead of holding the aggregate total constant).

### 3.3 Simulation results

Figure 1 shows the performance of the three types of rules at the database level, with type 1 error rate in the first column of graphs (no true effect with *c*_*e*_=0) and power in the rest of the columns (increasing effect with *c*_*e*_>0). The first row, labeled All, ignores imbalance. It has good power but increasing type 1 error with increasing confounding, reaching 1 for high confounding. The second row, labeled Nominal, uses the standard practice of rejecting studies where at least one standardized mean difference equals or exceeds 0.1. This rule rejects all studies with fewer than 4000 subjects even when confounding is zero due to chance imbalance, producing little power. The third row, labeled Signif, rejects studies where at least one standardized mean difference is statistically significantly greater than 0.1 after Bonferroni correction. Type 1 error rate is relatively controlled with an average of 0.054, with the highest rate of 0.14 at intermediate levels of confounding (*c*_*t*_=–0.06), where the confounding did not trigger a rejection but caused a false positive result. Power rises with effect size, reaching near one except where confounding is most strong, leading to rejection of those highly confounded studies.

**Figure 1.**
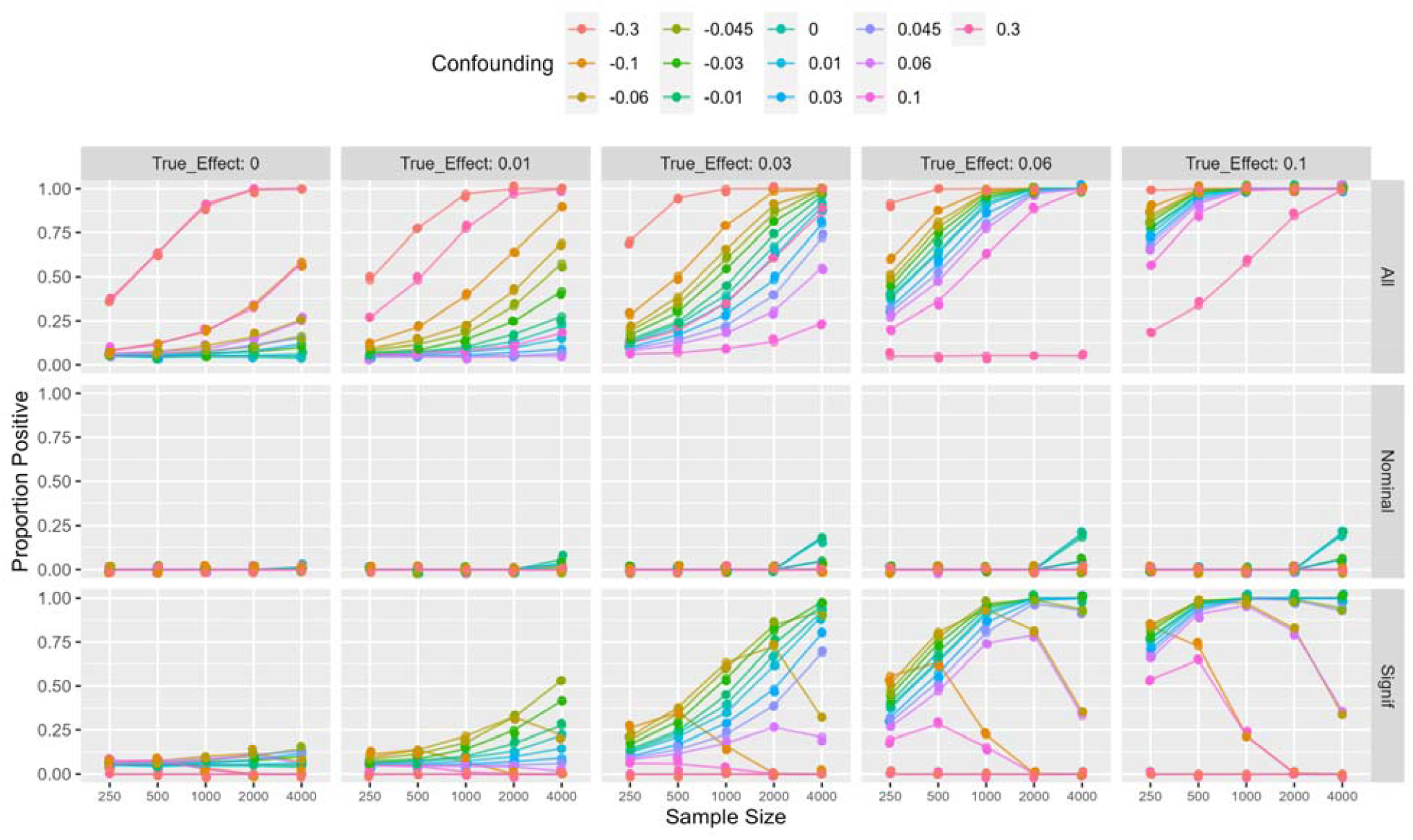
Rule performance at the database level on simulation. The proportion of study iterations that were not rejected by the rule and that had effect coefficients that were statistically significantly different from zero is plotted against database sample size. Colored lines represent different levels of confounding with *c*_*t*_ from –0.3 to 0.3, and graphs from left to right show different values for effect parameter *c*_*e*_ from 0 to 0.1. The rows represent the three types of rules applied only to a single database under study: **All** ignores imbalance, **Nominal** tests for any covariate’s standardized mean difference reaching or exceeding 0.1, and **Signif** tests for any covariate’s standardized mean difference statistically significantly reaching or exceeding 0.1. The first column, with *c*_*e*_=0, represents the type 1 error rate (i.e., higher proportion positive is undesired because it means higher type 1 error rate), and the other columns, with *c*_*e*_>0, represent the power with increasing effect size (i.e., higher proportion positive is desired because it means more power in detecting an effect). All has unacceptably high type 1 error with high confounding, Nominal has unacceptably low power, and **Signif** has moderate type 1 error rate and high power when confounding is low, and when confounding is high, more studies ought to be rejected so low power is expected. (Graph points are jittered to reveal overlapping colors but lines are drawn true.)

Figure 2 shows the performance of the nine rules for the base case of the network study. The first row, AllOnAll, illustrates that completely ignoring imbalance produces increasingly poor type 1 error rate with increasing confounding, getting to 1 for the highest confounding. The second row, AllOnNominal, illustrates that using the nominal rule of checking for standardized mean difference greater than or equal to 0.1 at the database level leads to rejecting all databases smaller than 4000 even when there is no systematic imbalance, resulting in a power of zero. For the same reason, rules NominalOnNominal and SignifOnNominal, which use the nominal test at the database level, fail with low power. The third row, rule AllOnSignif, illustrates that checking for statistically significant imbalance only at the database level leads to low type 1 error rate when there is no confounding and when there is high confounding (when it can be detected) but high type 1 error rate near 1 with intermediate confounding (*c*_*t*_=0.1 or –0.1) that is strong enough to cause a false positive result but not strong enough to trigger the rule to drop the database. The rows for rules NominalOnAll and NominalOnSign

**Figure 2.**
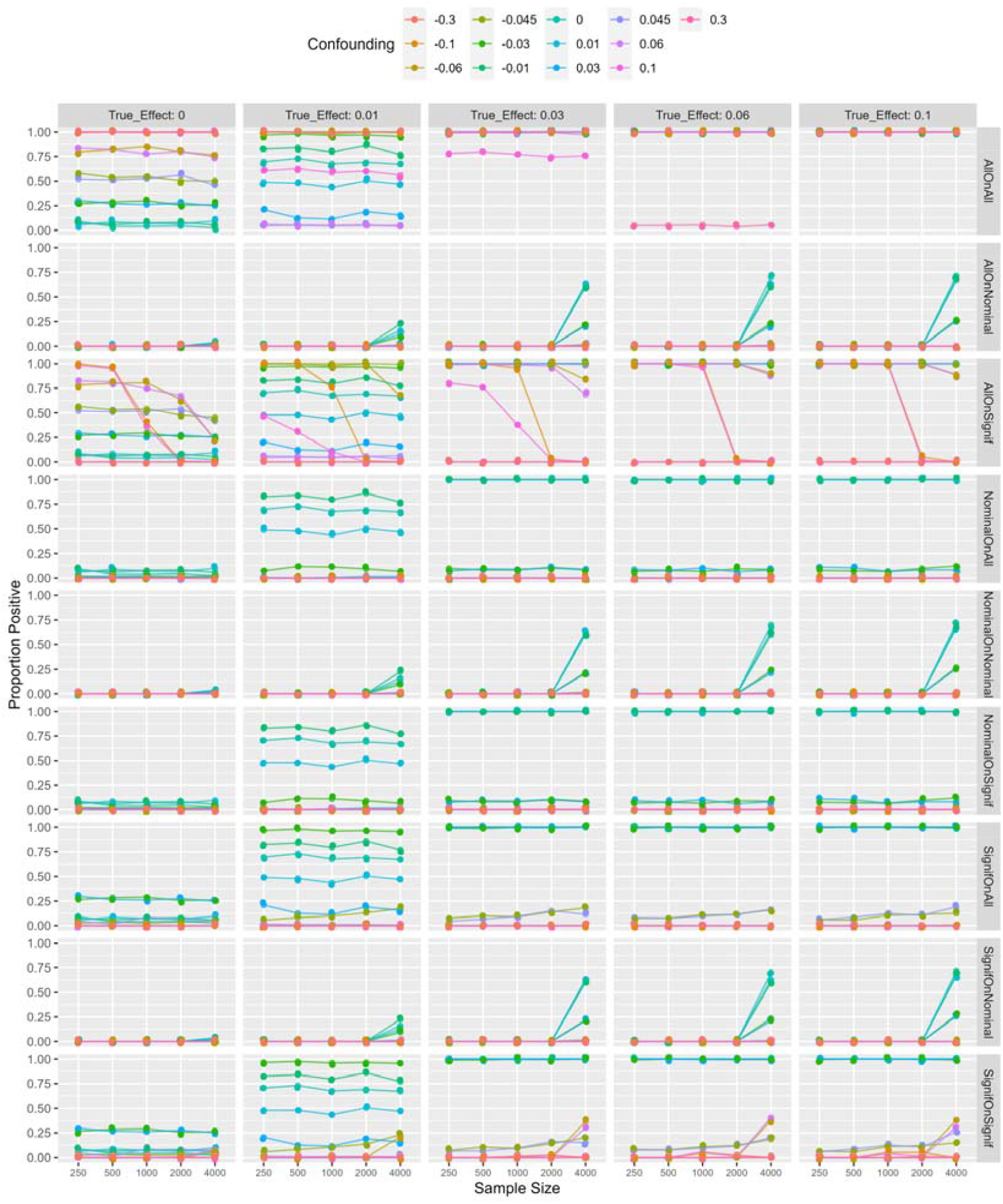
Rule performance at the network level on simulation. Graphs show the proportion of study iterations that were not rejected by the rule and that had effect coefficients that were statistically significantly different from zero plotted against database sample size. Colored lines represent different levels of confounding *c*_*t*_ from –0.3 to 0.3, and graphs from left to right show different values for effect parameter *c*_*e*_ from 0 to 0.1. The nine rows represent the nine rules listed in the Methods section. The first column, where *c*_*e*_=0, shows the type 1 error rate, and the other columns, where *c*_*e*_>0, show the power with increasing effect size. See text for an explanation of results. (Graph points are jittered to reveal overlapping colors but lines are drawn true.)

if illustrate that checking for nominally reaching the 0.1 threshold on the meta-analytic standardized mean difference estimates produces low type 1 error and high power if the confounding is low. The rows for rules SignifOnAll and SignifOnSignif illustrate that checking whether the meta-analytic standardized mean difference estimate is statistically significantly greater than or equal to 0.1 produces a generally reasonable type 1 error rate and high power for low confounding. We were able to find a combination that produced a high type 1 error rate of 0.295 when *c*_*t*_=0.03, representing an intermediate level of confounding.

Thus, four rules—NominalOnAll, NominalOnSignif, SignifOnAll, and SignifOnSignif—were viable according to the simulation, with the latter two showing some higher type 1 error. The four rules differentiated when the number of databases was limited to five (Figure S1). Using a nominal test for the meta-analytic standardized mean difference reaching the 0.1 threshold (rules NominalOnAll and NominalOnSignif) resulted in zero power for databases smaller than 1000 because chance imbalance always disqualified the study even when there was no confounding. Testing for statistical significance (rules SignifOnAll and SignifOnSignif) avoided discarding non-confounded small studies. In other words, the nominal threshold test of the meta-analytic threshold only worked when the network was large enough, but the statistical test behaved gracefully with smaller network sizes.

We also performed a number of further sensitivity analyses, shown in the Supplement. When outcome prevalence is low (Figure S2), the rules perform similarly to each other, and they all lose some power compared to the base case, where outcome prevalence is higher. When covariate prevalence is low (Figure S3), the four rules perform similarly to the base case. When confounding is heterogeneous (Figure S4), the four rules produce similar results; rules NominalOnAll and NominalOnSignif have less power, but because the study is confounded, greater power is not the goal. We tried the Signif rules without the Bonferroni correction (Figure S5); the type 1 error rate remained high at 0.260 for the c_t_=0.03 case, and it reduced power near 0 in the smallest studies.

We studied the effect of using fewer covariates. When the covariate count is 20 (Figure S6), we see similar results to the base case except that the curves are shifted to the left, with the same issues occurring across the nine rules, but with one quarter the sample size.

To provide context for the type 1 error rates at small sample sizes, we reproduced a more typical study with sample size increased to 20,000 and keeping the covariates at 20 (Figure S7). We show that type 1 error rates can get over 0.57 for all nine rules if we select confounding carefully. We also note the similarly of the results for the Nominal rules versus the Signif rules with this large sample size (Figures S7 and S16).

Figure 3 shows the 3-state results for a single database and Figure 4 shows the 3-state results for a network study, both for sample sizes of 500. Figure 3 confirms that only the Signif rule controls the number of false positive results when the true effect is zero while also producing positive results when the true effect is non-zero and there is no confounding. Power is low for smaller effect sizes, but it is similar to the All rule, which sets an upper bound on what effect can be detected (because no studies are rejected as invalid). When confounding is large enough, the Signif rule detects imbalance and rejects the study as invalid. Figure 4 shows a similar result to Figure 2, where only NominalOnAll, NominalOnSignif, SignifOnAll, and SignifOnSignif keep the false positive rate relatively low when there is no true effect and produce positive results when there is a true effect and confounding is low. The first two rules have lower false positive rates than the latter two. The results for other samples sizes are shown in the supplement and reveal similar findings.

**Figure 3.**
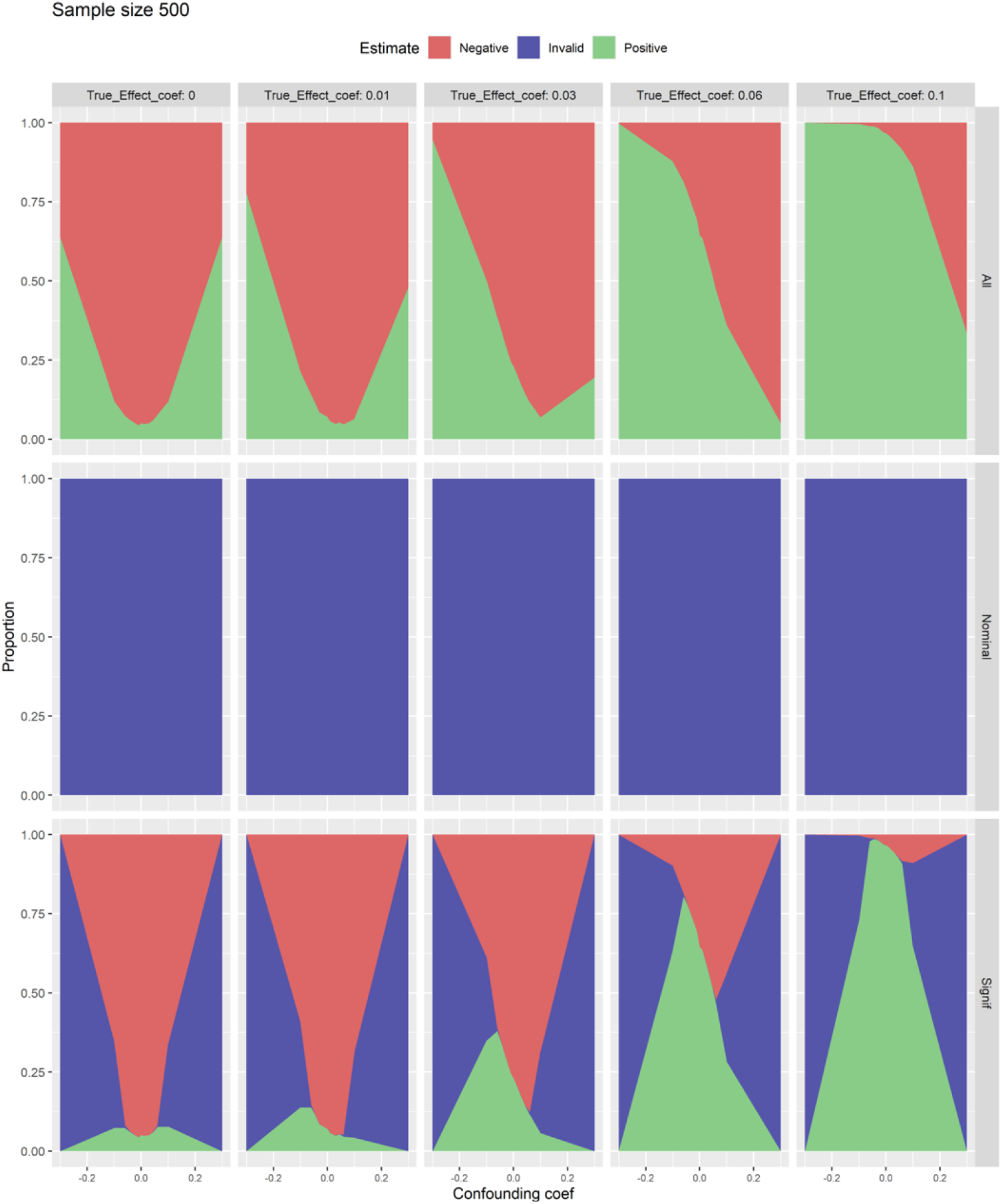
Three-state performance at the database level on simulation with sample size 500. Graphs show the proportion of studies that were positive (null hypothesis rejected), invalid (study rejected), and negative (null hypothesis not rejected) on a single-database study with sample size 500. The proportion is plotted against confounding coefficient *c*_*t*_ from –0.3 to 0.3, and graphs from left to right show different values for effect coefficient *c*_*e*_ from 0 to 0.1. The rows represent the three types of rules applied only to a single database under study: All ignores imbalance, **Nominal** tests for any covariate’s standardized mean difference reaching or exceeding 0.1, and **Signif** tests for any covariate’s standardized mean difference statistically significantly reaching or exceeding 0.1. The first column should have no more than 0.05 positive (green), and at *c*_*t*_=0, it should have no invalid (blue). The other columns should have positive as high as possible, and at *c*_*t*_=0, it should have no invalid. All has unacceptably high positive at *c*_*e*_=0; Nominal has unacceptably low positive at *c*_*e*_>0; and **Signif** has moderate positive at *c*_*e*_=0, high positive when confounding is low with *c*_*e*_>0, and more invalid when confounding is high.

**Figure 4.**
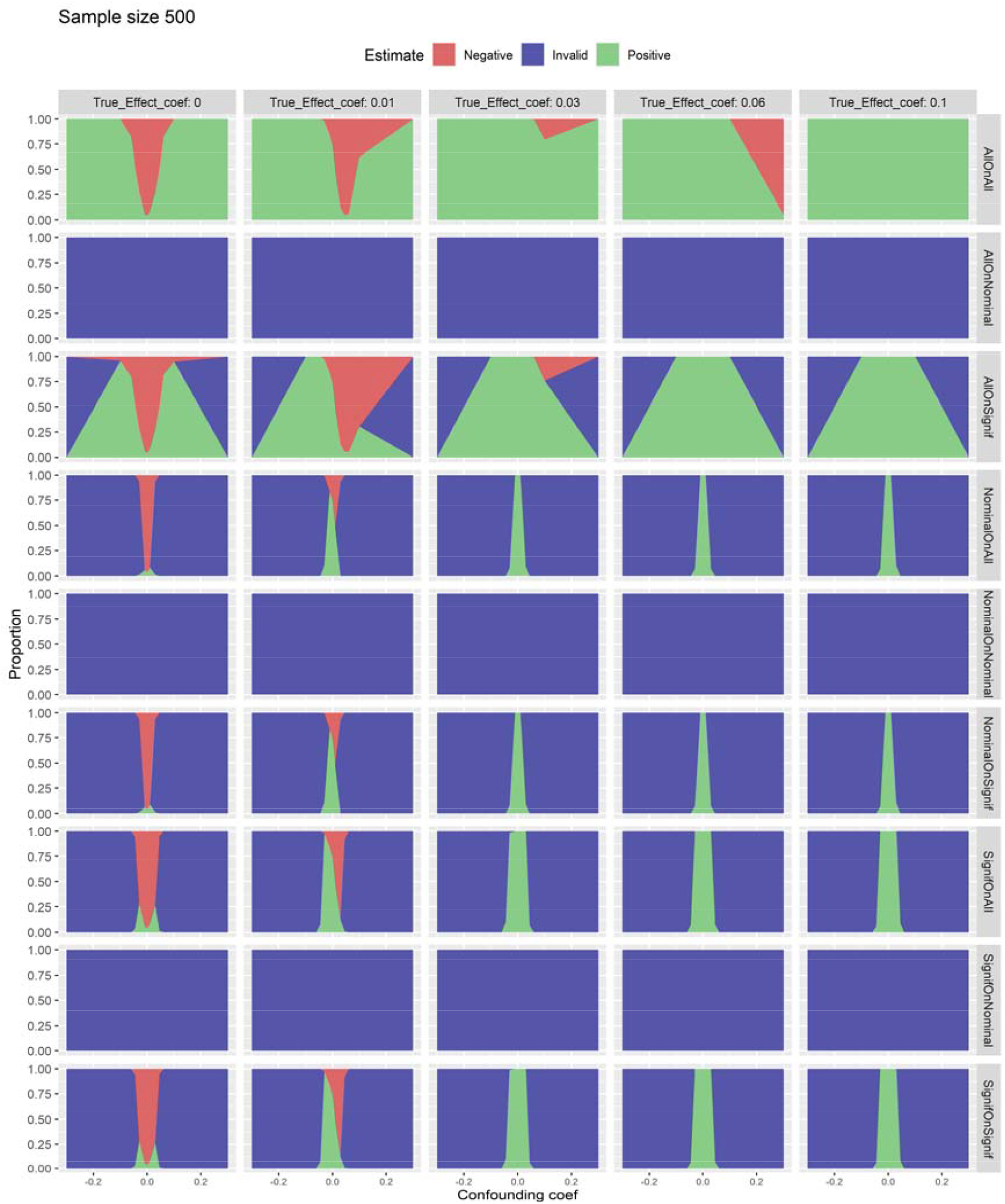
Three-state rule performance at the network level on simulation with sample size 500. Graphs show the proportion of studies that were positive (null hypothesis rejected), invalid (study rejected), and negative (null hypothesis not rejected) on a single-database study with sample size 500. The proportion is plotted against confounding coefficient *c*_*t*_ from –0.3 to 0.3, and graphs from left to right show different values for effect coefficient *c*_*e*_ from 0 to 0.1. The nine rows represent the nine rules listed in the text. The first column should have no more than 0.05 positive (green), and at *c*_*t*_=0, it should have no invalid (blue). The other columns should have positive as high as possible, and at *c*_*t*_=0, it should have no invalid with increasing invalid as confounding increases. See the text for an explanation of results.

## 4 REAL-WORLD DATA

### 4.1 Real-world data design

We exploited a data set and protocol used in two previous studies to illustrate the effect of choice of covariate balance detection on real-world covariates and confounding. The Observational Health Data Sciences and Informatics (OHDSI) ^41,42^ LEGEND hypertension ^5^ and type-2 diabetes studies ^7^ comprehensively evaluated the comparative effects of all pharmaceutical treatments for their respective disease areas. Here we select two comparisons from each, including the negative control outcomes used in these studies.

We used three databases shown in Table 2. Each database uses OHDSI’s Observational Medical Outcome Partnership (OMOP) common data model ^43^ populated with patient characteristics, health care visits, diseases, medications, procedures, and, optionally, other data types such as laboratory tests. Data elements were translated to standard terminologies ^44,45^ such as Systematized Nomenclature of Medicine (diseases, procedures), RxNorm (medications), and Logical Observation Identifiers Names and Codes (laboratory tests).

**Table 2.** Description and Characteristics of Administrative Claims Data Sources

| **Data Source** | **Claims Type** | **Population Enrolled** |
| --- | --- | --- |
| Merative Medicare Supplemental Database (MDCR) | Adjudicated health insurance claims of retirees with primary or Medicare supplemental coverage through privately insured fee-for-service, point-of-service or capitated health plans. | 10M starting 2000<br>Commercially insured, 65+ years |
| Merative MarketScan Multi-State Medicaid Database (MDCD) | Adjudicated health insurance claims for Medicaid enrollees from multiple states and includes hospital discharge diagnoses, outpatient diagnoses and procedures, and outpatient pharmacy claims. | 26M starting 2006<br>Medicaid enrollees, racially diverse |
| Optum® de-identified Electronic Health Record data set (Optum® EHR) | Clinical information, prescriptions, lab results, vital signs, body measurements, diagnoses and procedures derived from clinical notes using natural language processing. | 93M starting 2006<br>US, general |

The gold standard was the collection of 110 real negative controls used in the original studies as well as synthetic positive controls generated from them. Each negative control contained a target drug, a comparator drug, and an outcome that was determined through review of the literature, product labels, spontaneous reports, and clinical experts not to be causally associated with either drug. ^46^ Therefore, the hazard ratio of the appearance of the outcome between the two drugs should be 1, indicating no difference. Synthetic positive controls were generated from the negative controls by fitting an L1-regularized Poisson regression model ^47^ on presence of the outcome based on all pre-treatment covariates and then using that model to insert additional simulated events into the data set to achieve hazard ratios of 1.5, 2, and 4. More details are available. ^5,7^

For each of the three large databases, we created several sets of smaller databases to illustrate the effect of a network study across databases with small sample sizes. We kept the total number of cases constant at 20,000: 5 databases with 4000 cases each, 10 databases with 2000 cases each, 20 databases with 1000 cases each, 40 databases with 500 cases each, and 80 databases with 250 cases each. For each of these sets, we calculated large-scale propensity scores ^26^ for four treatment comparisons—lisinopril versus hydrochlorothiazide, lisinopril versus metoprolol, sitagliptin versus liraglutide, and sitagliptin versus glimepiride—on 98,681 covariates found in the three databases. Only pretreatment variables were used to minimize the probability of mediators or colliders, suspected instruments were removed, and lack of strong instruments was confirmed by measuring equipoise. We calculated propensity scores two ways: on each small database and on a pooled sample of all 20,000 cases. We used three methods to apply the propensity score to a causal analysis: no adjustment (crude), 1-to-1 matching, and stratification. We then used a Cox proportionate hazards model to estimate the hazard ratio for each hypothesis, calculated a confidence interval, and tested for significance based the confidence interval excluding no effect (hazard ratio of 1).

The result was a set of analyses characterized by which of three large databases it came from, by the four hypotheses, by the sample size of the generated database (250, 500, 1000, 2000, 4000), by the underlying hazard ratio (1, 1.5, 2, 24), and by the analysis method (unadjusted=crude, matched on the database-level propensity score, stratified on the database-level propensity score, matched on the pooled propensity score, stratified on the pooled propensity score). Each analysis had an effect estimate and a standardized mean difference for every covariate.

We then further carried out a meta-analysis as described in the Simulation methods section across the 80, 40, 20, 10, or 5 generated data sets (for 250, 500, 1000, 2000, and 4000 sample sizes, respectively) getting meta-analytic estimates for the effect and all the covariate SMDs. We applied the nine covariate balance rules described above in the Simulation methods section. We calculated the proportion of studies that passed the covariate balance rule and report the type 1 error rate and power for each rule for each sample size and for two groups of analyses: unadjusted=crude versus all propensity-adjusted methods. The unadjusted analyses should display greater confounding than the adjusted ones.

### 4.2 Real-world data results

Figure 5 shows the type 1 error (RR=1) and power (RR>1) for all nine rules on the real-world data for different sample sizes and levels of adjustment (adjusted versus unadjusted, likely reflecting lower and higher residual confounding). When covariate imbalance was ignored (rule AllOnAll), the type 1 error rate was high at 0.09 for the unadjusted study (no covariate adjustment and therefore presumably higher confounding) when sample size was 250. As was observed in the simulation, when covariate balance was checked at the database level using a threshold of the standardized mean difference nominally exceeding 0.1 (rules AllOnNominal, NominalOnNominal, and SignifOnNominal), all studies were rejected resulting in no power. Checking at the network level for the meta-analytic standardized mean difference nominally exceeding 0.1 (rules NominalOnAll, NominalOnNomimal, and NominalOnSignif) also resulted in rejecting most studies with reduced power. This left only rules that checked for statistically significant covariate imbalance. Of note, for rule AllOnSignif, which only checks for covariate imbalance at the database level, the type 1 error rate was high at 0.07 when sample size was 250 with the crude analysis, reflecting the consequence of ignoring network-level estimates of standardized mean difference. Rules SignifOnAll and SignifOnSignif, which test for statistically significant imbalance at the network level, both produced type 1 error less than 0.05 and showed power similar to no checking (rule AllOnAll) in the adjusted studies (when confounding was likely to be low) and showed low power due to appropriately rejecting unadjusted studies (when confounding is likely to be high).

**Figure 5.**
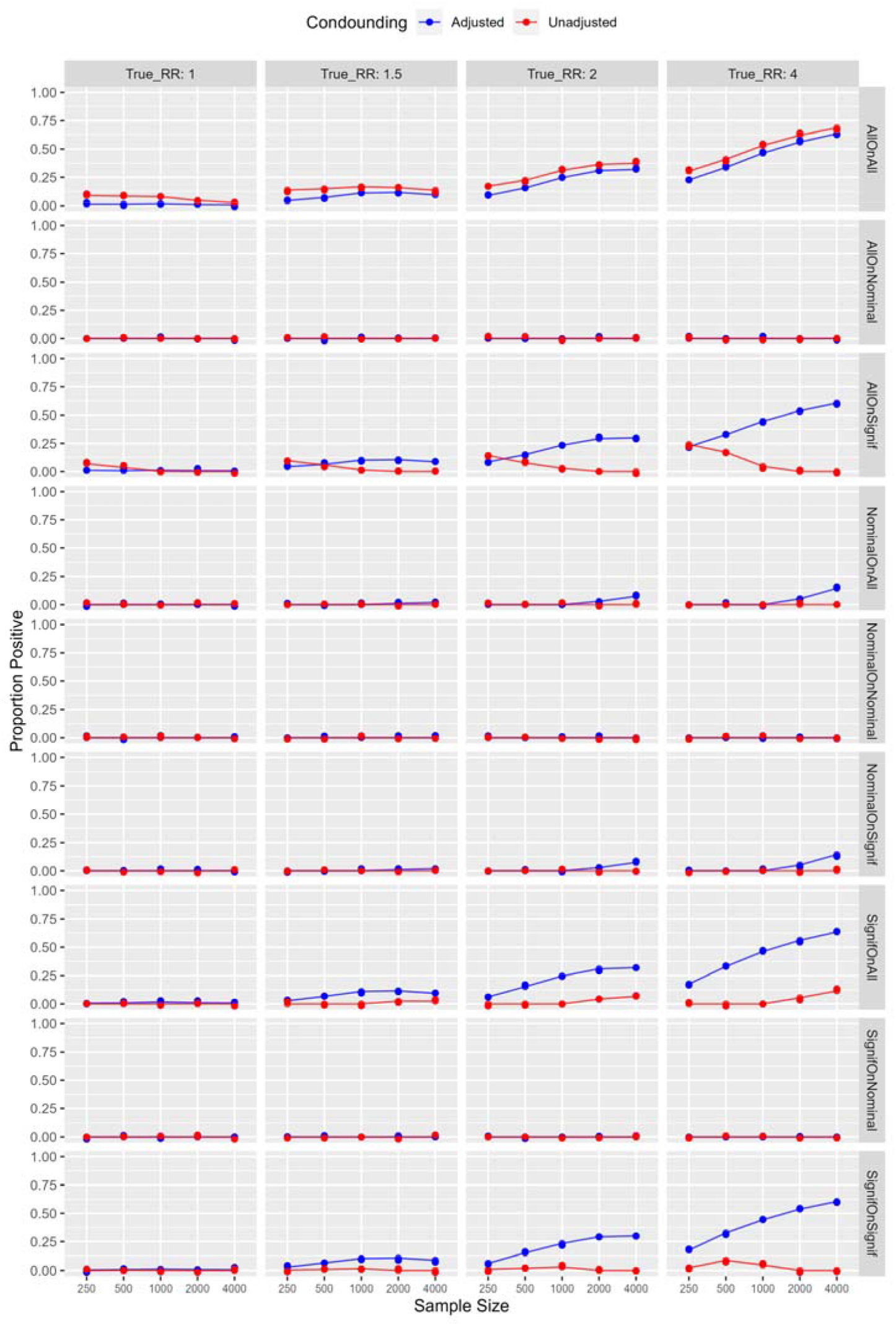
Rule performance at the network level on real-world data. Graphs show the proportion of study iterations that were not rejected by the rule and that had effect coefficients that were statistically significantly different from zero plotted against database sample size. Colored lines represent adjusted and unadjusted analyses, reflecting lower and higher residual confounding. Graphs from left to right show different true relative risks (RR). The nine rows represent the nine rules listed in the Methods section. The first column, where RR=1, shows the type 1 error rate, and the other columns, where RR>1, show the power with increasing effect size. See text for an explanation of results. (Graph points are jittered to reveal overlapping colors but lines are drawn true.)

Figures 6 and 7 show the 3-state results for the real-world data for the adjusted and unadjusted analyses, respectively. In Figure 6, where confounding is expected to be lower due to propensity score adjustment, AllOnAll, AllOnSignif, SignifOnAll, and SignifOnSignif show good control of false positives when the relative risk is 1, and they show similar moderately high power when the relative risk is above 1. In Figure 7, where confounding is expected to be higher due to lack of adjustment for confounding, AllOnAll and AllOnSignif both climb above 0.05 in their false positive rate at small sample sizes when relative risk is 1, whereas SignifOnAll and SignifOnSignif control the false positive rate and generally detect the imbalance that is likely due to confounding.

**Figure 6.**
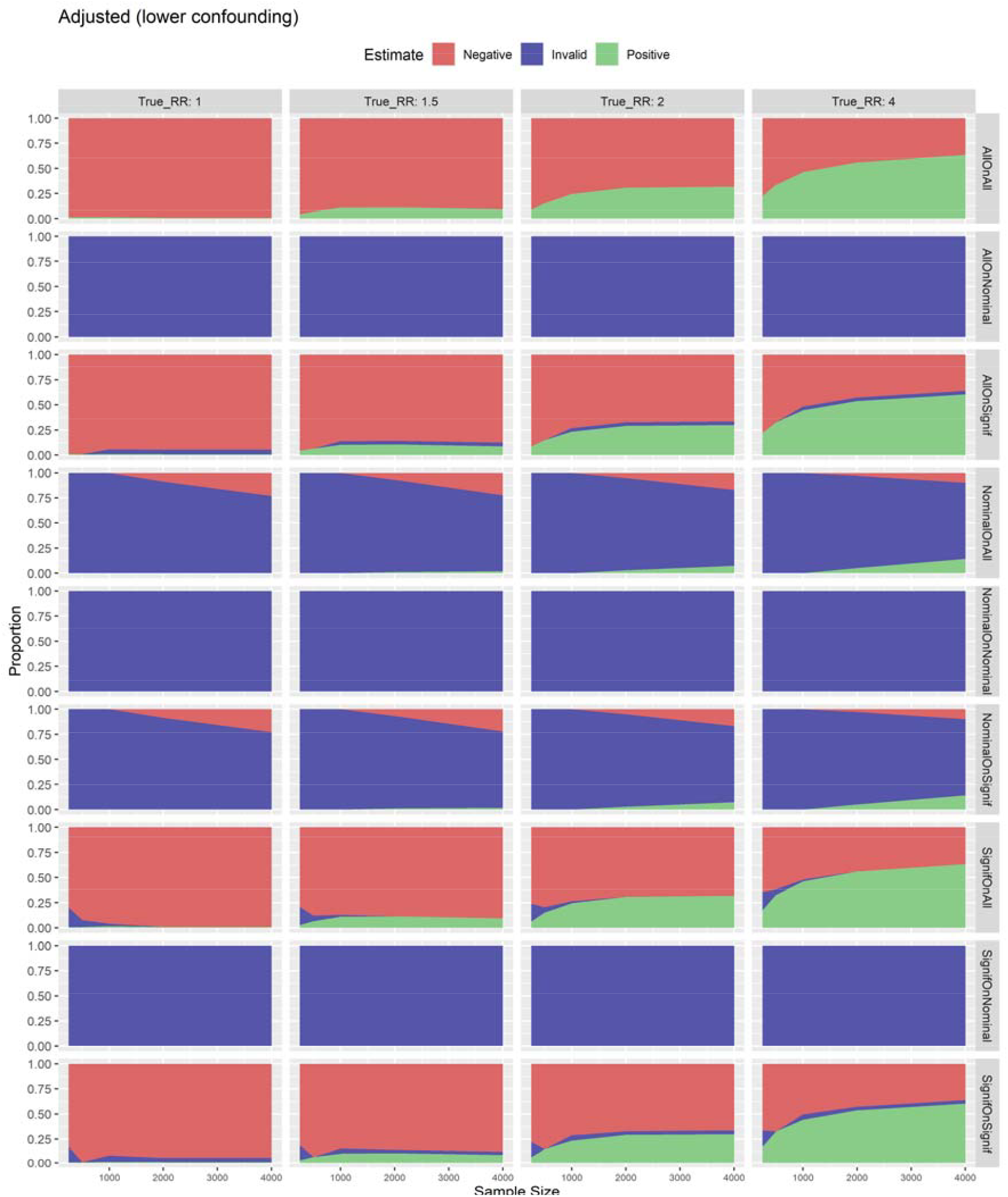
Three-state performance at the network level on real-world data on an adjusted study. Graphs show the proportion of studies that were positive (null hypothesis rejected), invalid (study rejected), and negative (null hypothesis not rejected) on the real-world data in an adjusted (less confounded) analysis. The proportion is plotted against database sample size, and graphs from left to right show different true relative risks (RR). The nine rows represent the nine rules listed in the text. The first column, where RR=1, should have positive (green) no more than 0.05. The other columns, where RR>1, should show as much positive as possible. In those columns, invalid (blue) may represent residual confounding, although low positive in the first column would point against that. See the text for an explanation of results.

**Figure 7.**
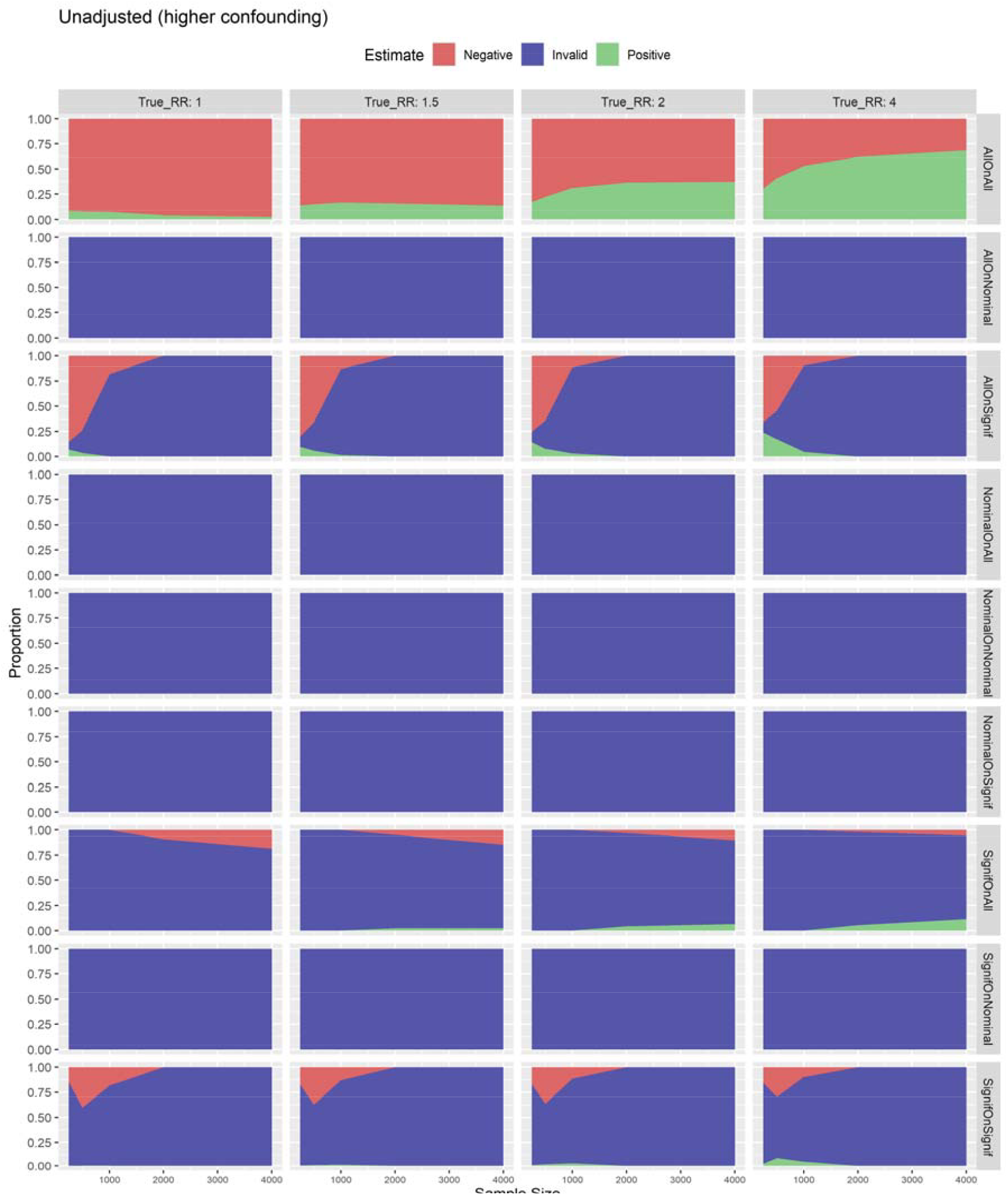
Three-state performance at the network level on real-world data on an unadjusted study. Graphs show the proportion of studies that were positive (null hypothesis rejected), invalid (study rejected), and negative (null hypothesis not rejected) on the real-world data in an unadjusted (more highly confounded) analysis. The proportion is plotted against database sample size, and graphs from left to right show different true relative risks (RR). The nine rows represent the nine rules listed in the text. The first column, where RR=1, should have positive (green) no more than 0.05. The other columns, where RR>1, may have significant invalid (blue) given the level of confounding and otherwise should show positive. See the text for an explanation of results.

## 5 DISCUSSION

Our study illustrates two principles and identifies an algorithm that appears to best address the tradeoff between type 1 error and power in the simulation and real-world data. First, as sample size decreases, using a nominal test of imbalance such as described by Austin ^11^ will result in over-calling of consequential imbalance and near-certain rejection of the study even with no confounding due to chance imbalance, even if that chance imbalance does not significantly raise the type 1 error rate. This effect occurs with few covariates as well as many covariates. Using a statistical test of imbalance exceeding a threshold like 0.1 will maintain power without substantially raising the type 1 error rate. When doing a study with small sample sizes, it may be hard to detect small-to-moderate confounding, but our results illustrate that the small study’s wide confidence intervals will avert a high type 1 error rate. That is, it takes significant confounding to cause high type 1 error rate. Second, when doing a network study, it is important to carry out a meta-analysis not just of the effect estimate but also of the diagnostics such as standardized mean difference. The meta-analysis of the effect estimate may potentially produce a more precise effect estimate and a narrow confidence interval so that small-to-moderate confounding can yield too many false positive results. The meta-analysis of the standardized mean differences, however, permits the detection of lower levels of confounding despite the sample sizes being individually small.

Best overall performance appeared to be achieved by testing for statistically significant imbalance defined as standardized mean difference over 0.1 and using a Bonferroni correction across covariates. For network analyses, rule SignifOnSignif, which tested for statistically significant imbalance at both the single database level and across the database network using a meta-analysis of standardized mean differences, worked best. It produced power at all studied sample sizes, and its power was generally not too much less than ignoring imbalance, yet it achieved substantially lower type 1 error rates. In the simulation, we used iterative parameter testing to find a combination that produced a type 1 error rate approaching 0.3, but we emphasize that we could achieve such high error rates even with standard practice. For example, even in a much larger study with a sample size of 20,000, using only 20 covariates, setting c_t_ to 0.02, c_y_ to 0.2, and c_e_ to 0, and using a nominal threshold of 0.1, the type 1 error rate was 0.57. Given any threshold, it is possible to design a simulation that thwarts it.

The most important question is what happens with real-world data, and in fact, rule SignifOnSignif produced a type 1 error rate that was always less than or equal to 0.05 and power almost as high as not checking for imbalance at all. Rule SignifOnSignif had other good properties. If the confounding was heterogeneous, doing the standardized mean difference test at both levels worked best: drop the databases that fail at the individual level and do a meta-analysis on the rest. As the number of databases varied from 1 to 5 up to 80, rule SignifOnSignif produced a stable output, and it worked whether the number of covariates was 20, 1000, or 98,681. When covariate prevalence or outcome prevalence was low, the results were similar.

Rules that appeared to work on simulation failed on the real-world data. For example, rule NominalOnAll, which tested for any meta-analytic standardized mean difference to be nominally over 0.1, had too-low power in real-world data compared to testing for statistical significance: even with 4000 cases in a study with low confounding, most studies were rejected, rendering the rule ineffective for small studies. It also failed in the simulation when the number of databases in the network was only 5. The two techniques that most clearly failed represent standard practice: doing a nominal test for imbalance at the database level (AllOnNominal) rejected all databases regardless of confounding as sample size fell; and doing no test for imbalance (AllOnAll) led to very poor type 1 error as confounding rose.

Applying meta-analysis to study diagnostics will clearly be difficult given that very few studies share the details of their balance results (although it has been suggested ^48^). The approach is still relevant to distributed networks that use meta-analysis to combine their results; sharing of study diagnostics can be incorporated into the study protocol. ^49^ It is important to at least recognize this limitation of meta-analysis on observational research without study diagnostics.

Accepting studies in which confounding is non-zero can be seen as misguided. Should not the metric instead be the ability to detect confounded studies? We believe that power is the right metric. As Austin ^11^ showed, the goal is not to detect increasingly minute imbalance, but to detect imbalance that might reflect confounding that matters. As sample size falls, confidence intervals widen, and it takes more confounding to alter the result appreciably. If important confounding is slipping through the imbalance test, then that will be reflected in the type 1 error rate. If the type 1 error rate stays near its nominal value (say 0.05), then we argue that power reflects acceptance of studies that while they might be confounded to some degree, that confounding is insufficient to cause a frequent false result. All observational studies in fields like medicine have some small degree of confounding, so it is important that the goal be recognized as achieving best power given a nominal type 1 error rate, not identifying and eliminating all studies.

This study potentially informs other observational research as well. The current custom is to check balance only on the covariates suspected to be confounders and adjusted for using propensity scores, ignoring potentially useful information about other covariates. Given our results, it may be more informative to incorporate all available covariate information. If a covariate is unbalanced, that should be explained as an instrument if domain knowledge is available or the study should be suspected to be biased. Our work demonstrates that it is possible to test for imbalance without triggering too many false positives and also without missing confounding that could substantially affect the results.

Our approach is similar to that of Han and Sidell, who define pseudo p-values. ^39^ The difference is mainly in the goal. They calculate the probability of producing a sample as imbalanced or more than the observed sample and use it as a descriptive statistic. We seek to create a thresholded heuristic to identify studies to be rejected or otherwise handled, and we judge it based on its performance in type 1 error and power. Whether that imbalance is due to systematic confounding or excessive chance imbalance of an outcome predictor in the current sample, our heuristic provides useful information. Han and Sidell’s null hypothesis is zero imbalance, and our null hypothesis is imbalance no more than the 0.1 threshold. We also address meta-analysis, whereas Han and Sidell do not.

As authors have pointed out, ^11,19,50,51^ using a statistical test to detect the presence of imbalance (difference from 0) performs poorly and does not achieve the proper goal, yet we believe that this test for exceeding a threshold (exceed 0.1) is in fact useful. Imai et al. ^50^ argue three points: that a statistical test is dependent on sample size yet the actual imbalance is not, that any threshold like a p-value is arbitrary, and that the target of analysis is the sample itself and not some underlying population. On the first, we too are dependent on sample size but we believe that that is appropriate. As the size of the sample shrinks, the effect estimate will become less precise with a larger confidence interval and any given level of confounding will have a proportionately smaller influence on the conclusion of the study. Therefore, the threshold for imbalance ought to become less stringent as sample size falls. On the second, we agree that a threshold is arbitrary at first, but the observational research field including most of these authors have come to relative agreement on a 0.1 constant threshold ^11-23^ despite the fact that no threshold can guarantee immunity from important bias. The appropriateness of a threshold is decided slowly over time as real study results are compared to baseline knowledge and validated in later randomized experiments. Our use of statistical comparison to 0.1 is no more arbitrary than current practice. On the third, we acknowledge that we are concerned with the current sample and not some underlying population. For example, chance imbalance that is extreme should still be identified. We use the statistical test to adjust the level of the threshold to the sample size, in effect accounting for the size of the effect estimate sample size. Our test may be better seen as a heuristic that uses a threshold whose level varies with sample size. Furthermore, analogous to the sample size dependence of our test, when a binary covariate is rare, there is insufficient power to detect imbalance in that covariate, but that is accompanied by a concomitant lack of power to sway the effect estimate.

Our use of a Bonferroni correction may be questioned, but we argue it is necessary and appropriate. With increasing numbers of independent covariates, the likelihood of chance imbalance will rise, so as we have demonstrated (Figure S5), correction for multiple hypotheses is needed to avoid rejecting too many studies. One may question why existing confounders should be allowed more imbalance just by adding new independent covariates. First, and most important, we include many covariates because small sets of manually chosen or empirically selected confounders are likely missing confounding (both directly and indirectly measured ^26^), which is likely why large-scale propensity adjustment appears to perform better than other methods. ^25,26,36,37^ That is, we agree that domain knowledge is effective for identifying important confounders but we believe it has little ability to rule out other confounders. Our further simulations (see Supplement section III) demonstrate that if confounders are distributed among the covariates, then adding covariates actually increases our ability to detect confounding despite the Bonferroni correction. We also point out that for the range of standard errors of standardized mean difference that we saw in the real-world data (0.044) adding a Bonferroni correction for 100,000 covariates when testing for exceeding at threshold of 0.1 is still stricter than not using Bonferroni correction but using a threshold of 0.25, which is a previously accepted alternative threshold. ^19,20^ In the end, we believe that the question of how many covariates to include is an empirical one, balancing the benefit of covering more confounders with the risk of including inappropriate variables, and our experience so far is that more has been better. ^25,26,36,37^ This experiment shows that on real-world data, chance imbalance can be addressed by rule SignifOnSignif even with 98,861 covariates. Nevertheless, whether a researcher selects 20 covariates or 100,000 covariates, the results of this experiment remain relevant. As noted above, researchers may consider less conservative corrections for multiple hypotheses than Bonferroni; ^40^ such modifications will need to be tested on whether they produce a better type-1-error versus power tradeoff in real-world data.

One can also argue whether observational studies with small sample sizes can be trusted. They are clearly important because even in large databases, uncommon treatments can result in small cohorts. As we demonstrate in our experiment, such studies can achieve type 1 error rates and power comparable to larger studies. With modern regularized regression, ^47^ even propensity models with 100,000 covariates can be stably estimated with sample sizes down to 250. ^52^ Therefore, we see no need to exclude them.

## Data Availability

All data produced in the present work are contained in the manuscript. The underlying clinical databases on which analyses were run are available by license from the respective data holders indicated in Table 1.

## Notes

**Funding statement**: This work is partially supported through US National Institutes of Health grants (T15 LM007079, R01 LM006910, and R01 HL169954).

**Conflict of interest statement:** PBR and MJS are employees of Johnson & Johnson. MAS and GH receive contracts from Johnson & Johnson to support methods research not directly related to this study. Johnson & Johnson and Janssen did not have input in the design, execution, interpretation of results or decision to publish. All other authors declare no competing interests.

### Competing Interest Statement

: PBR and MJS are employees of Johnson & Johnson. MAS and GH receive contracts from Johnson & Johnson to support methods research not directly related to this study. Johnson & Johnson and Janssen did not have input in the design, execution, interpretation of results or decision to publish. All other authors declare no competing interests.

### Funding Statement

This work is partially supported through US National Institutes of Health grants (T15 LM007079, R01 LM006910, and R01 HL169954).

### Author Declarations

This study was approved by the Columbia University institutional review board.

### Summary of Updates

This version has clarifications and minor edits.

