## Supplement for "Assessing Covariate Balance with Small Sample Sizes"

### I. Sensitivity analyses on rule performance at the network level using simulation.

Figures S1 to S6 show sensitivity analyses for the base case, shown in Figure 2 in the main paper. Graphs below show the proportion of study iterations that were not rejected by the rule and that had effect coefficients that were statistically significantly different from zero plotted against database sample size. Colored lines represent different levels of confounding $c_t$ from −0.3 to 0.3 (Figure S4 heterogeneous confounding has a single line with confounding mixed). Graphs from left to right show different values for effect parameter $c_e$ from 0 to 0.1. The nine rows represent the nine rules listed in the Methods section. The first column, where $c_e$=0, shows the type 1 error rate, and the other columns, where $c_e$>0, show the power with increasing effect size. See main paper text for an explanation of results. (Graph points are jittered to reveal overlapping colors but lines are drawn true.)

**Figure S1. Rule performance when number of databases is limited to 5.** See caption above.

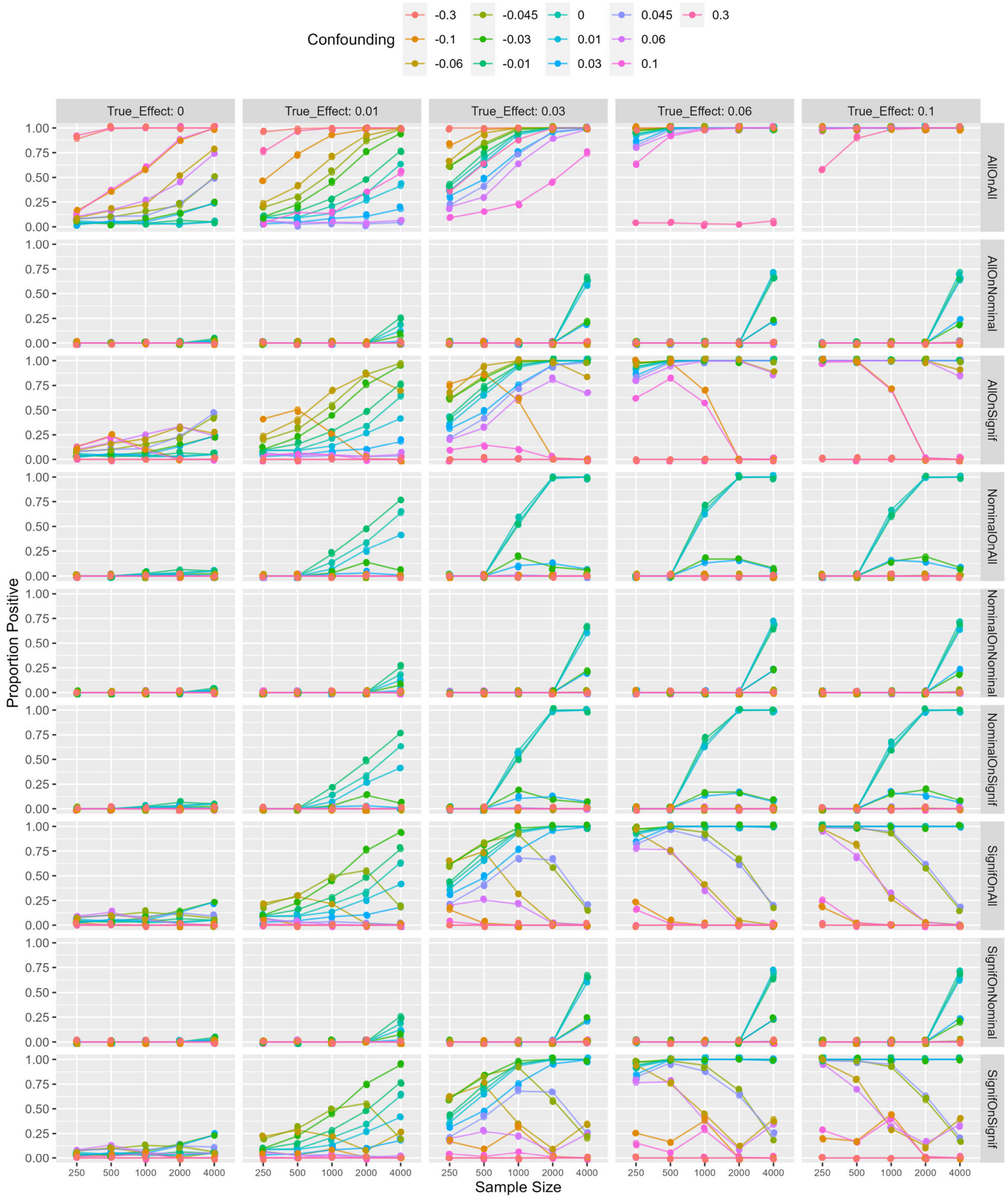

**Figure S2. Rule performance when outcome prevalence is low (1%).** See caption above.

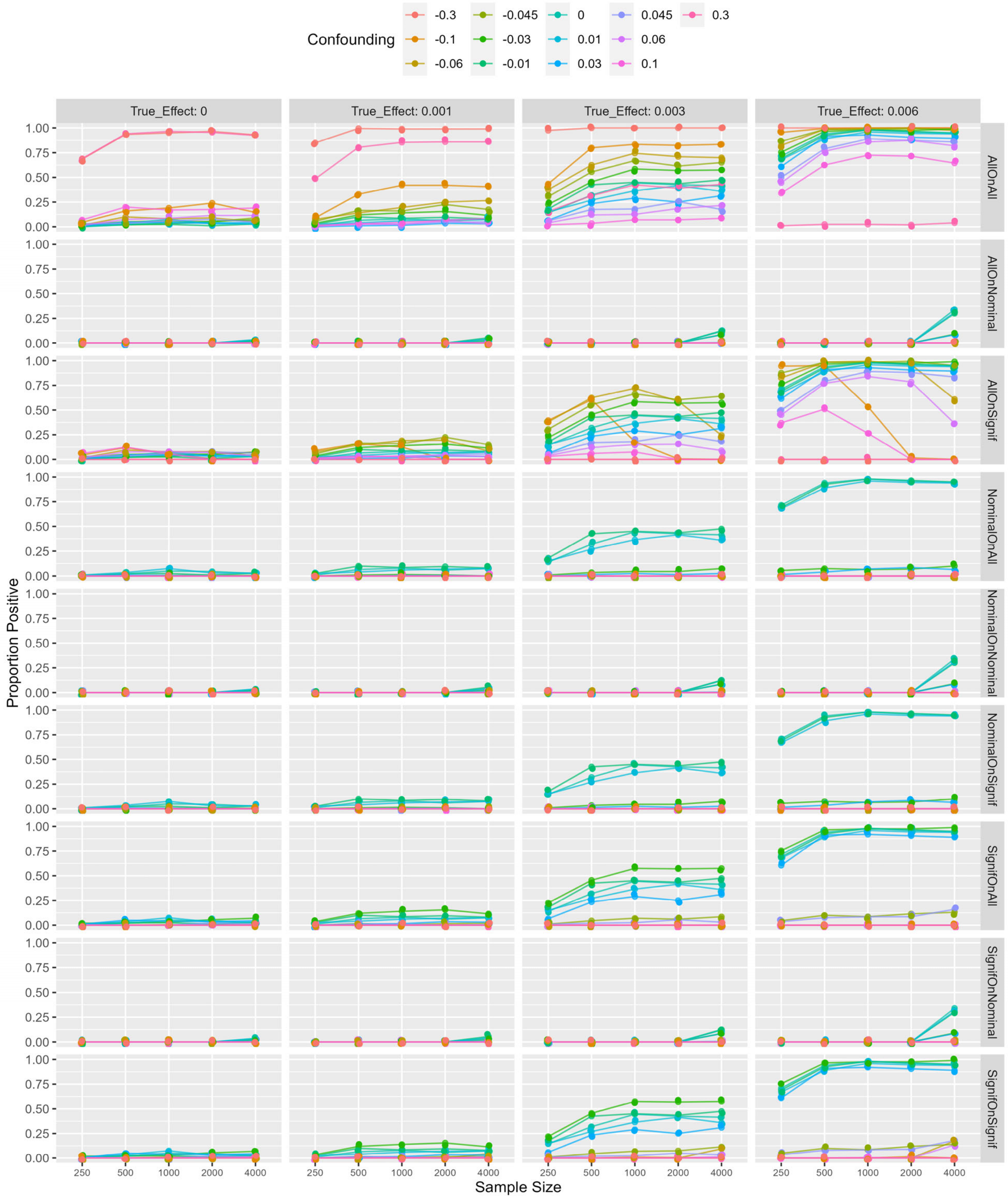

**Figure S3. Rule performance when covariate prevalence is low (10%).** See caption above.

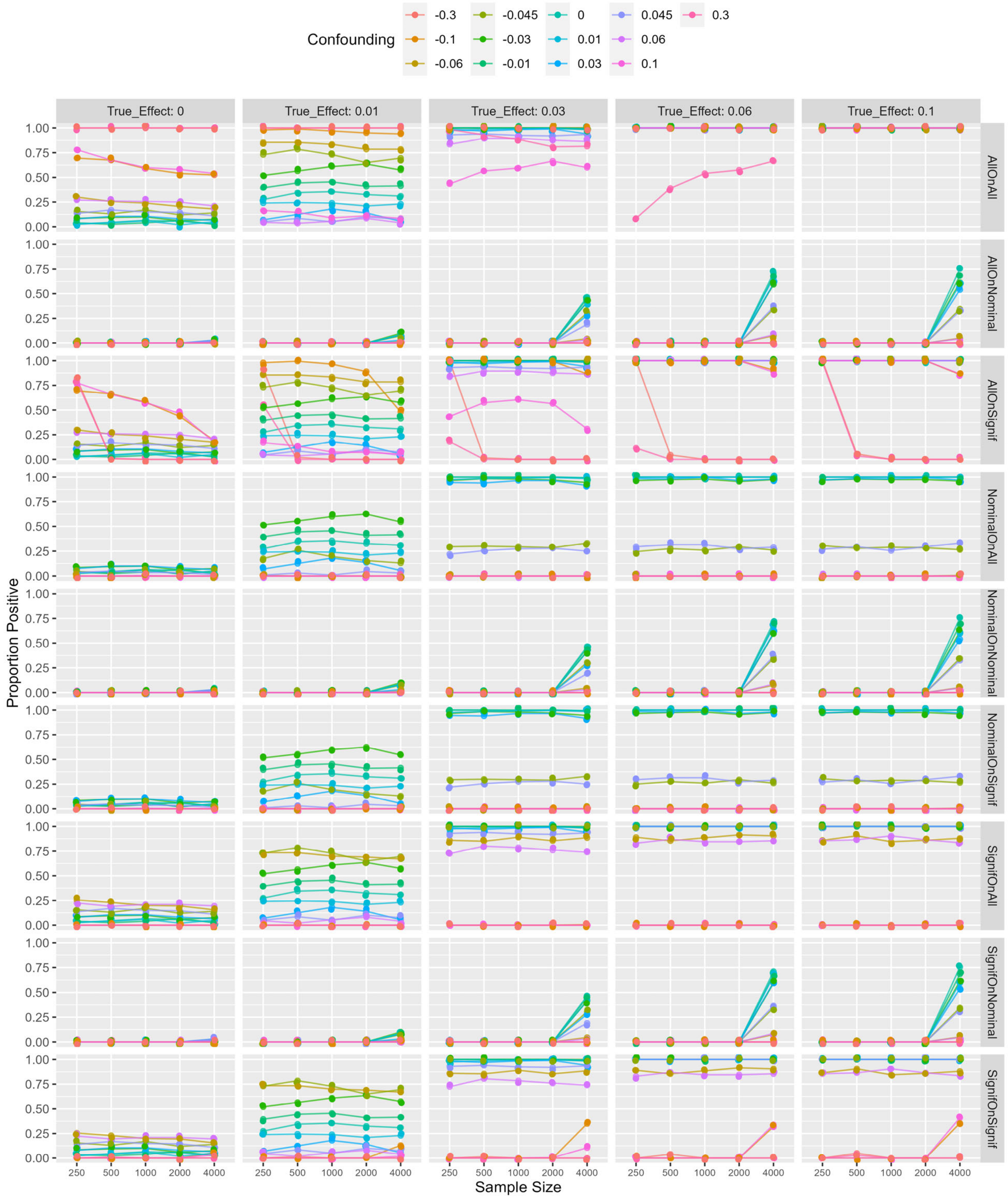

**Figure S4. Rule performance when confounding is heterogeneous ($c_t$ −0.3 to 0.3).**
See caption above.

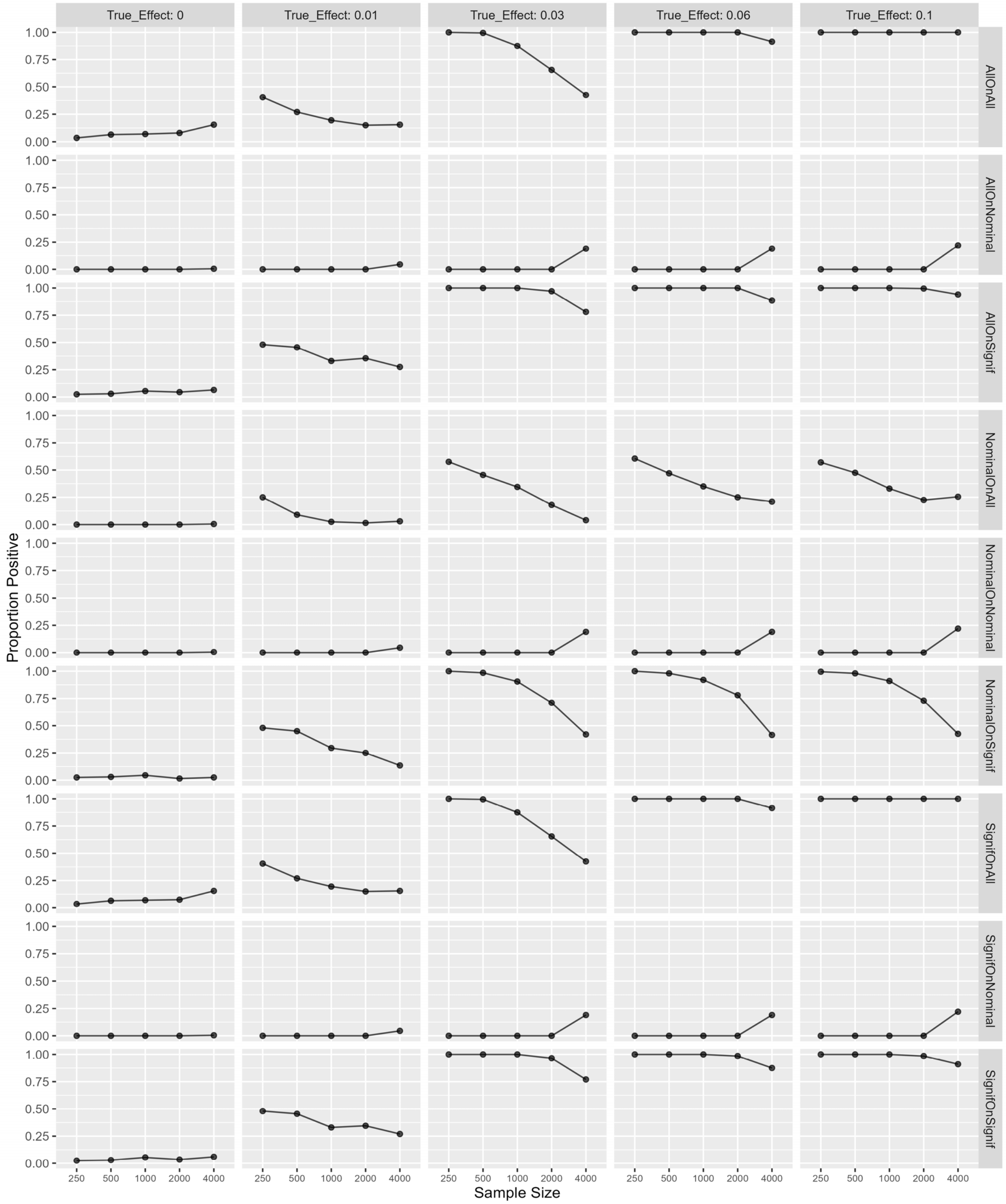

**Figure S5. Rule performance without Bonferroni correction on the Signif rules.** See caption above.

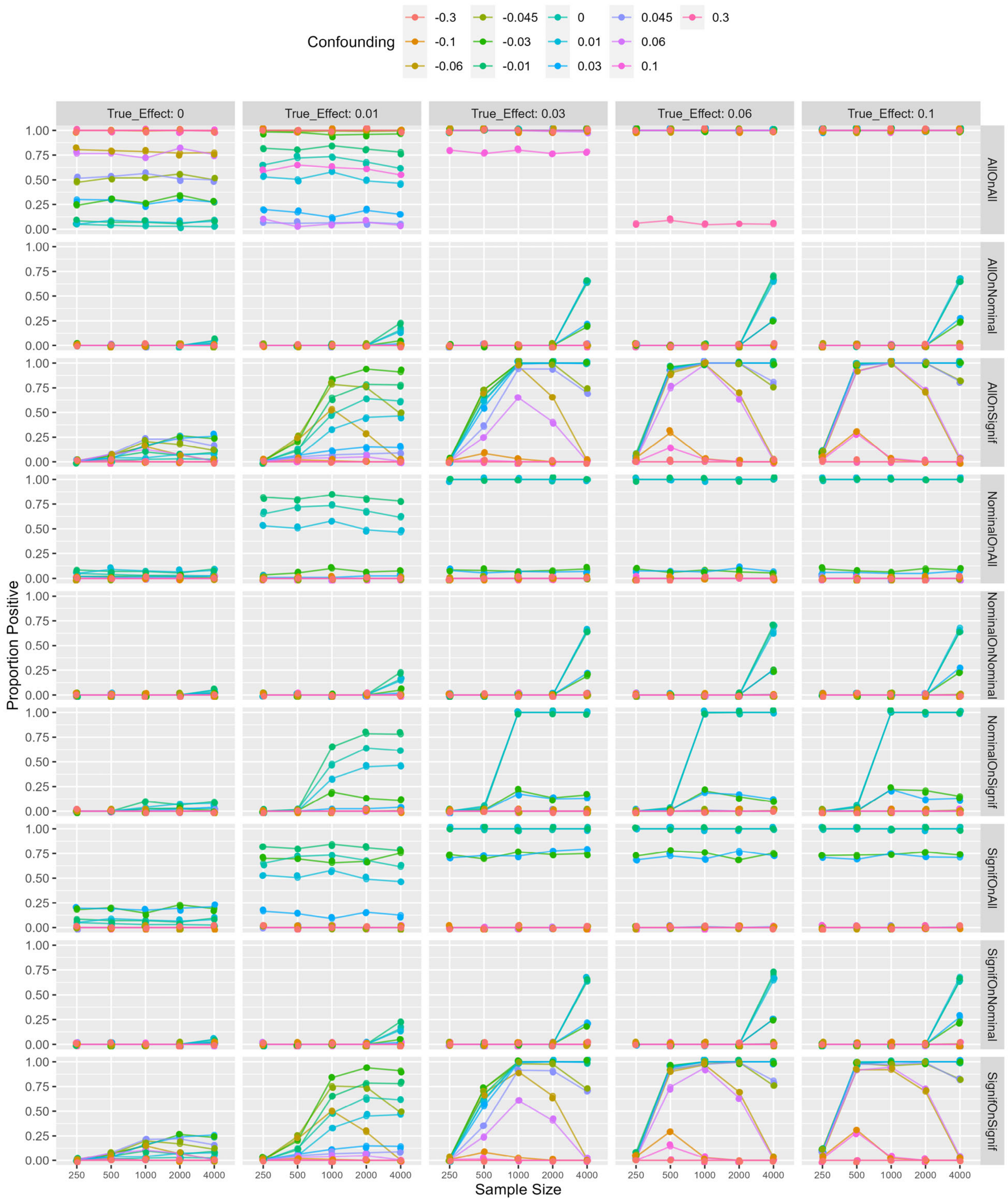

**Figure S6. Rule performance with 20 covariates.** See caption above.

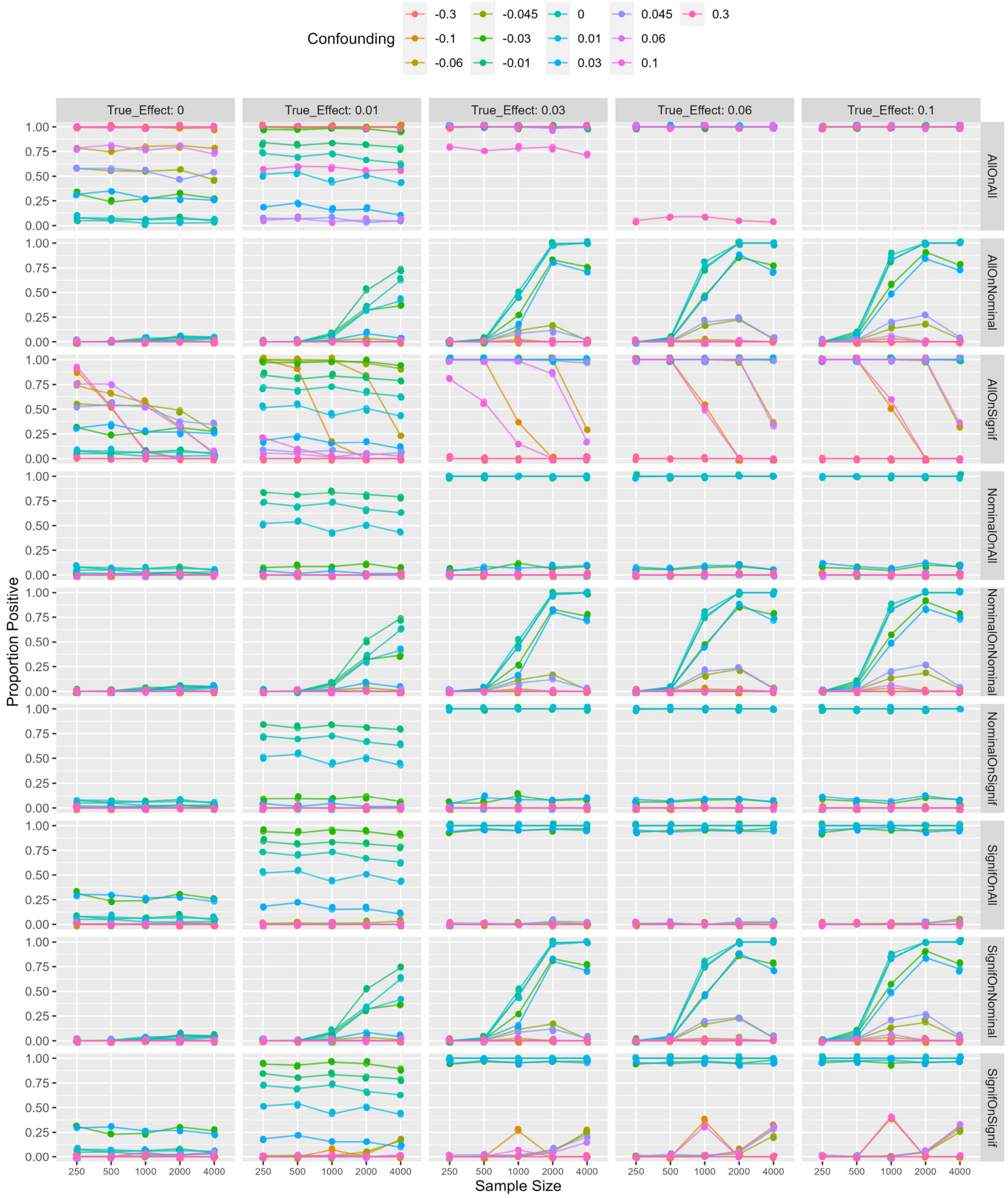

**Figure S7. Rule performance with sample size 20,000 cases and 20 covariates.** Graphs below show the proportion of study iterations that were not rejected by the rule and that had effect coefficients that were statistically significantly different from zero plotted against degree of confounding ($c_t$ from 0.008 to 0.3). Each graph represents one of the nine rules listed in the Methods section. In all graphs, there is no true effect ($c_e$=0), so they all show the type 1 error rate. See main paper text for an explanation of results.

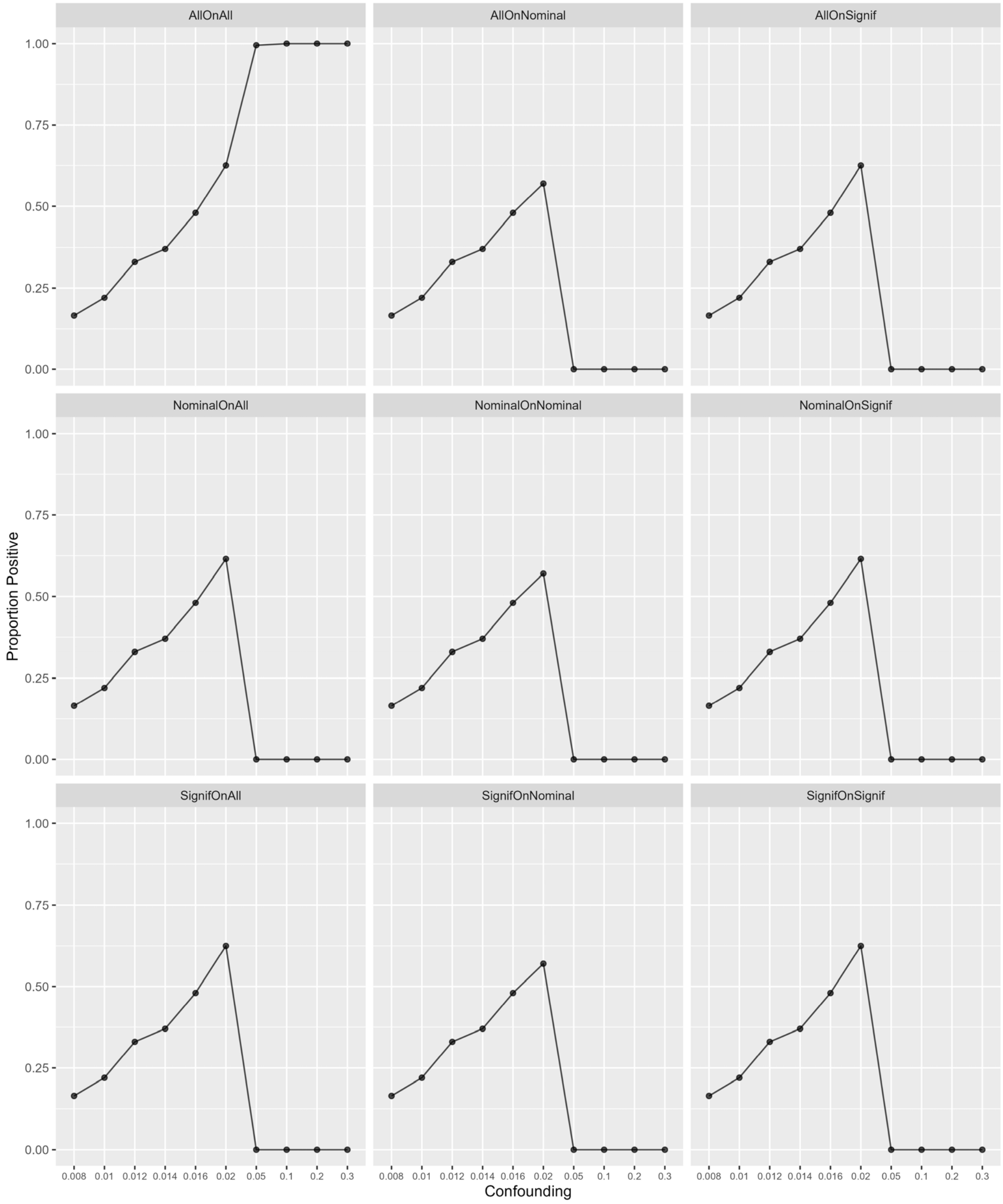

### II. Proportion of studies that were valid (not rejected).

In Figures S8 to S15 and S17, graphs are shown for each level of true effect ($c_e$) for completeness even though the probability of being rejected by the rule is constant with respect to true effect. Minor differences between columns of graphs are due to chance variation among the simulations, and are shown to match against corresponding graphs of rule performance, Figures 1, 2, S1, S2, S3, S4, S5, S6, and 3, respectively.

**Figure S8. Proportion of studies that were valid at the single database level on simulation.** The proportion of study iterations that were not rejected by the rule is plotted against database sample size. Colored lines represent different levels of confounding with $c_t$ from –0.3 to 0.3, and graphs from left to right show different values for effect parameter $c_e$ from 0 to 0.1. The rows represent the three types of rules applied only to a single database under study: **All** ignores imbalance, **Nominal** tests for any covariate's standardized mean difference reaching or exceeding 0.1, and **Signif** tests for any covariate's standardized mean difference statistically significantly reaching or exceeding 0.1. (Graph points are jittered to reveal overlapping colors but lines are drawn true.)

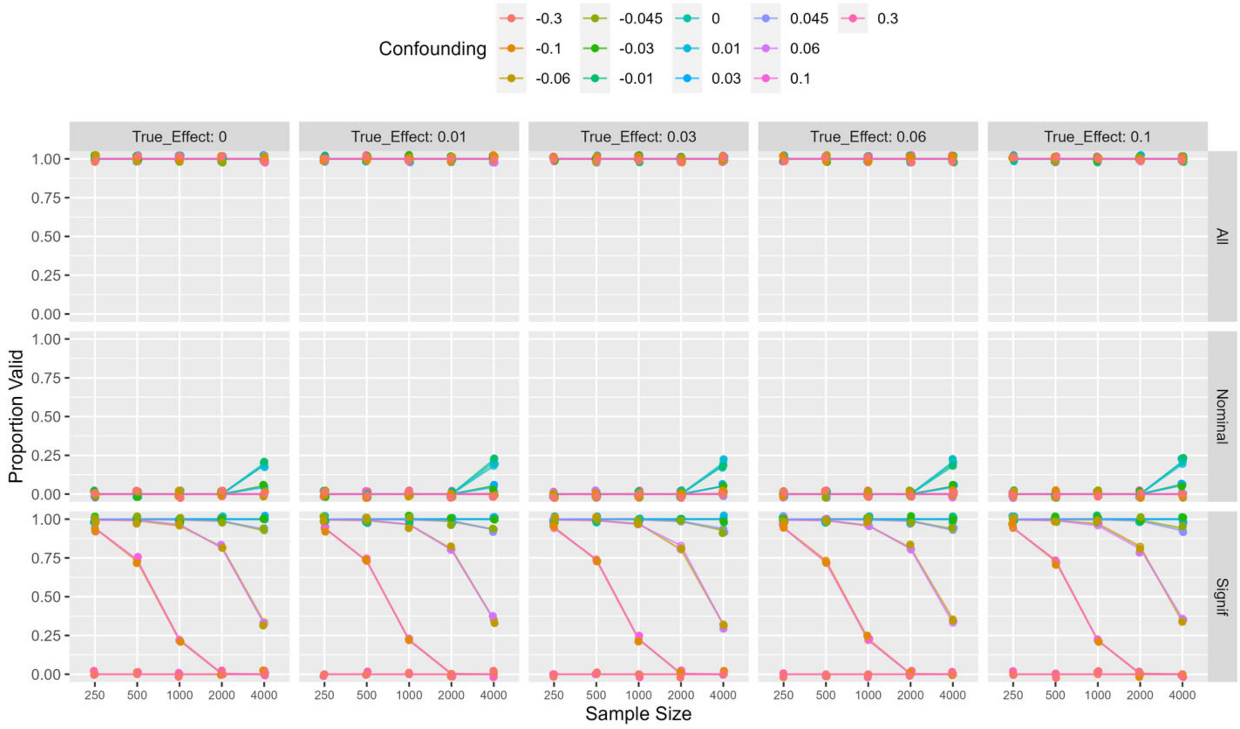

**Figure S9. Proportion of studies that were valid at the network level on simulation for the base case.** Graphs show the proportion of study iterations not rejected by the rule plotted against database sample size. Colored lines represent different levels of confounding $c_t$ from −0.3 to 0.3, and graphs from left to right show different values for effect parameter $c_e$ from 0 to 0.1. The nine rows represent the nine rules listed in the Methods section. (Graph points are jittered to reveal overlapping colors but lines are drawn true.)

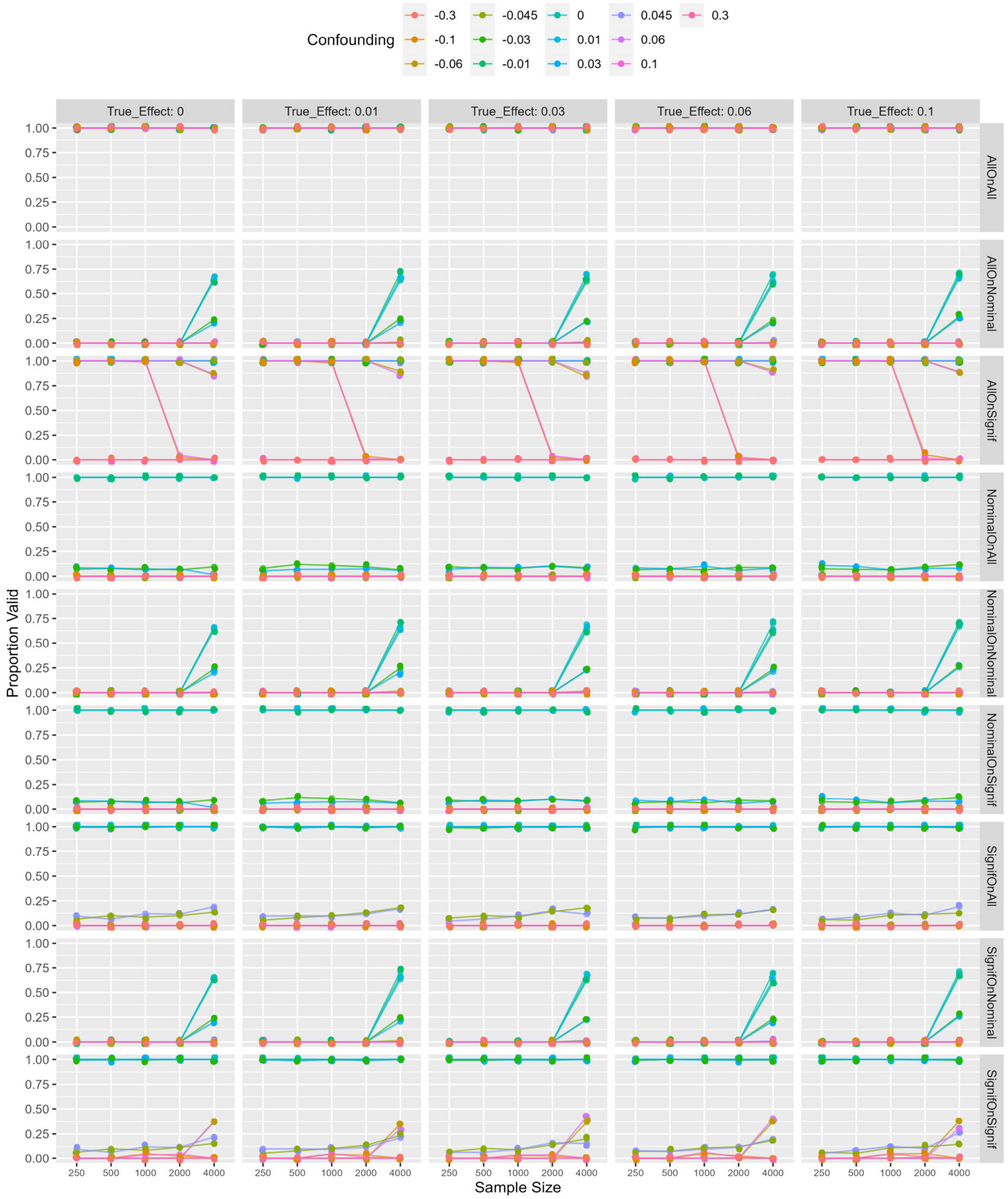

**Figure S10. Proportion of studies that were valid at the network level on simulation when databases are limited to 5.** Graphs show the proportion of study iterations not rejected by the rule plotted against database sample size. Colored lines represent different levels of confounding $c_t$ from –0.3 to 0.3, and graphs from left to right show different values for effect parameter $c_e$ from 0 to 0.1. The nine rows represent the nine rules listed in the Methods section. (Graph points are jittered to reveal overlapping colors but lines are drawn true.)

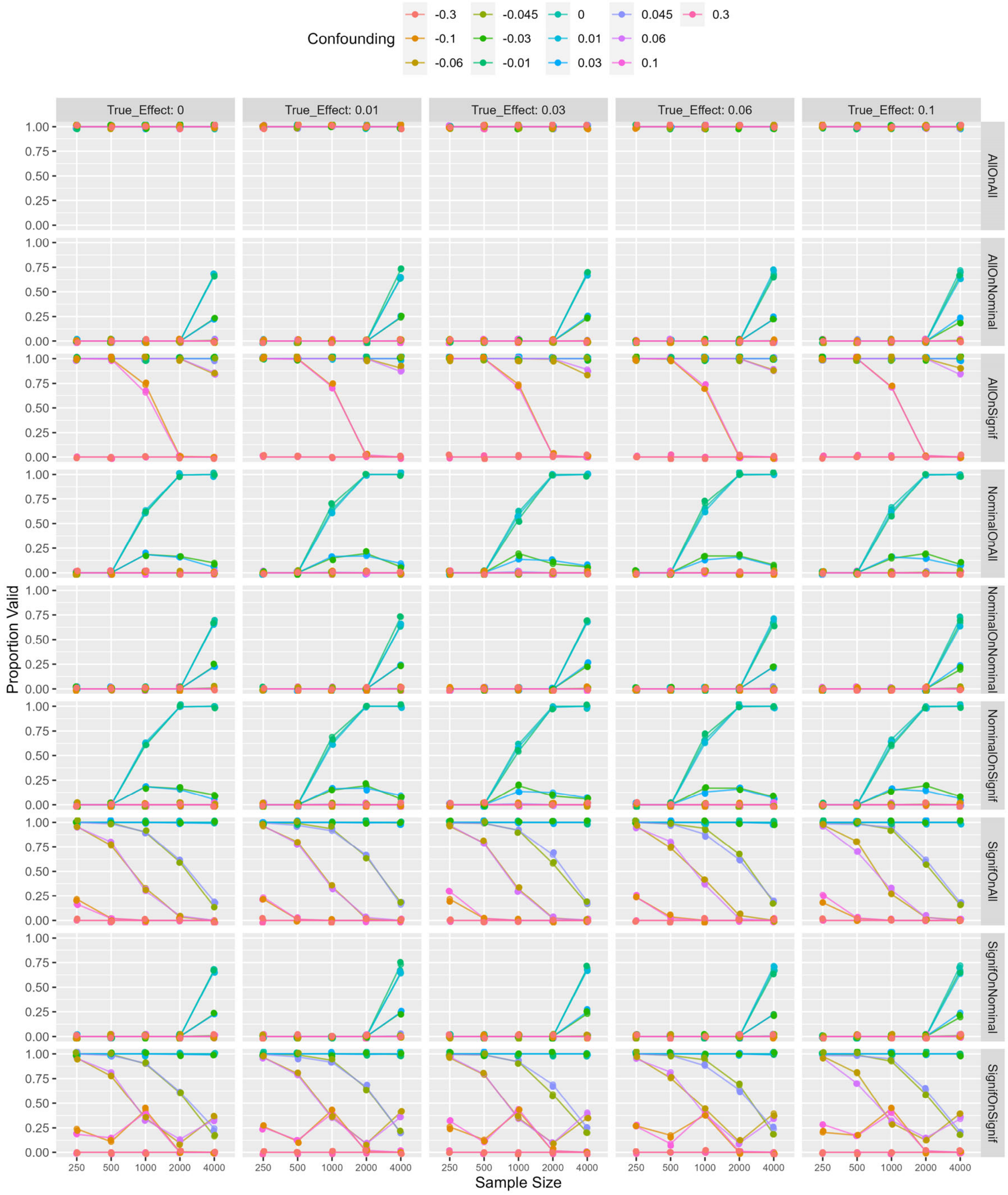

**Figure S11. Proportion of studies that were valid at the network level on simulation with low outcome prevalence.** Graphs show the proportion of study iterations not rejected by the rule plotted against database sample size. Colored lines represent different levels of confounding $c_t$ from −0.3 to 0.3, and graphs from left to right show different values for effect parameter $c_e$ from 0 to 0.1. The nine rows represent the nine rules listed in the Methods section. (Graph points are jittered to reveal overlapping colors but lines are drawn true.)

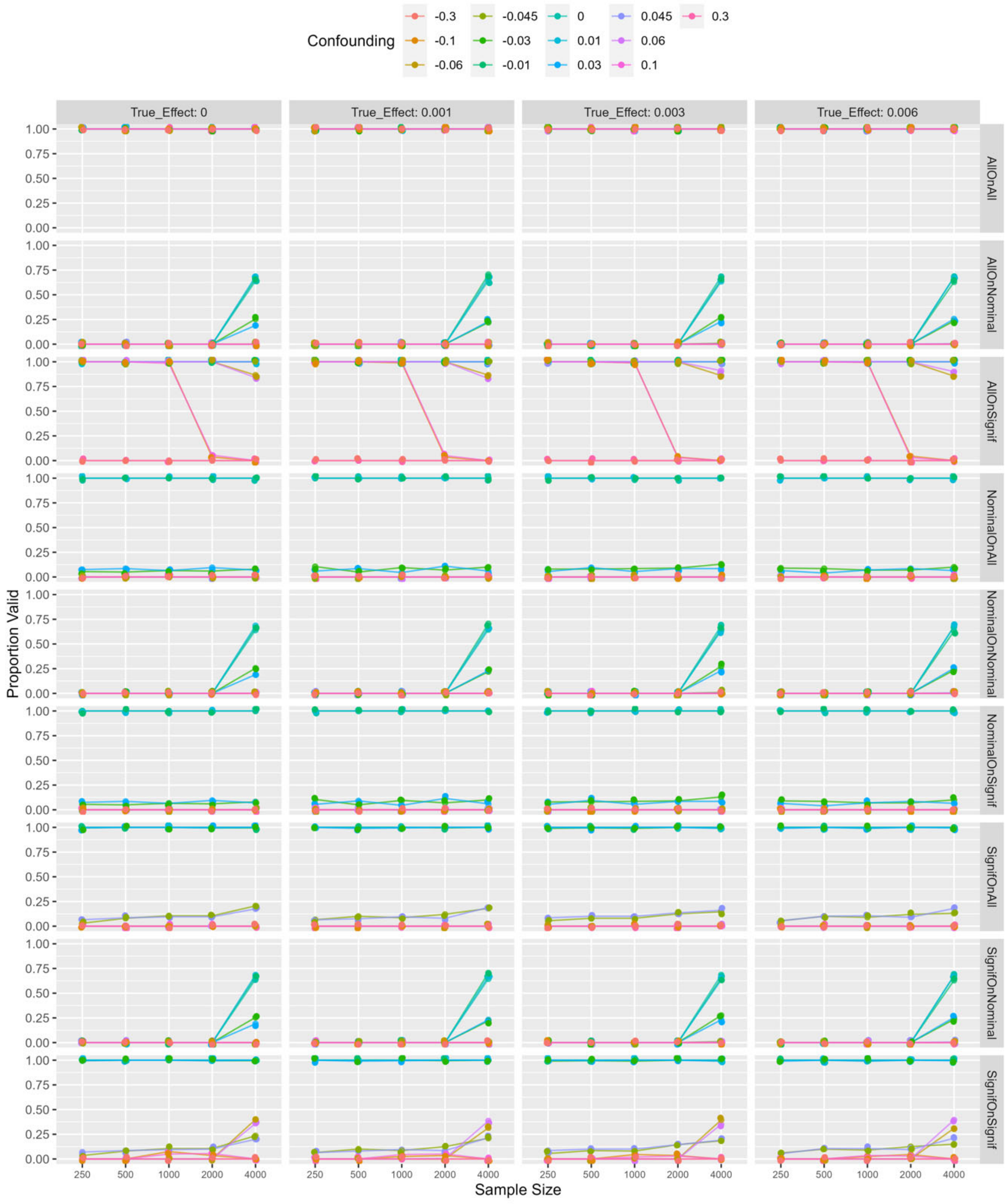

**Figure S12. Proportion of studies that were valid at the network level on simulation with low covariate prevalence.** Graphs show the proportion of study iterations not rejected by the rule plotted against database sample size. Colored lines represent different levels of confounding $c_t$ from −0.3 to 0.3, and graphs from left to right show different values for effect parameter $c_e$ from 0 to 0.1. The nine rows represent the nine rules listed in the Methods section. (Graph points are jittered to reveal overlapping colors but lines are drawn true.)

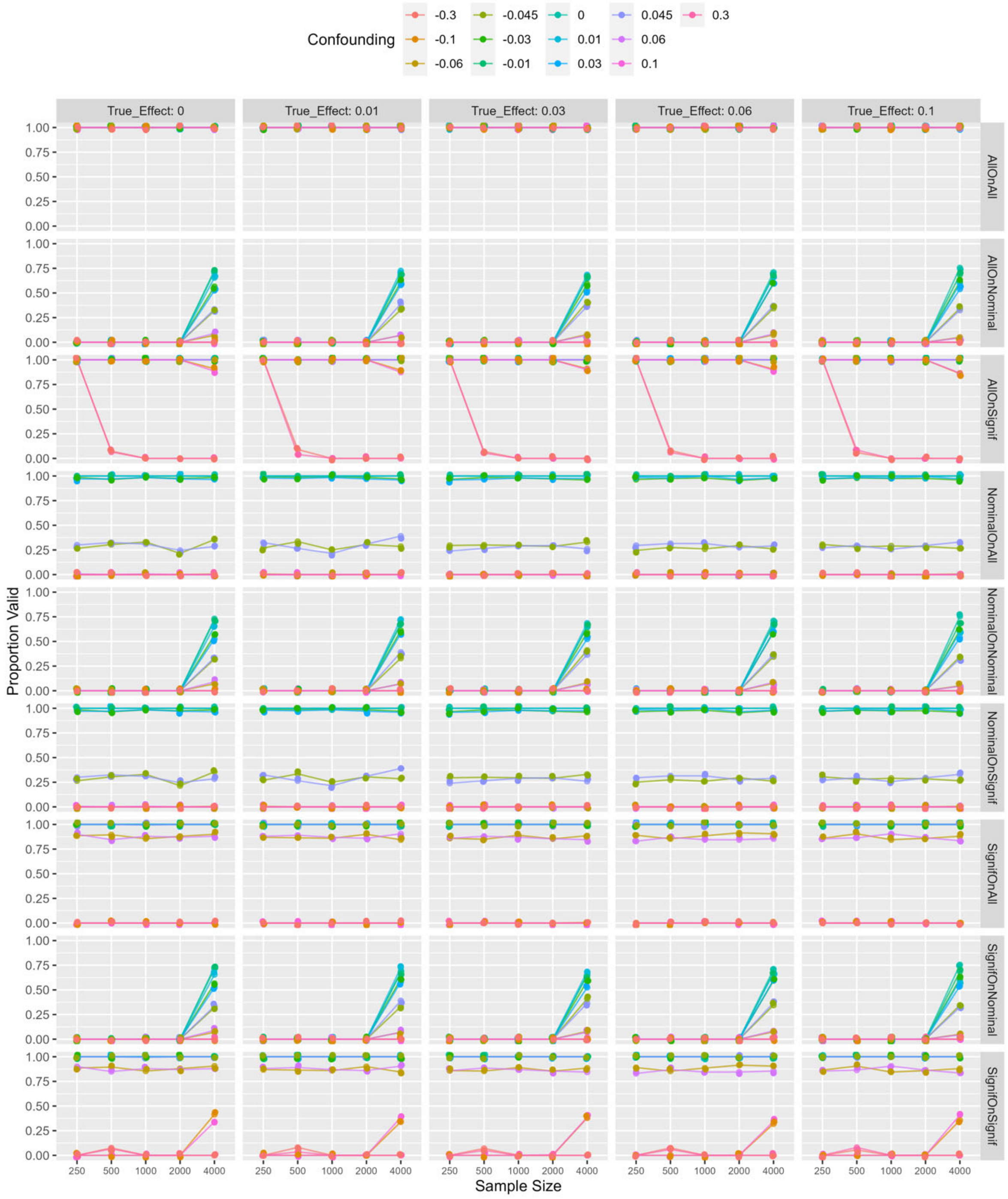

**Figure S13. Proportion of studies that were valid at the network level on simulation with heterogeneous confounding.** Graphs show the proportion of study iterations not rejected by the rule plotted against database sample size. Confounding was heterogeneous ($c_t$ from –0.3 to 0.3). Graphs from left to right show different values for effect parameter $c_e$ from 0 to 0.1. The nine rows represent the nine rules listed in the Methods section.

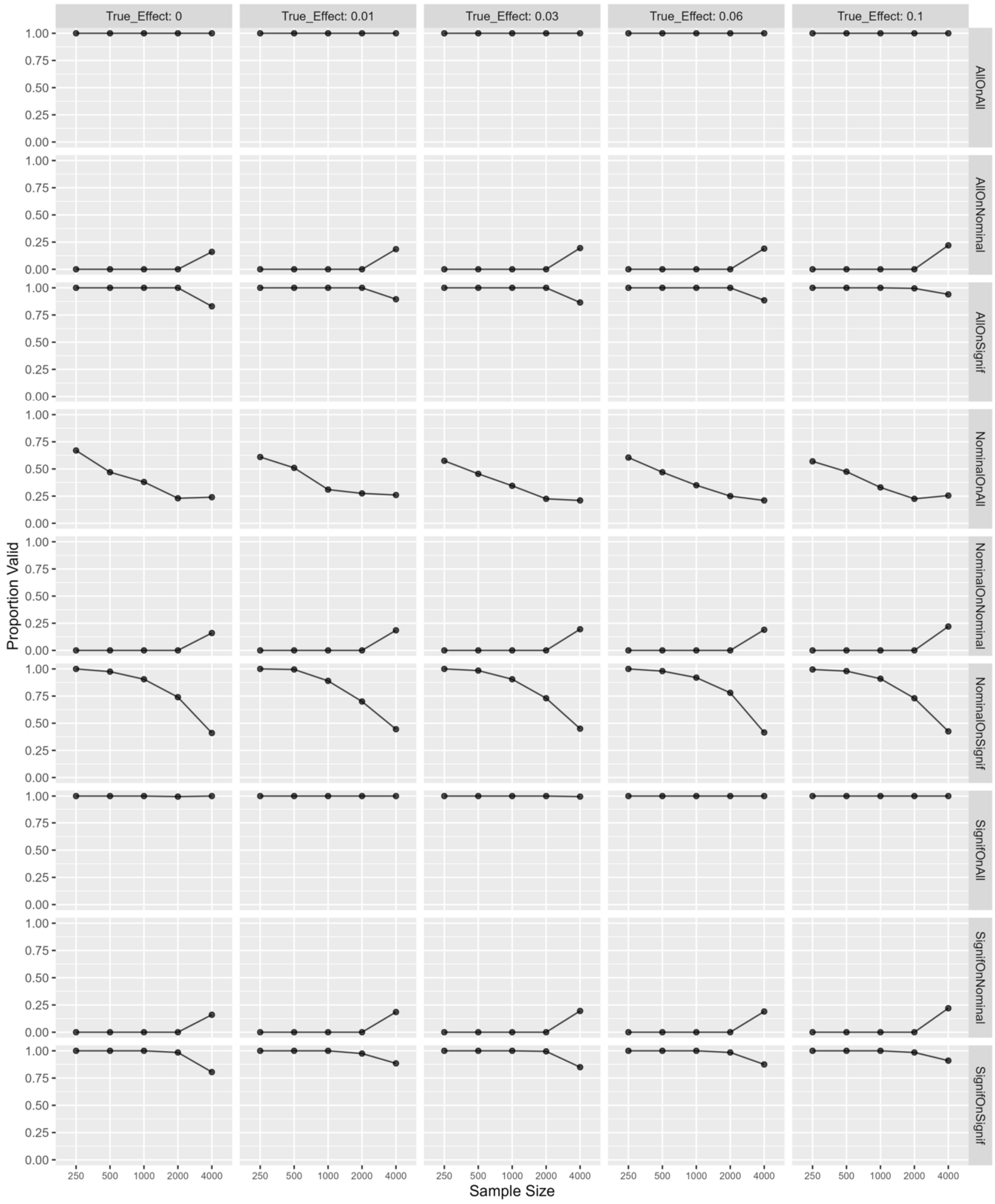

**Figure S14. Proportion of studies that were valid at the network level on simulation without Bonferroni correction on Signif rules.** Graphs show the proportion of study iterations not rejected by the rule plotted against database sample size. Colored lines represent different levels of confounding $c_t$ from –0.3 to 0.3, and graphs from left to right show different values for effect parameter $c_e$ from 0 to 0.1. The nine rows represent the nine rules listed in the Methods section. (Graph points are jittered to reveal overlapping colors but lines are drawn true.)

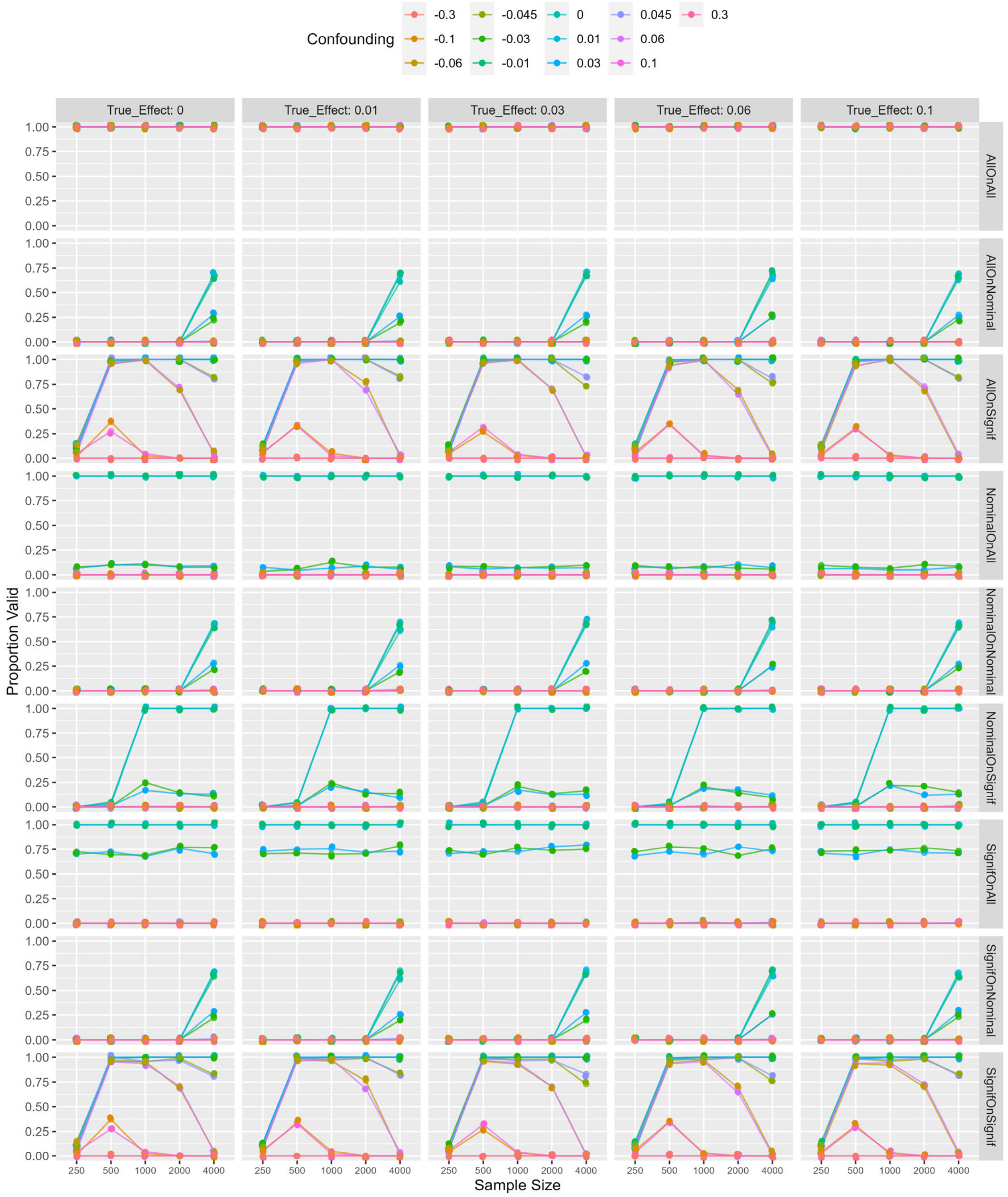

**Figure S15. Proportion of studies that were valid at the network level on simulation with 20 covariates.** Graphs show the proportion of study iterations not rejected by the rule plotted against database sample size. Colored lines represent different levels of confounding $c_t$ from –0.3 to 0.3, and graphs from left to right show different values for effect parameter $c_e$ from 0 to 0.1. The nine rows represent the nine rules listed in the Methods section. (Graph points are jittered to reveal overlapping colors but lines are drawn true.)

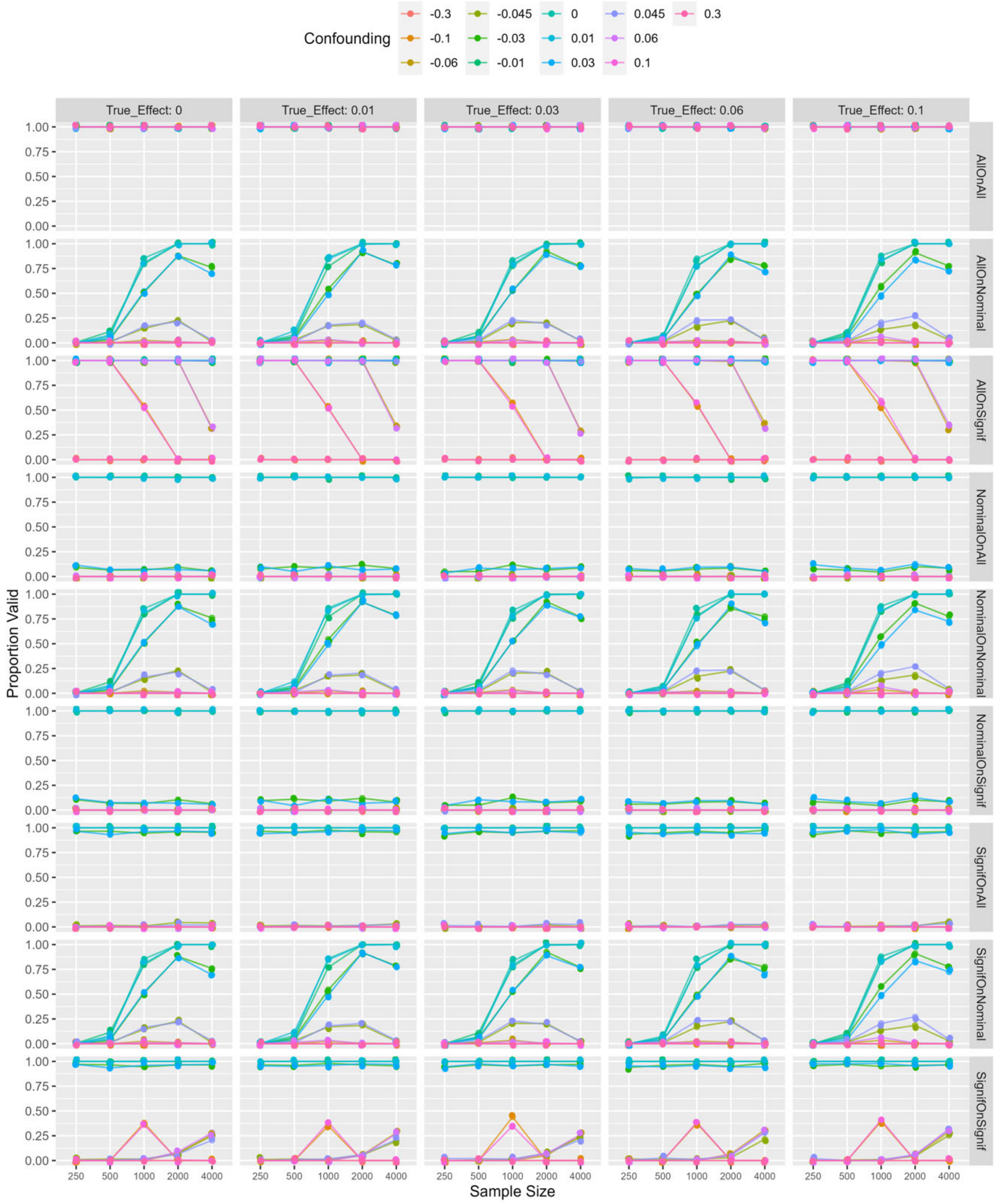

**Figure S16. Proportion of studies that were valid at the network level on simulation with sample size 20,000 cases and 20 covariates.** Graphs below show the proportion of study iterations not rejected by the rule plotted against degree of confounding ($c_t$ from 0.008 to 0.3). Each graph represents one of the nine rules listed in the Methods section. In all graphs, there is no true effect ($c_e$=0).

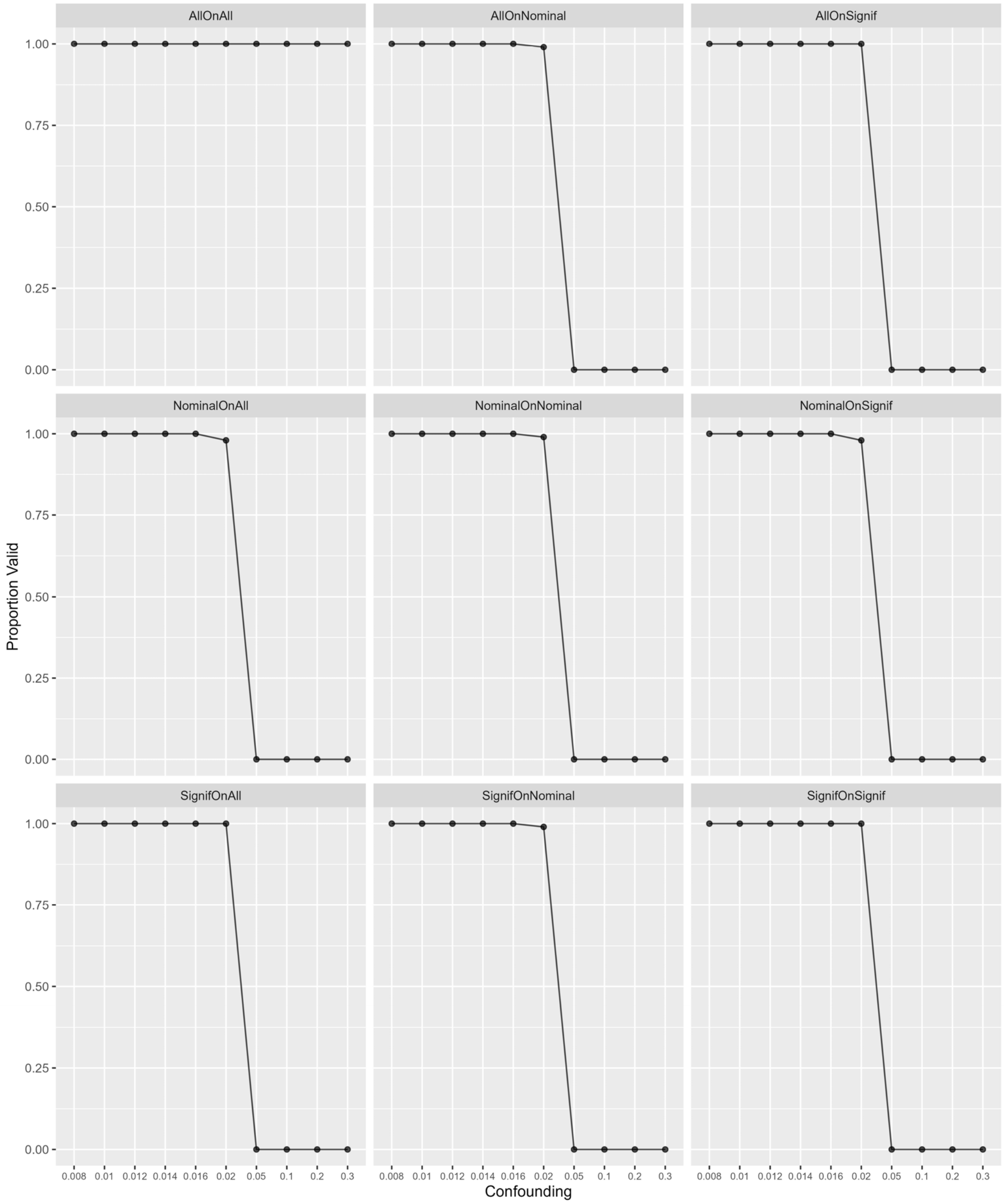

**Figure S17. Proportion of studies that were valid at the network level on real-world data.** Graphs show the proportion of study iterations that were not rejected by the rule plotted against database sample size. Colored lines represent adjusted and unadjusted analyses, reflecting lower and higher residual confounding. Graphs from left to right show different true relative risks (RR). The nine rows represent the nine rules listed in the Methods section. (Graph points are jittered to reveal overlapping colors but lines are drawn true.)

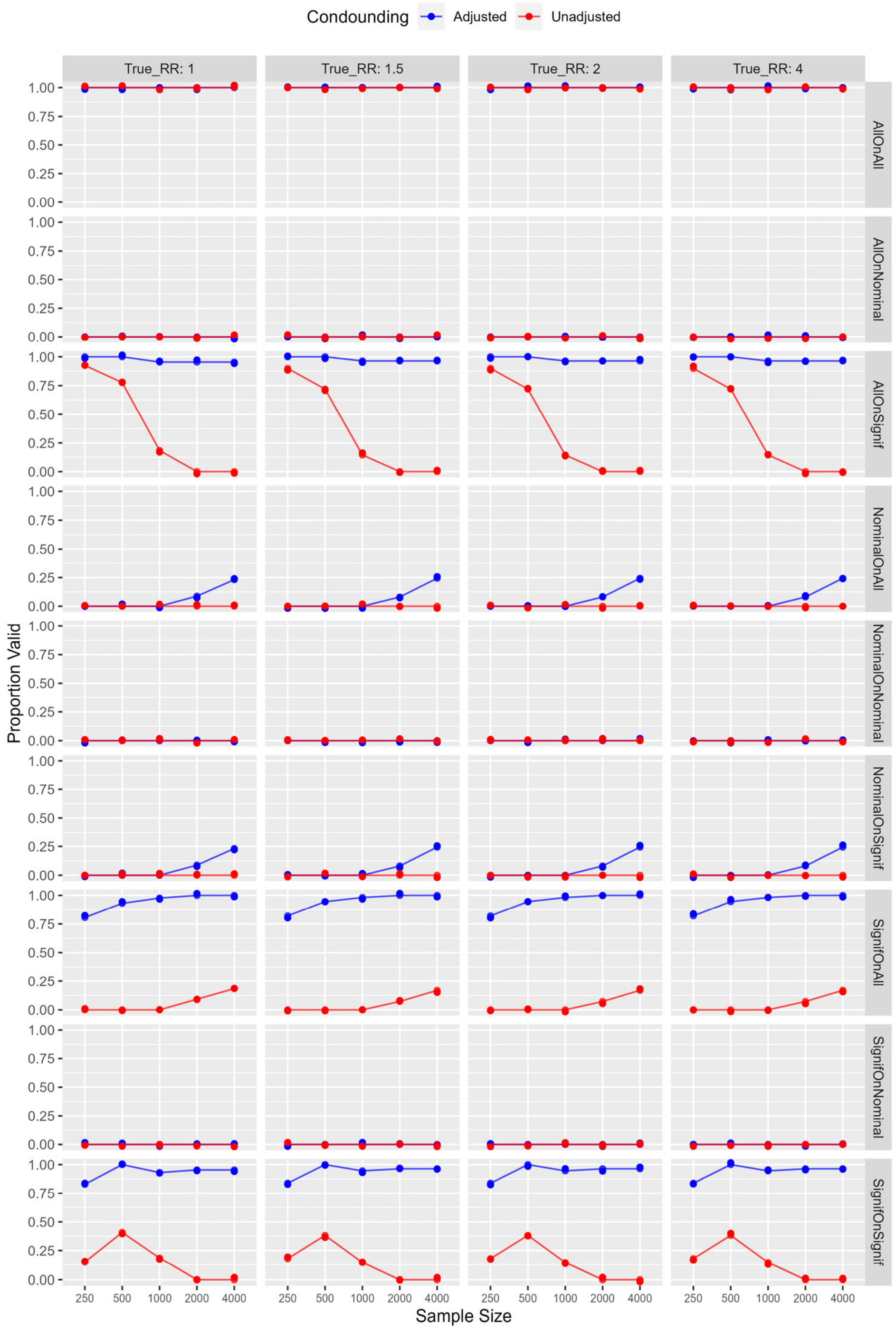

**III. Effect of increasing number of covariates on ability to detect imbalance.**

We carried out simulations of adding covariates under the assumption that confounders are distributed among the covariates (which is the motivation for including them). Given a probability of 0.001 of each covariate being imbalanced and given the degree of imbalance being 0.05, for a sample size of 4000, with 10,000 covariates, there was a 0.62 chance of detecting imbalance and rejecting the study. With 60,000 covariates, rejection rate rose to 1.0.

**IV. Three-state performance**.

**S18. Three-state performance at the database level on simulation with sample size 250.** Graphs show the proportion of studies that were positive (green, null hypothesis rejected), invalid (blue, study rejected), and negative (red, null hypothesis not rejected) on a single-database study with sample size 250. The proportion is plotted against confounding coefficient $c_t$ from –0.3 to 0.3, and graphs from left to right show different values for effect coefficient $c_e$ from 0 to 0.1. The rows represent the three types of rules applied only to a single database under study: **All** ignores imbalance, **Nominal** tests for any covariate's standardized mean difference reaching or exceeding 0.1, and **Signif** tests for any covariate's standardized mean difference statistically significantly reaching or exceeding 0.1.

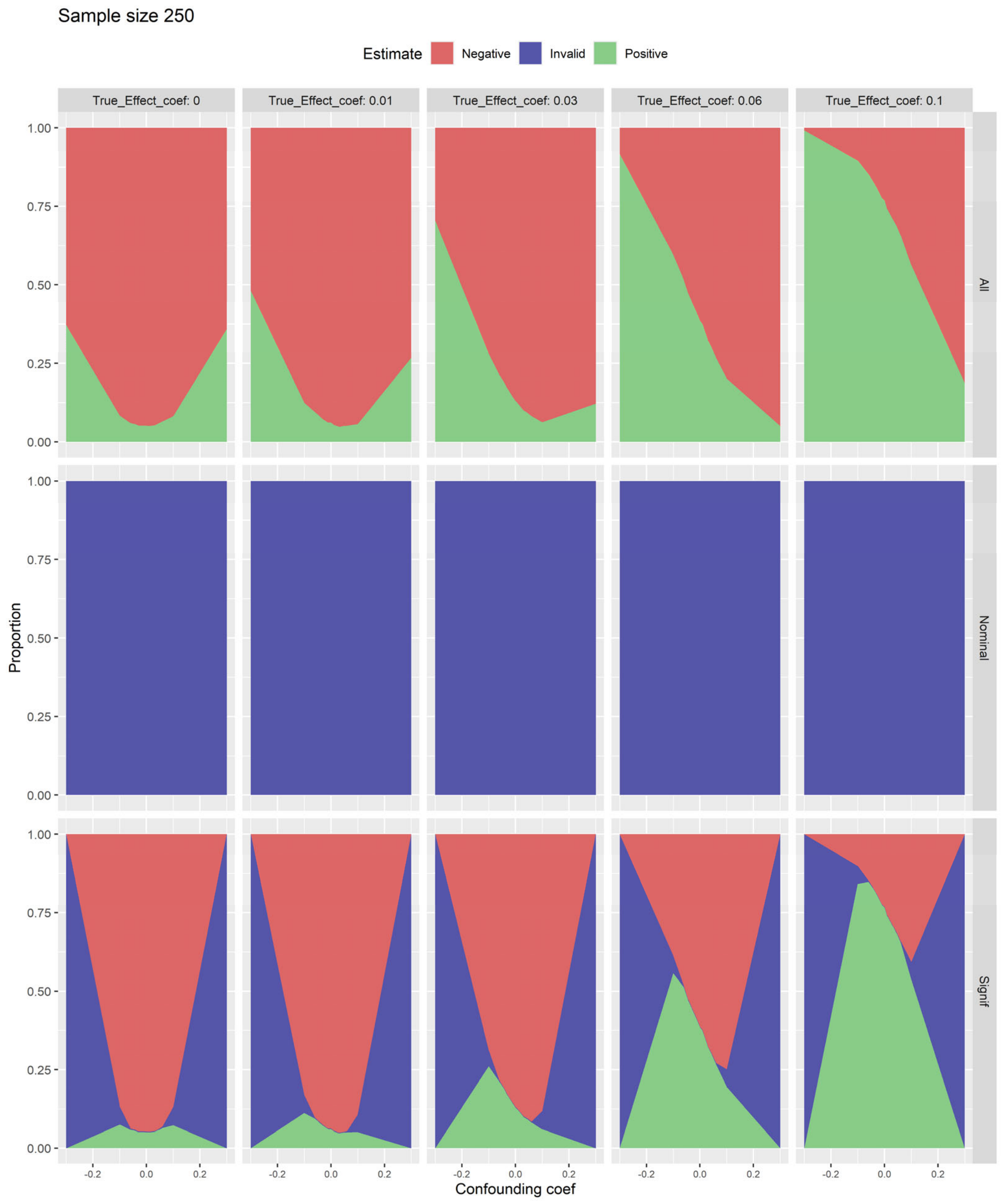

**S19. Three-state performance at the database level on simulation with sample size 1000.** Graphs show the proportion of studies that were positive (green, null hypothesis rejected), invalid (blue, study rejected), and negative (red, null hypothesis not rejected) on a single-database study with sample size 1000. The proportion is plotted against confounding coefficient $c_t$ from $-0.3$ to 0.3, and graphs from left to right show different values for effect coefficient $c_e$ from 0 to 0.1. The rows represent the three types of rules applied only to a single database under study: **All** ignores imbalance, **Nominal** tests for any covariate's standardized mean difference reaching or exceeding 0.1, and **Signif** tests for any covariate's standardized mean difference statistically significantly reaching or exceeding 0.1.

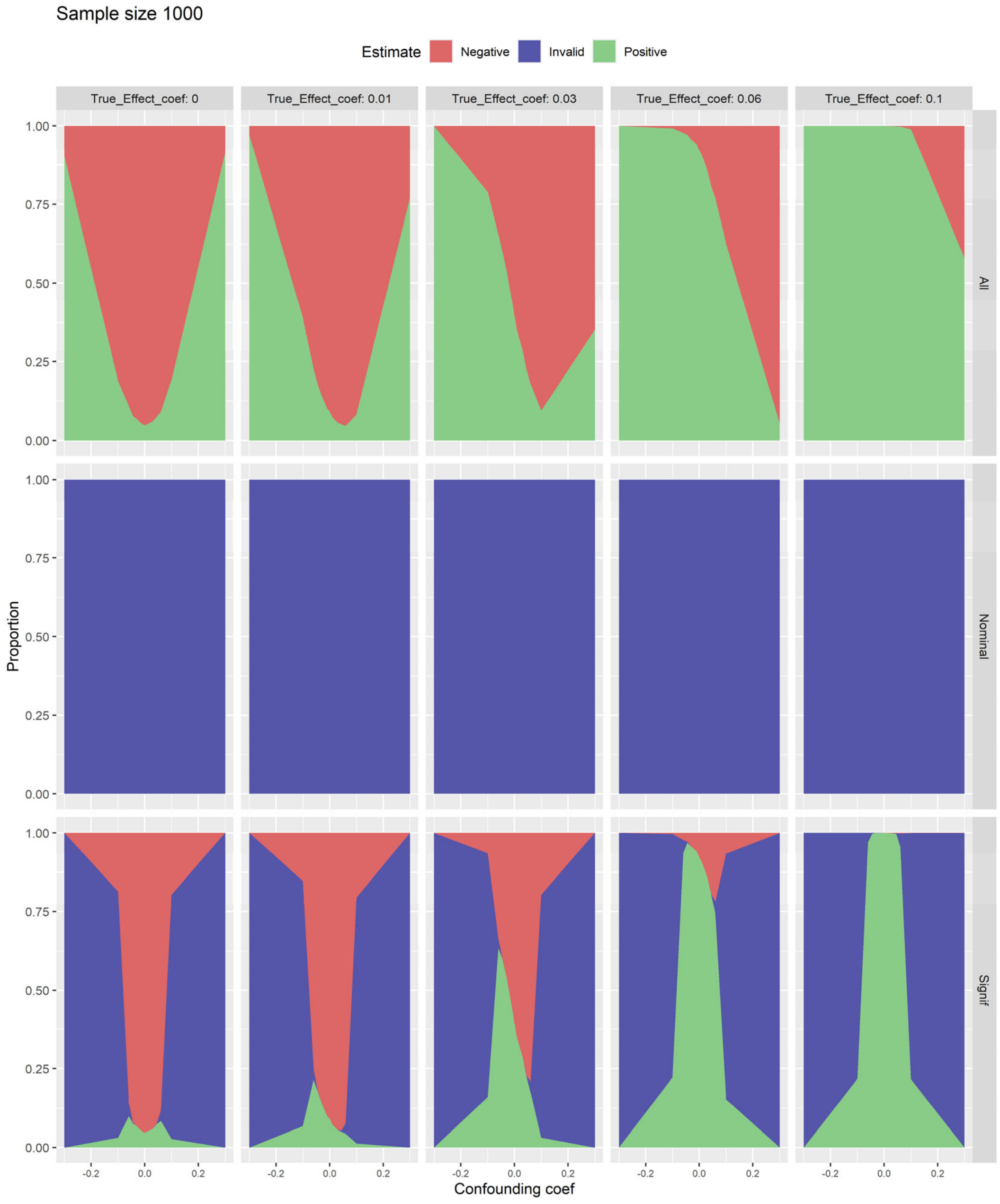

**S20. Three-state performance at the database level on simulation with sample size 2000.** Graphs show the proportion of studies that were positive (green, null hypothesis rejected), invalid (blue, study rejected), and negative (red, null hypothesis not rejected) on a single-database study with sample size 2000. The proportion is plotted against confounding coefficient $c_t$ from −0.3 to 0.3, and graphs from left to right show different values for effect coefficient $c_e$ from 0 to 0.1. The rows represent the three types of rules applied only to a single database under study: **All** ignores imbalance, **Nominal** tests for any covariate's standardized mean difference reaching or exceeding 0.1, and **Signif** tests for any covariate's standardized mean difference statistically significantly reaching or exceeding 0.1.

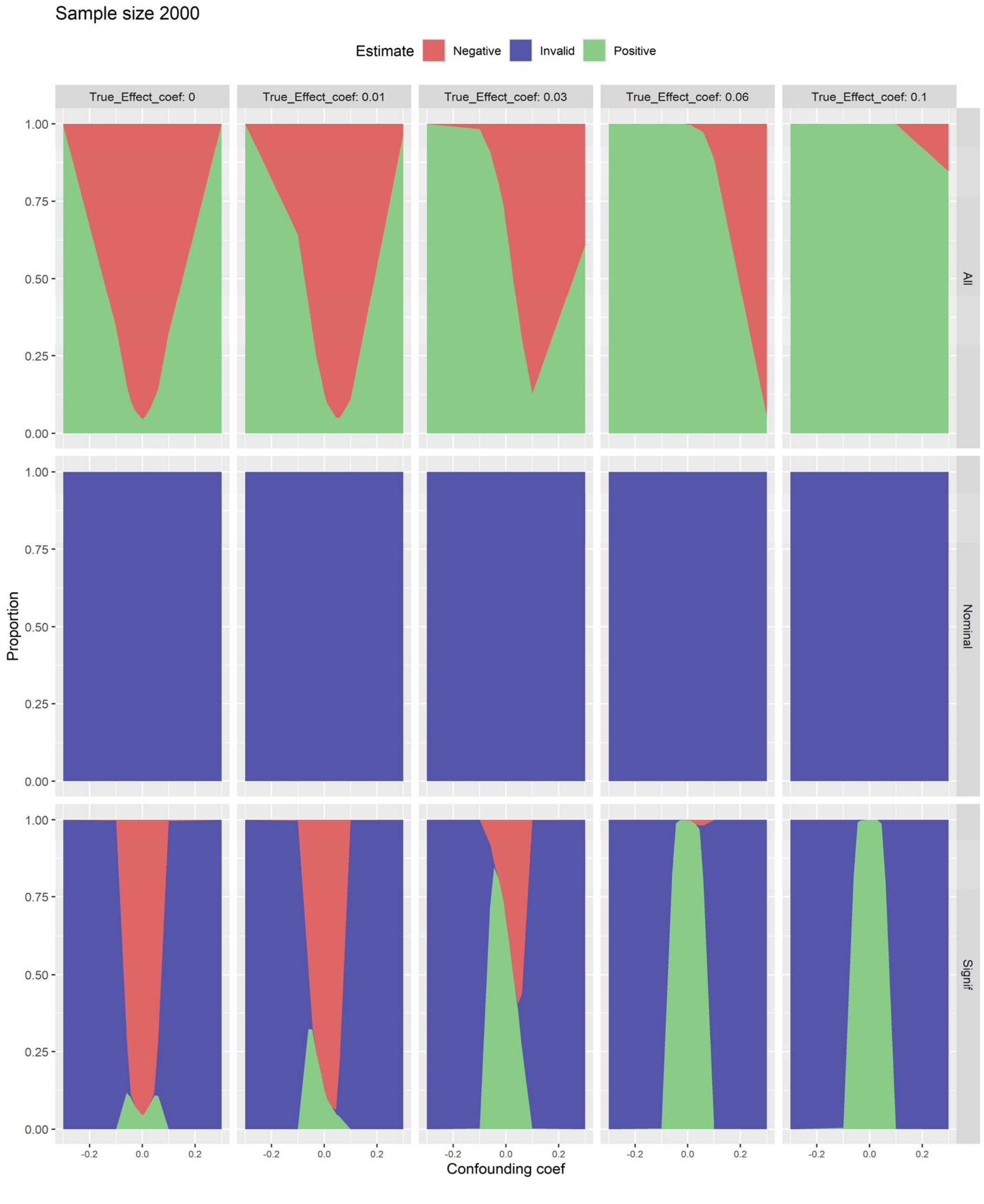

**S21. Three-state performance at the database level on simulation with sample size 4000.** Graphs show the proportion of studies that were positive (green, null hypothesis rejected), invalid (blue, study rejected), and negative (red, null hypothesis not rejected) on a single-database study with sample size 4000. The proportion is plotted against confounding coefficient $c_t$ from –0.3 to 0.3, and graphs from left to right show different values for effect coefficient $c_e$ from 0 to 0.1. The rows represent the three types of rules applied only to a single database under study: **All** ignores imbalance, **Nominal** tests for any covariate's standardized mean difference reaching or exceeding 0.1, and **Signif** tests for any covariate's standardized mean difference statistically significantly reaching or exceeding 0.1.

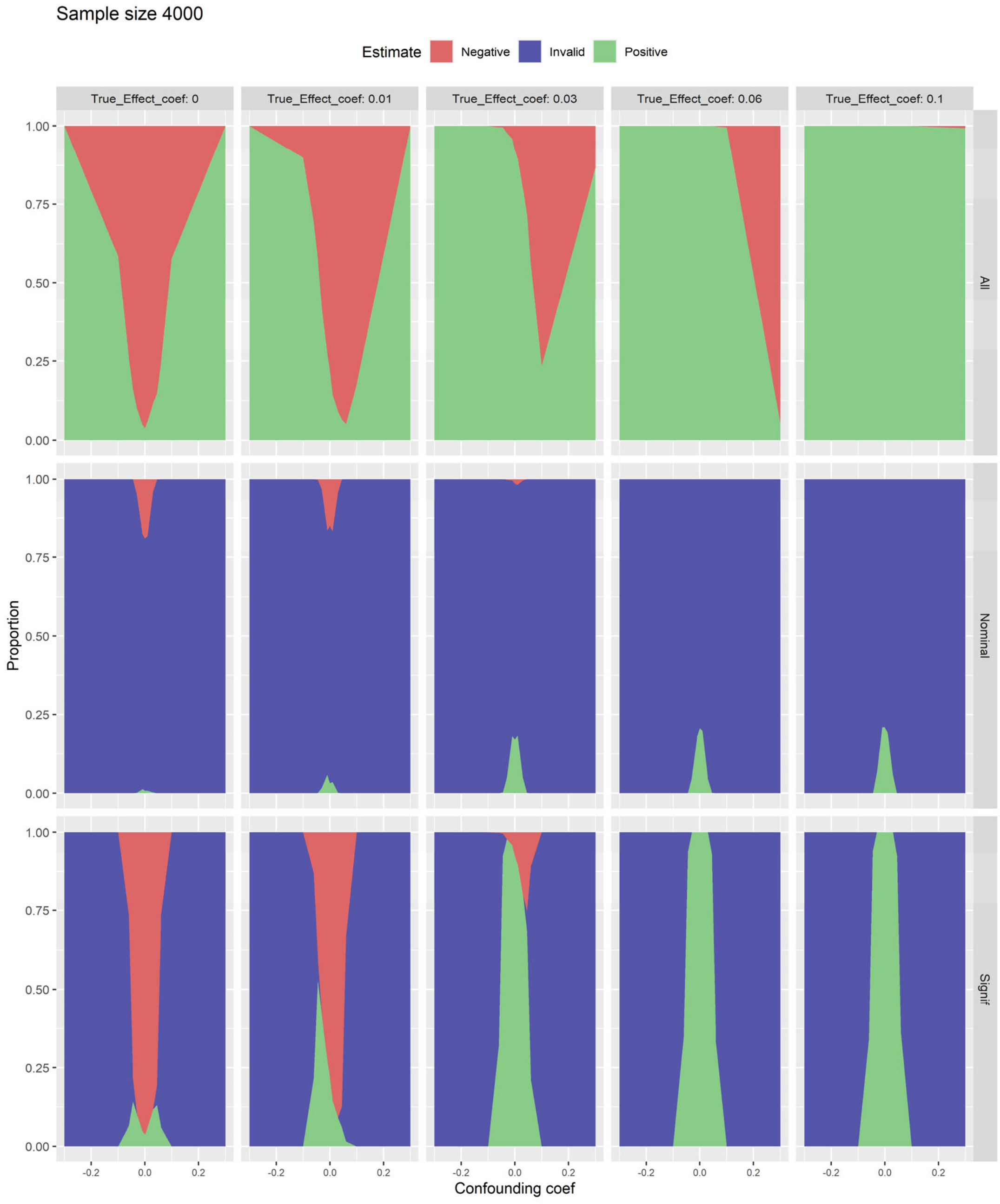

**S22. Three-state rule performance at the network level on simulation with sample size 250.** Graphs show the proportion of studies that were positive (green, null hypothesis rejected), invalid (blue, study rejected), and negative (red, null hypothesis not rejected) with sample size 250. The proportion is plotted against confounding coefficient $c_t$ from $-0.3$ to 0.3, and graphs from left to right show different values for effect coefficient $c_e$ from 0 to 0.1. The nine rows represent the nine rules listed in the text.

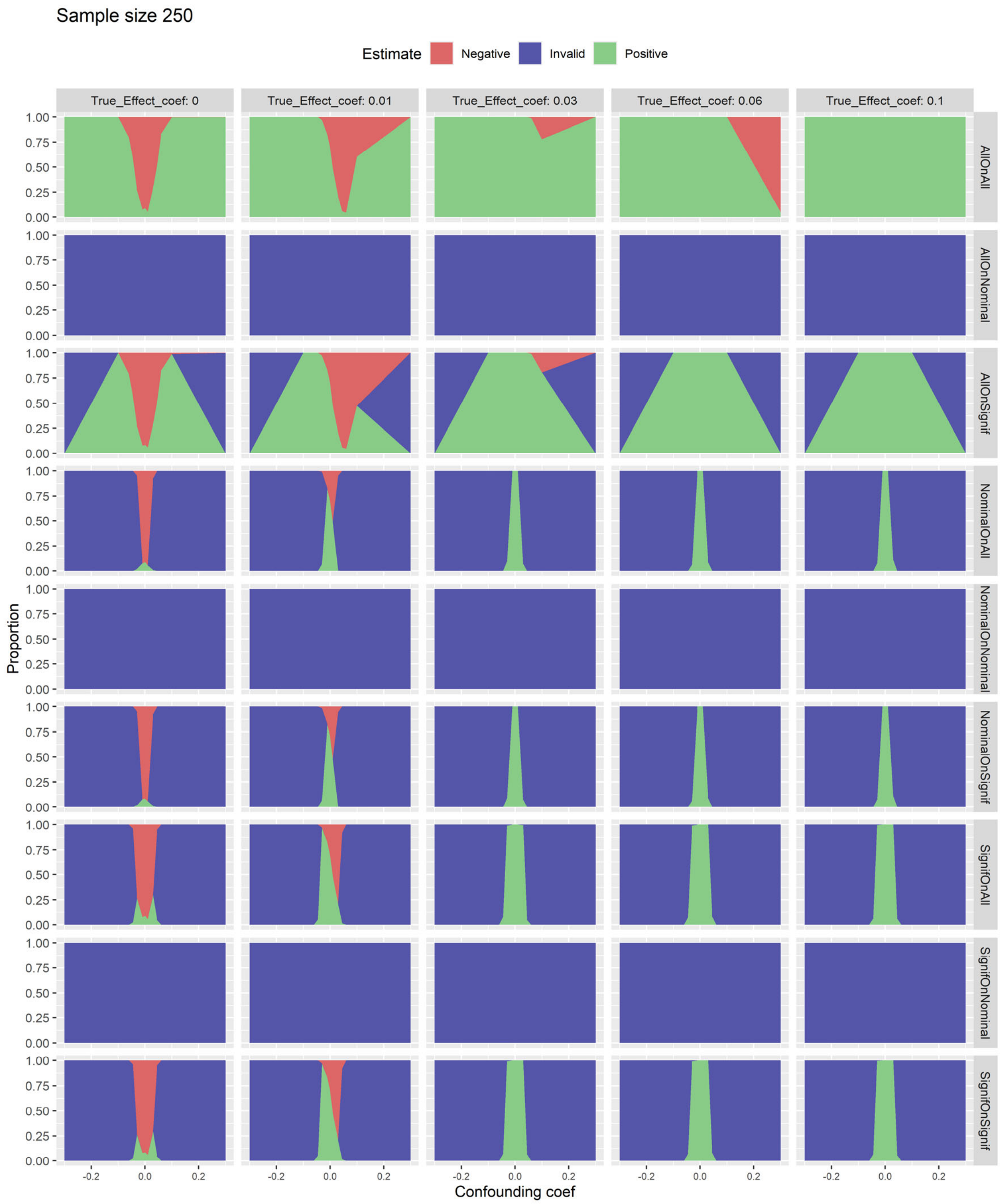

**S23. Three-state rule performance at the network level on simulation with sample size 1000.** Graphs show the proportion of studies that were positive (green, null hypothesis rejected), invalid (blue, study rejected), and negative (red, null hypothesis not rejected) with sample size 1000. The proportion is plotted against confounding coefficient $c_t$ from –0.3 to 0.3, and graphs from left to right show different values for effect coefficient $c_e$ from 0 to 0.1. The nine rows represent the nine rules listed in the text.

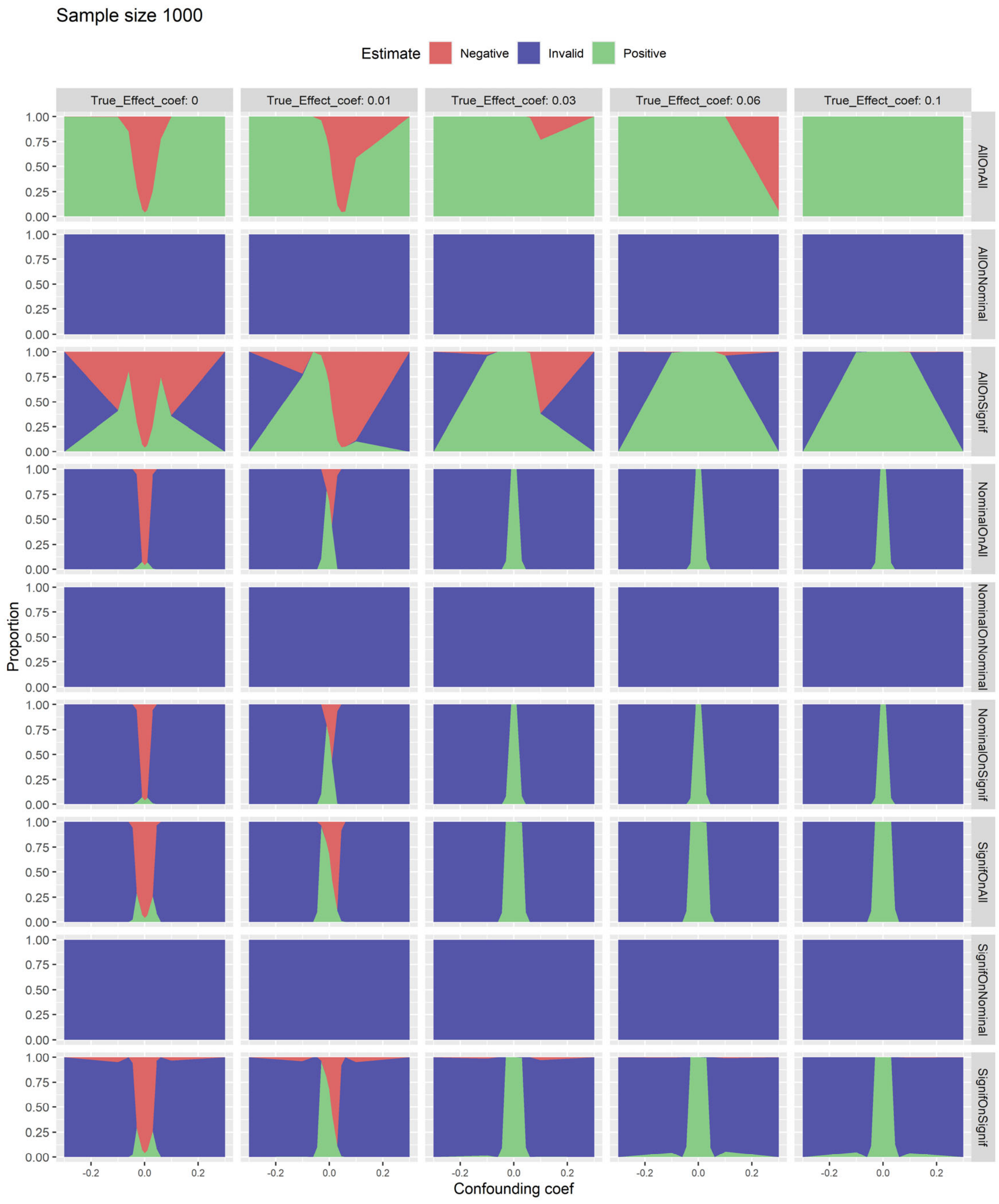

**S24. Three-state rule performance at the network level on simulation with sample size 2000.** Graphs show the proportion of studies that were positive (green, null hypothesis rejected), invalid (blue, study rejected), and negative (red, null hypothesis not rejected) with sample size 2000. The proportion is plotted against confounding coefficient $c_t$ from −0.3 to 0.3, and graphs from left to right show different values for effect coefficient $c_e$ from 0 to 0.1. The nine rows represent the nine rules listed in the text.

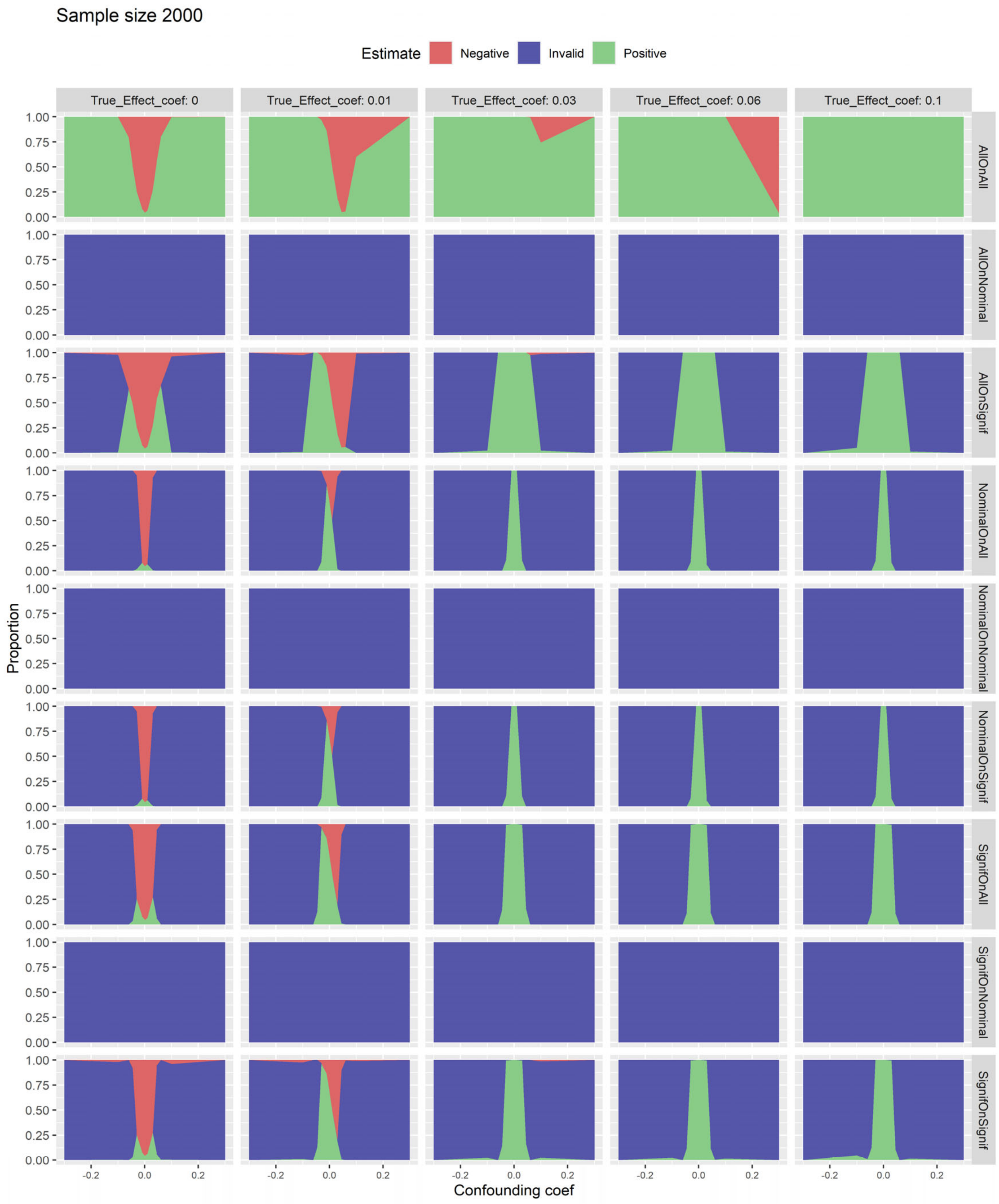

**S25. Three-state rule performance at the network level on simulation with sample size 4000.** Graphs show the proportion of studies that were positive (green, null hypothesis rejected), invalid (blue, study rejected), and negative (red, null hypothesis not rejected) with sample size 4000. The proportion is plotted against confounding coefficient $c_t$ from –0.3 to 0.3, and graphs from left to right show different values for effect coefficient $c_e$ from 0 to 0.1. The nine rows represent the nine rules listed in the text.

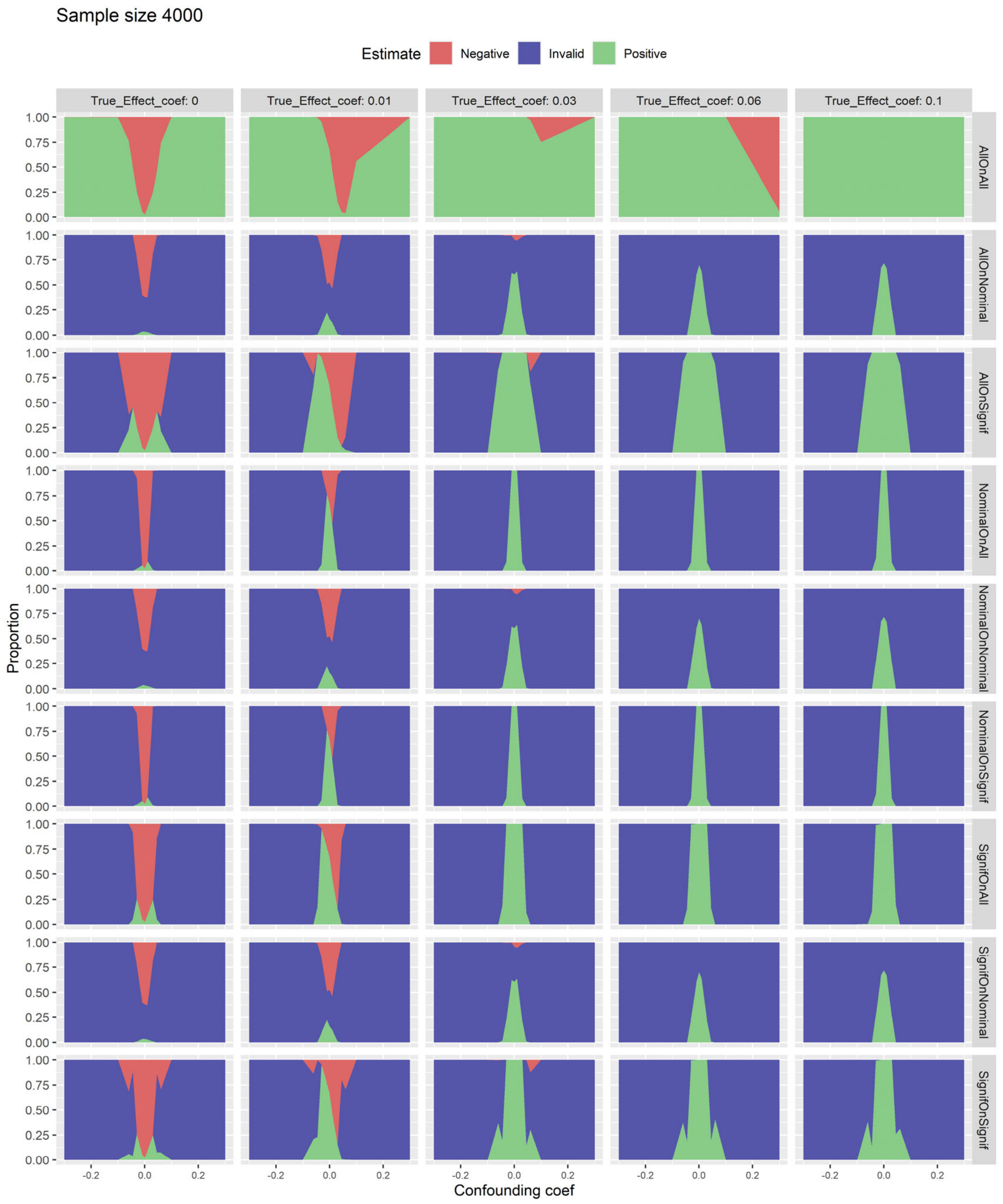

**S26. Three-state rule performance at the network level on simulation when number of databases is limited to 5 with sample size 250.** Graphs show the proportion of studies that were positive (green, null hypothesis rejected), invalid (blue, study rejected), and negative (red, null hypothesis not rejected) with sample size 250. The proportion is plotted against confounding coefficient $c_t$ from −0.3 to 0.3, and graphs from left to right show different values for effect coefficient $c_e$ from 0 to 0.1. The nine rows represent the nine rules listed in the text.

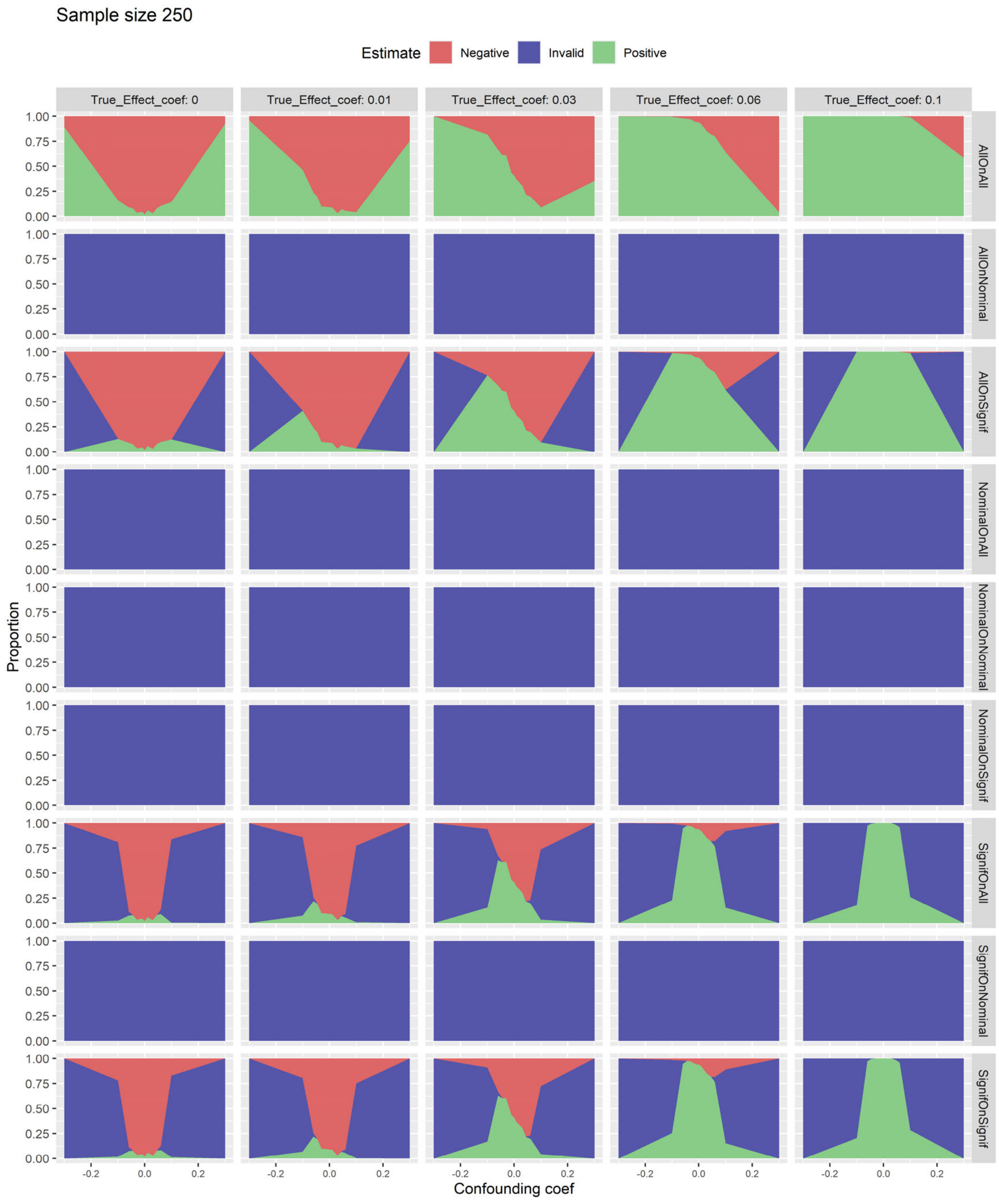

**S27. Three-state rule performance at the network level on simulation when number of databases is limited to 5 with sample size 500.** Graphs show the proportion of studies that were positive (green, null hypothesis rejected), invalid (blue, study rejected), and negative (red, null hypothesis not rejected) with sample size 500. The proportion is plotted against confounding coefficient $c_t$ from $-0.3$ to 0.3, and graphs from left to right show different values for effect coefficient $c_e$ from 0 to 0.1. The nine rows represent the nine rules listed in the text.

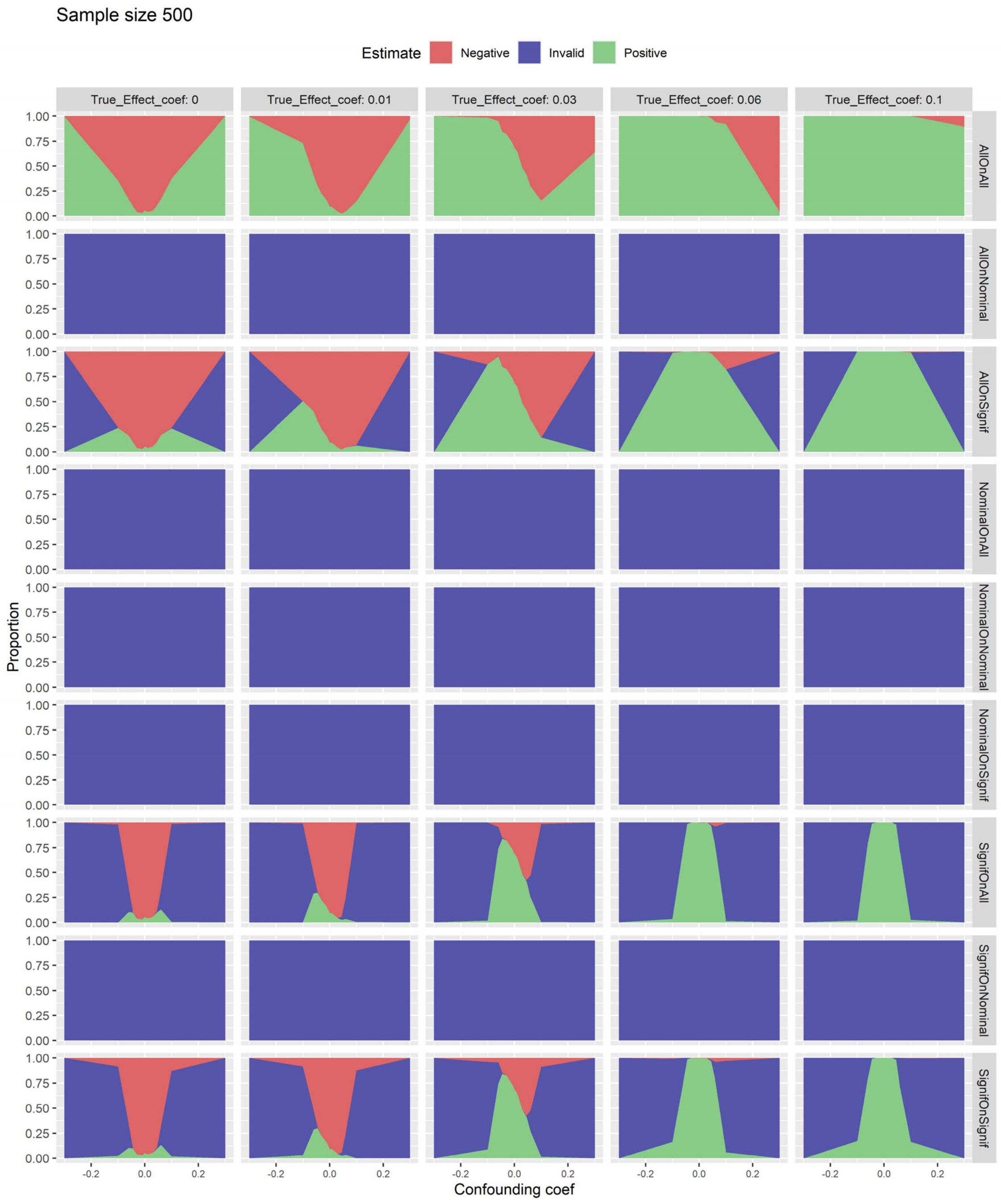

**S28. Three-state rule performance at the network level on simulation when number of databases is limited to 5 with sample size 1000.** Graphs show the proportion of studies that were positive (green, null hypothesis rejected), invalid (blue, study rejected), and negative (red, null hypothesis not rejected) with sample size 1000. The proportion is plotted against confounding coefficient $c_t$ from –0.3 to 0.3, and graphs from left to right show different values for effect coefficient $c_e$ from 0 to 0.1. The nine rows represent the nine rules listed in the text.

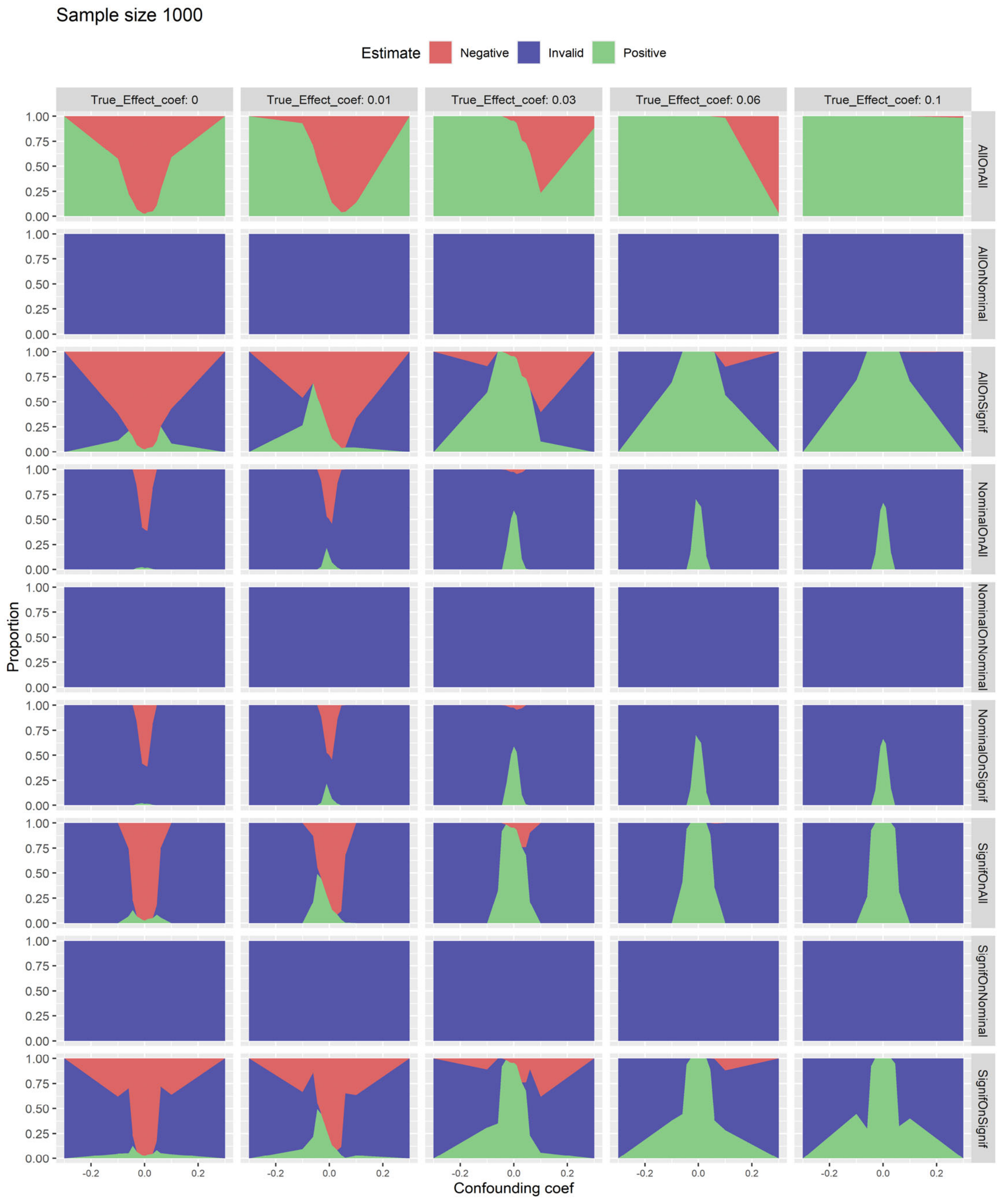

**S29. Three-state rule performance at the network level on simulation when number of databases is limited to 5 with sample size 2000.** Graphs show the proportion of studies that were positive (green, null hypothesis rejected), invalid (blue, study rejected), and negative (red, null hypothesis not rejected) with sample size 2000. The proportion is plotted against confounding coefficient $c_t$ from −0.3 to 0.3, and graphs from left to right show different values for effect coefficient $c_e$ from 0 to 0.1. The nine rows represent the nine rules listed in the text.

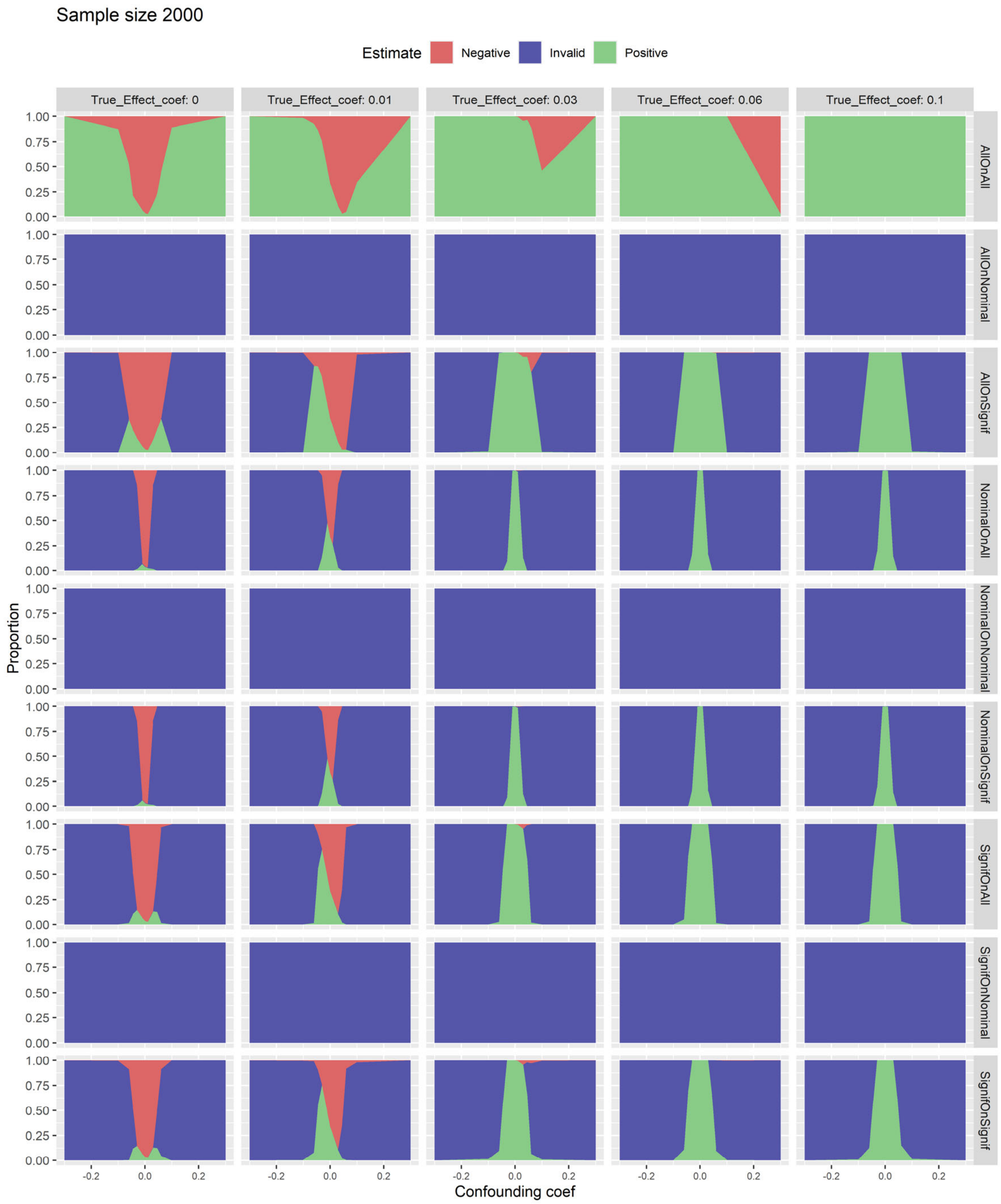

**S30. Three-state rule performance at the network level on simulation when number of databases is limited to 5 with sample size 4000.** Graphs show the proportion of studies that were positive (green, null hypothesis rejected), invalid (blue, study rejected), and negative (red, null hypothesis not rejected) with sample size 4000. The proportion is plotted against confounding coefficient $c_t$ from $-0.3$ to 0.3, and graphs from left to right show different values for effect coefficient $c_e$ from 0 to 0.1. The nine rows represent the nine rules listed in the text.

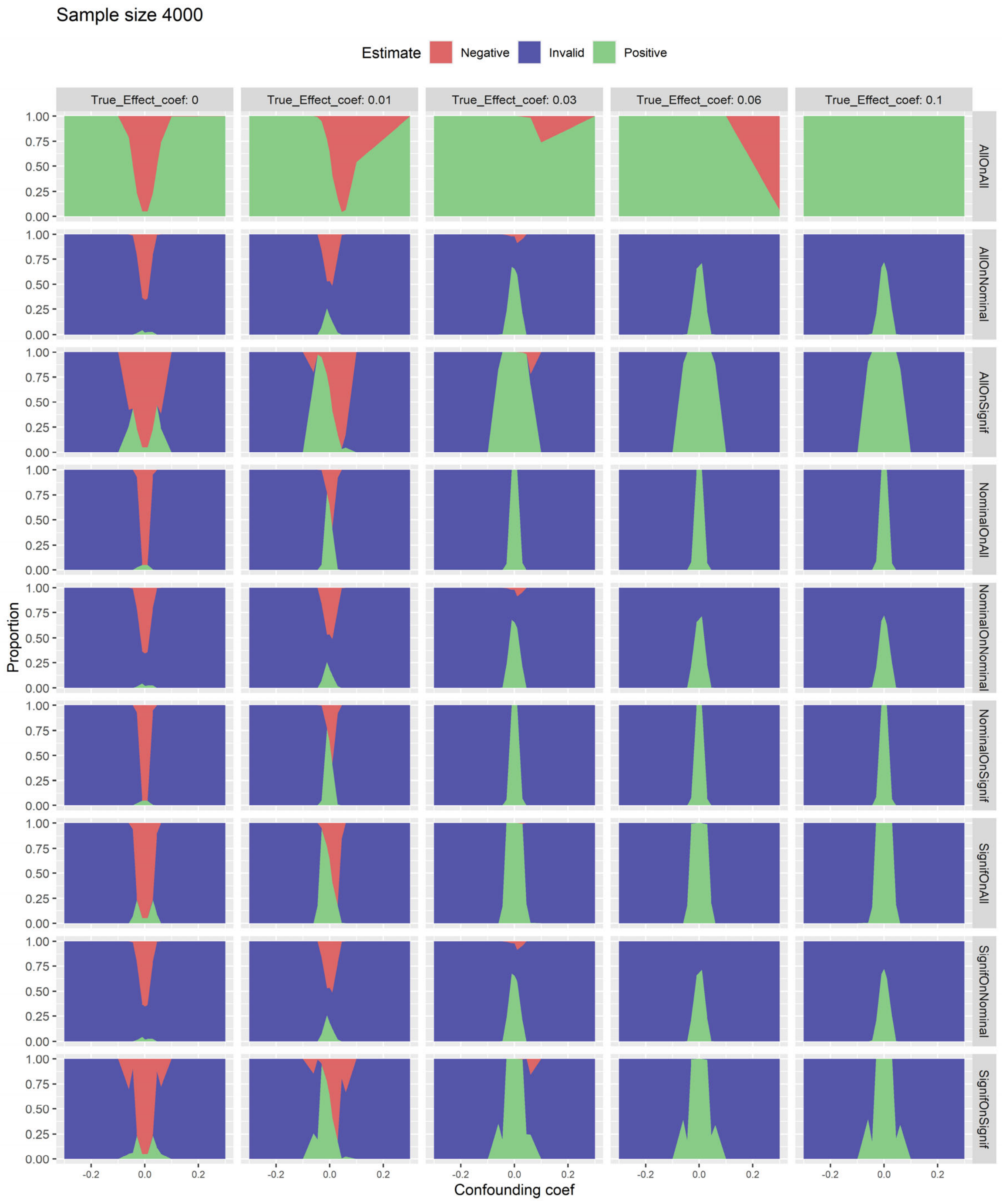

**S31. Three-state rule performance at the network level on simulation when outcome prevalence is low (1%) with sample size 250.** Graphs show the proportion of studies that were positive (green, null hypothesis rejected), invalid (blue, study rejected), and negative (red, null hypothesis not rejected) with sample size 250. The proportion is plotted against confounding coefficient $c_t$ from $-0.3$ to 0.3, and graphs from left to right show different values for effect coefficient $c_e$ from 0 to 0.1. The nine rows represent the nine rules listed in the text.

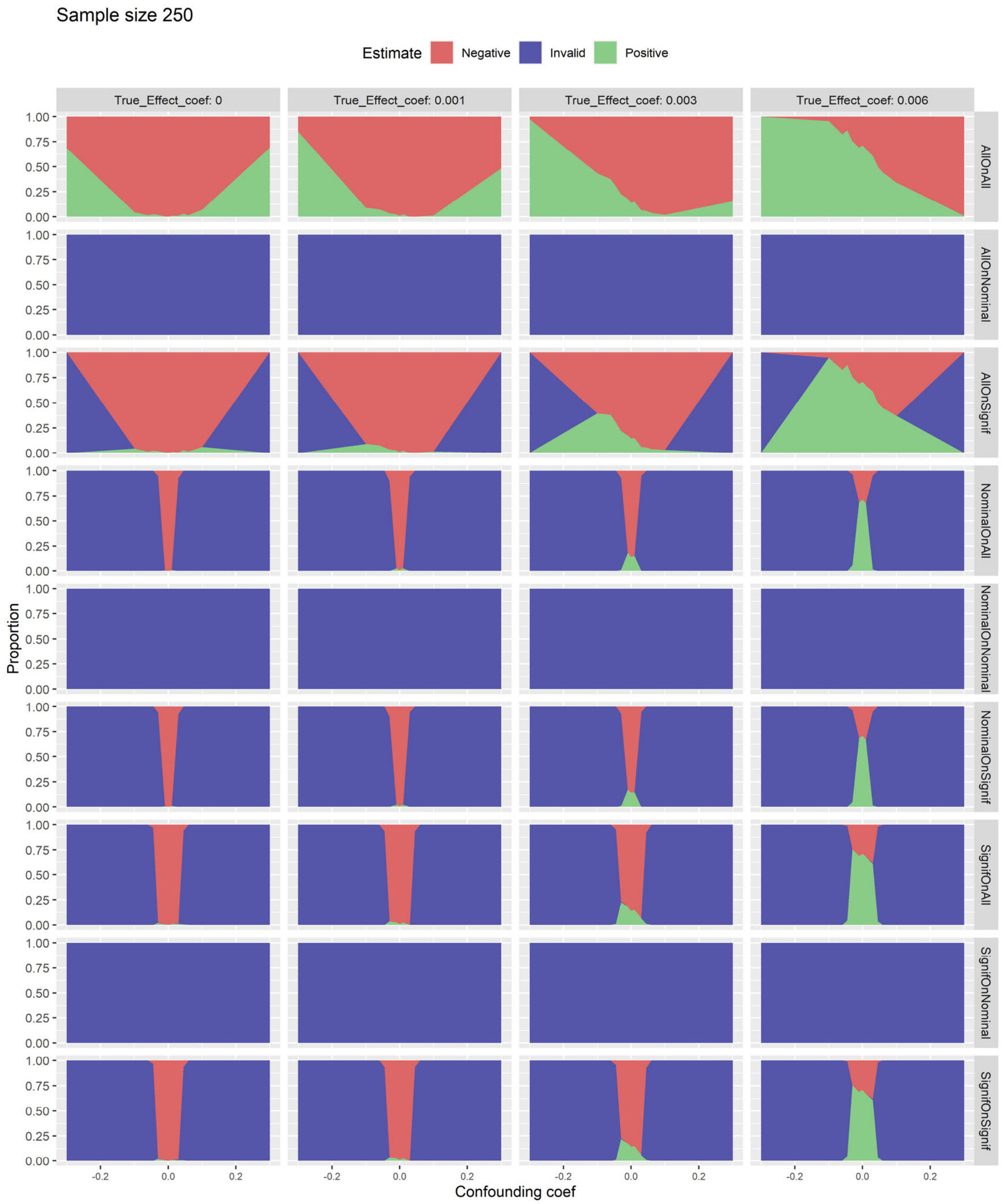

**S32. Three-state rule performance at the network level on simulation when outcome prevalence is low (1%) with sample size 500.** Graphs show the proportion of studies that were positive (green, null hypothesis rejected), invalid (blue, study rejected), and negative (red, null hypothesis not rejected) with sample size 500. The proportion is plotted against confounding coefficient $c_t$ from $-0.3$ to 0.3, and graphs from left to right show different values for effect coefficient $c_e$ from 0 to 0.1. The nine rows represent the nine rules listed in the text.

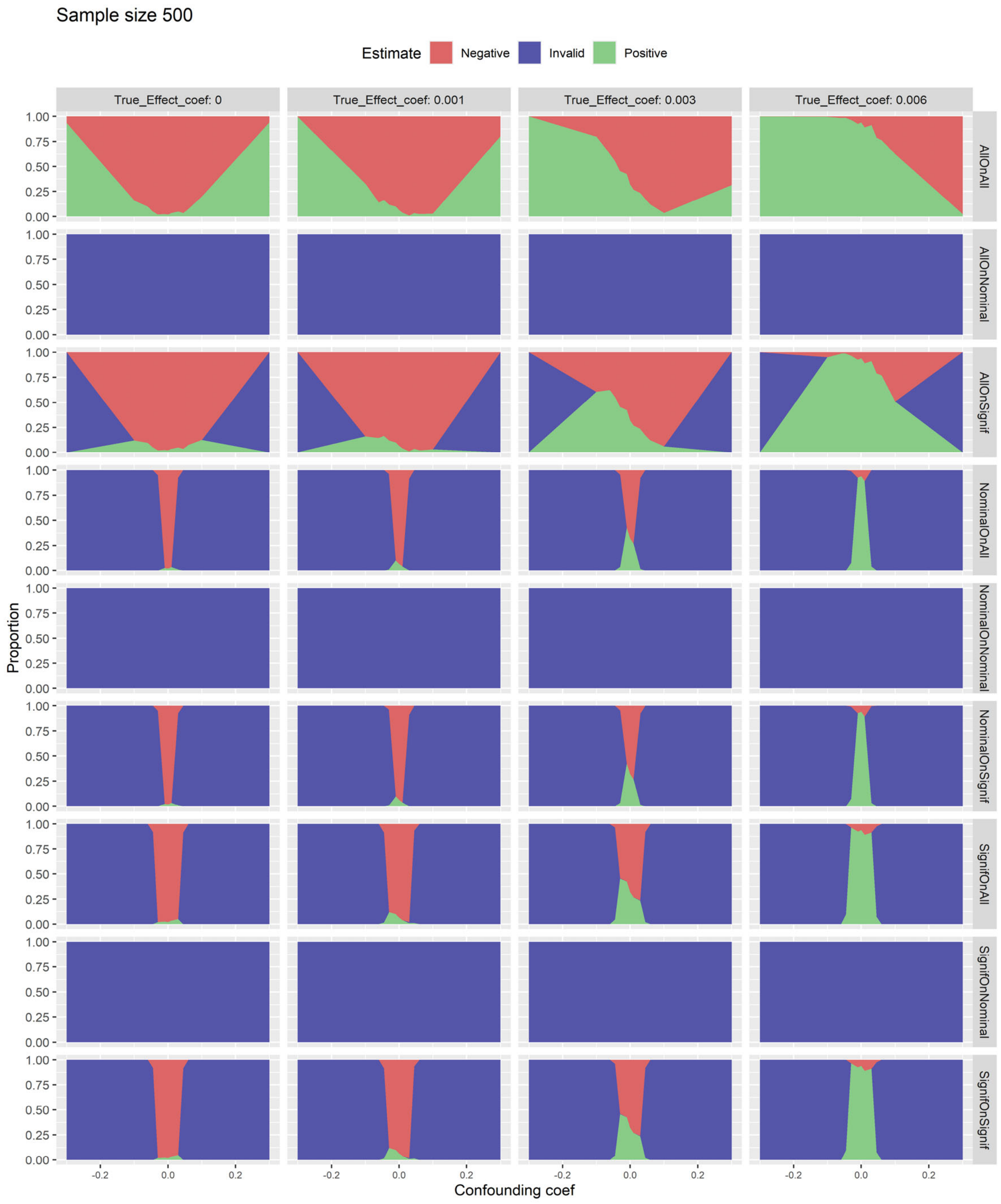

**S33. Three-state rule performance at the network level on simulation when outcome prevalence is low (1%) with sample size 1000.** Graphs show the proportion of studies that were positive (green, null hypothesis rejected), invalid (blue, study rejected), and negative (red, null hypothesis not rejected) with sample size 1000. The proportion is plotted against confounding coefficient $c_t$ from −0.3 to 0.3, and graphs from left to right show different values for effect coefficient $c_e$ from 0 to 0.1. The nine rows represent the nine rules listed in the text.

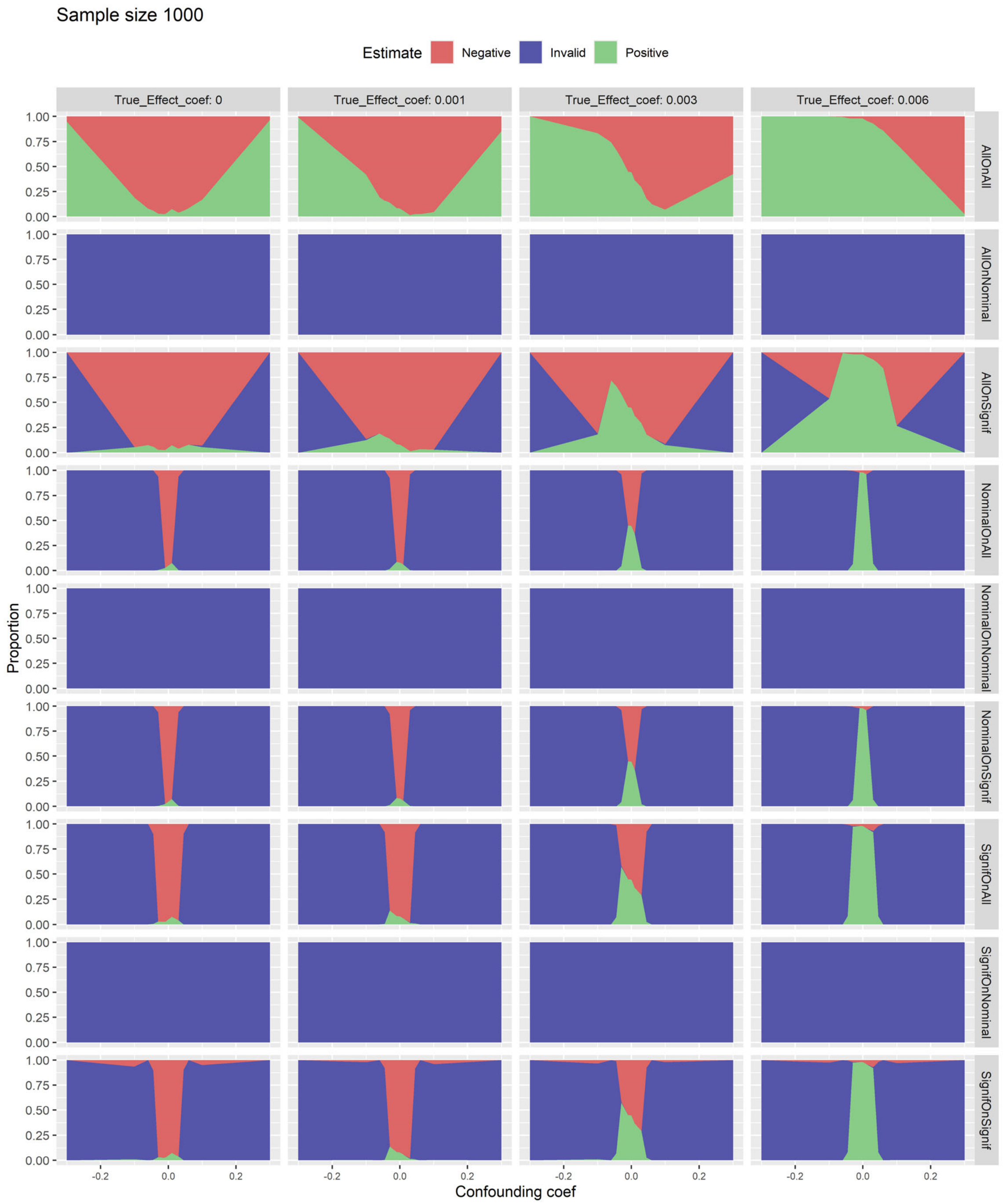

**S34. Three-state rule performance at the network level on simulation when outcome prevalence is low (1%) with sample size 2000.** Graphs show the proportion of studies that were positive (green, null hypothesis rejected), invalid (blue, study rejected), and negative (red, null hypothesis not rejected) with sample size 2000. The proportion is plotted against confounding coefficient $c_t$ from –0.3 to 0.3, and graphs from left to right show different values for effect coefficient $c_e$ from 0 to 0.1. The nine rows represent the nine rules listed in the text.

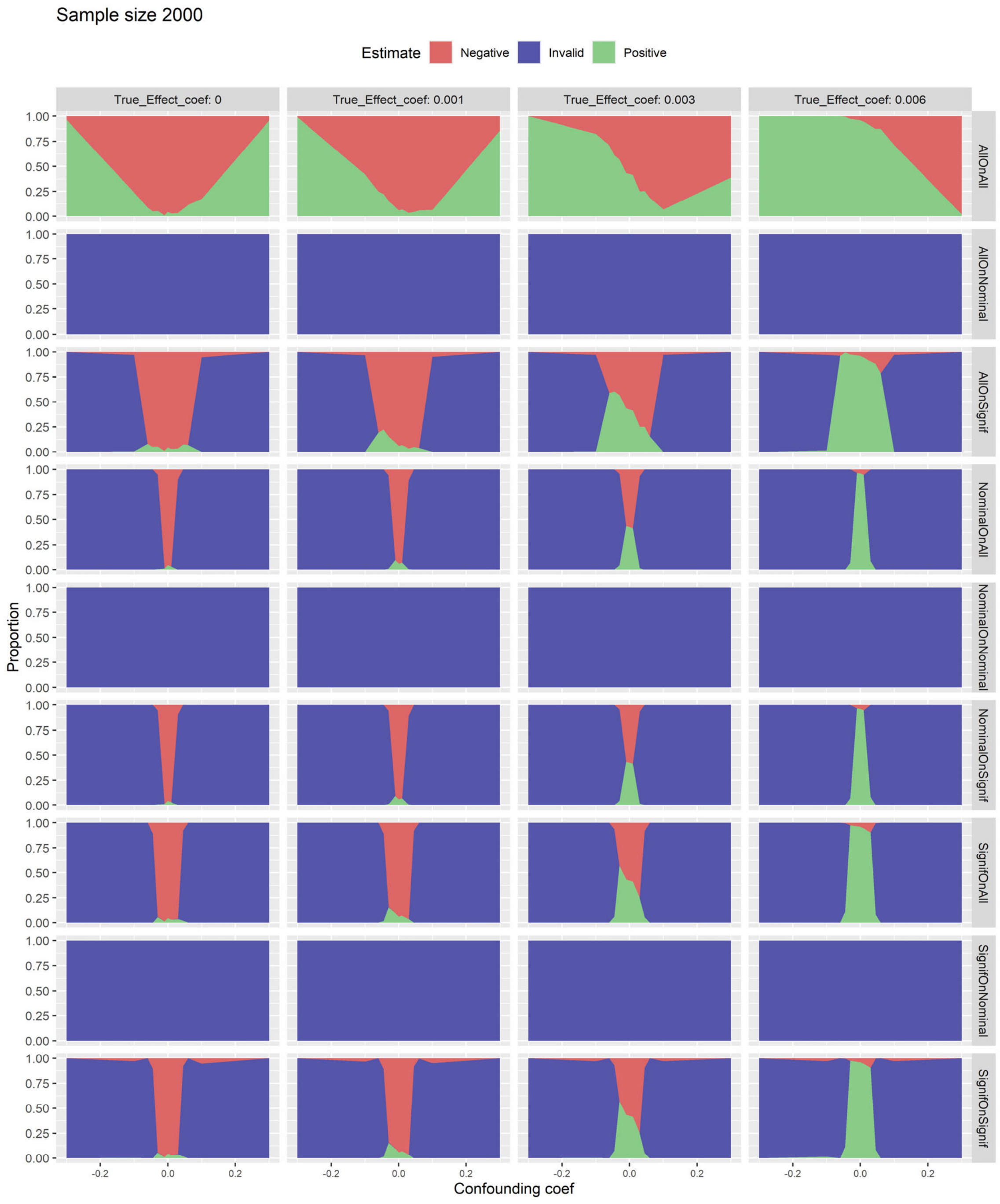

**S35. Three-state rule performance at the network level on simulation when outcome prevalence is low (1%) with sample size 4000.** Graphs show the proportion of studies that were positive (green, null hypothesis rejected), invalid (blue, study rejected), and negative (red, null hypothesis not rejected) with sample size 4000. The proportion is plotted against confounding coefficient $c_t$ from –0.3 to 0.3, and graphs from left to right show different values for effect coefficient $c_e$ from 0 to 0.1. The nine rows represent the nine rules listed in the text.

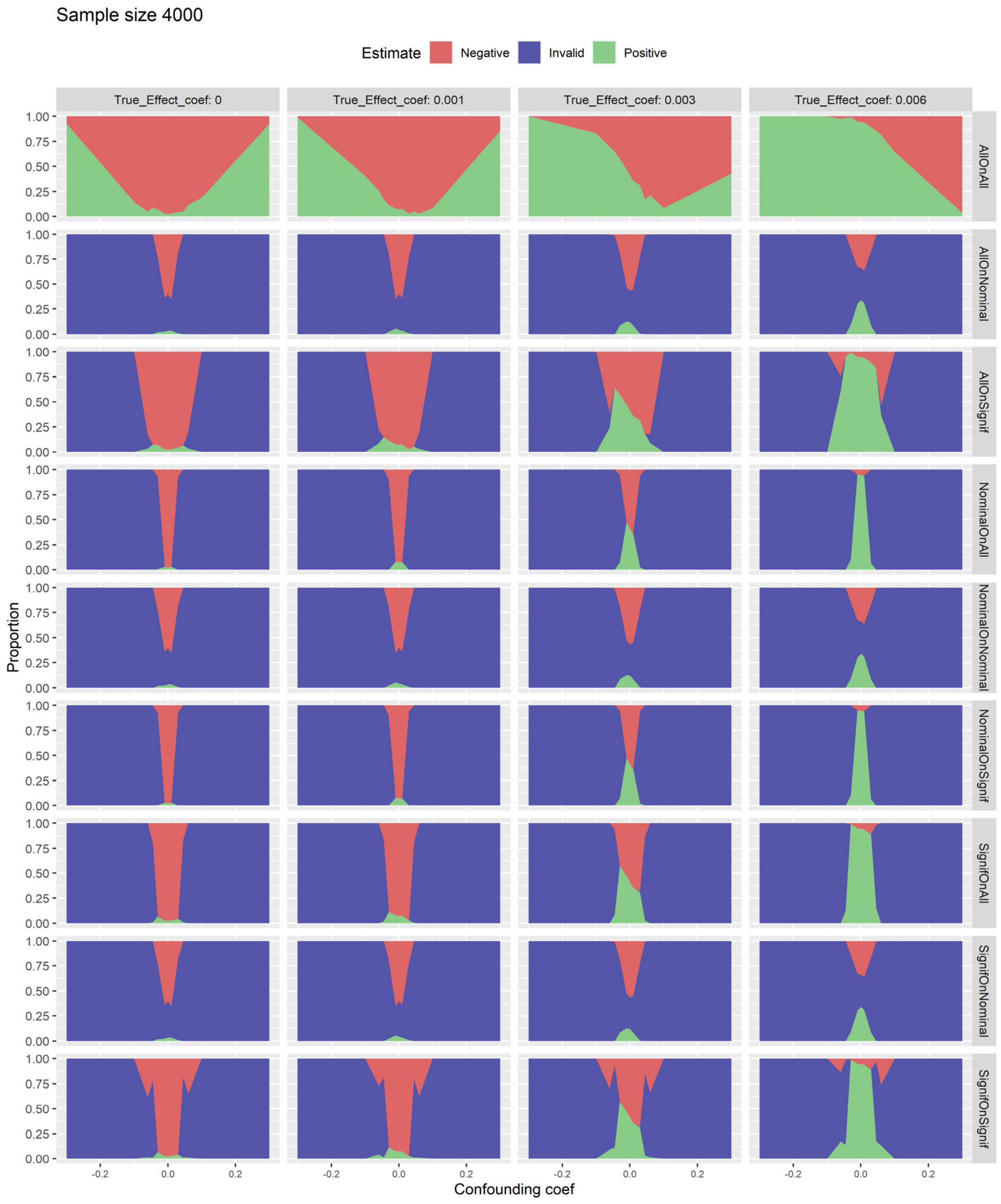

**S36. Three-state rule performance at the network level on simulation when covariate prevalence is low (10%) with sample size 250.** Graphs show the proportion of studies that were positive (green, null hypothesis rejected), invalid (blue, study rejected), and negative (red, null hypothesis not rejected) with sample size 250. The proportion is plotted against confounding coefficient $c_t$ from −0.3 to 0.3, and graphs from left to right show different values for effect coefficient $c_e$ from 0 to 0.1. The nine rows represent the nine rules listed in the text.

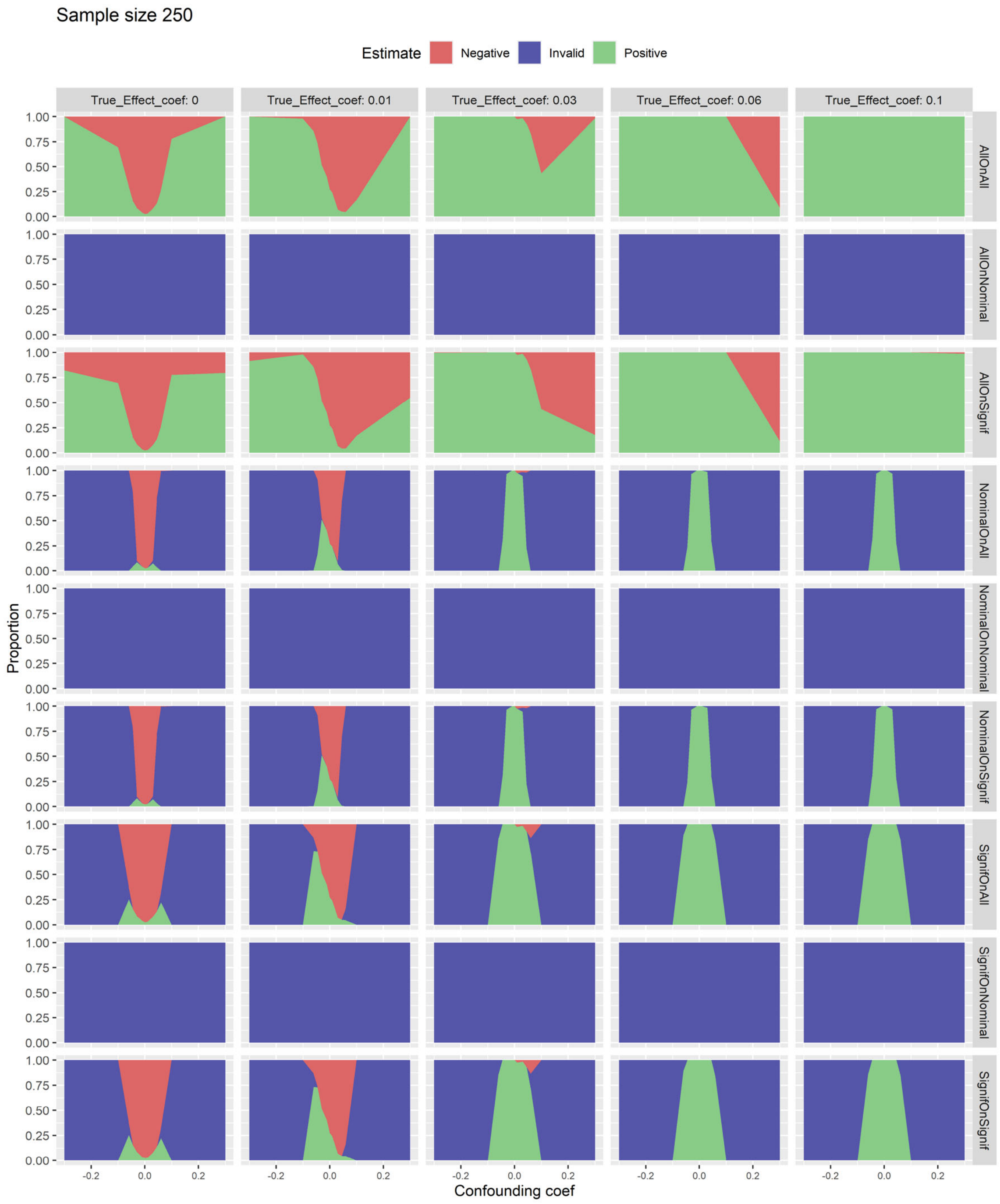

**S37. Three-state rule performance at the network level on simulation when covariate prevalence is low (10%) with sample size 500.** Graphs show the proportion of studies that were positive (green, null hypothesis rejected), invalid (blue, study rejected), and negative (red, null hypothesis not rejected) with sample size 500. The proportion is plotted against confounding coefficient $c_t$ from –0.3 to 0.3, and graphs from left to right show different values for effect coefficient $c_e$ from 0 to 0.1. The nine rows represent the nine rules listed in the text.

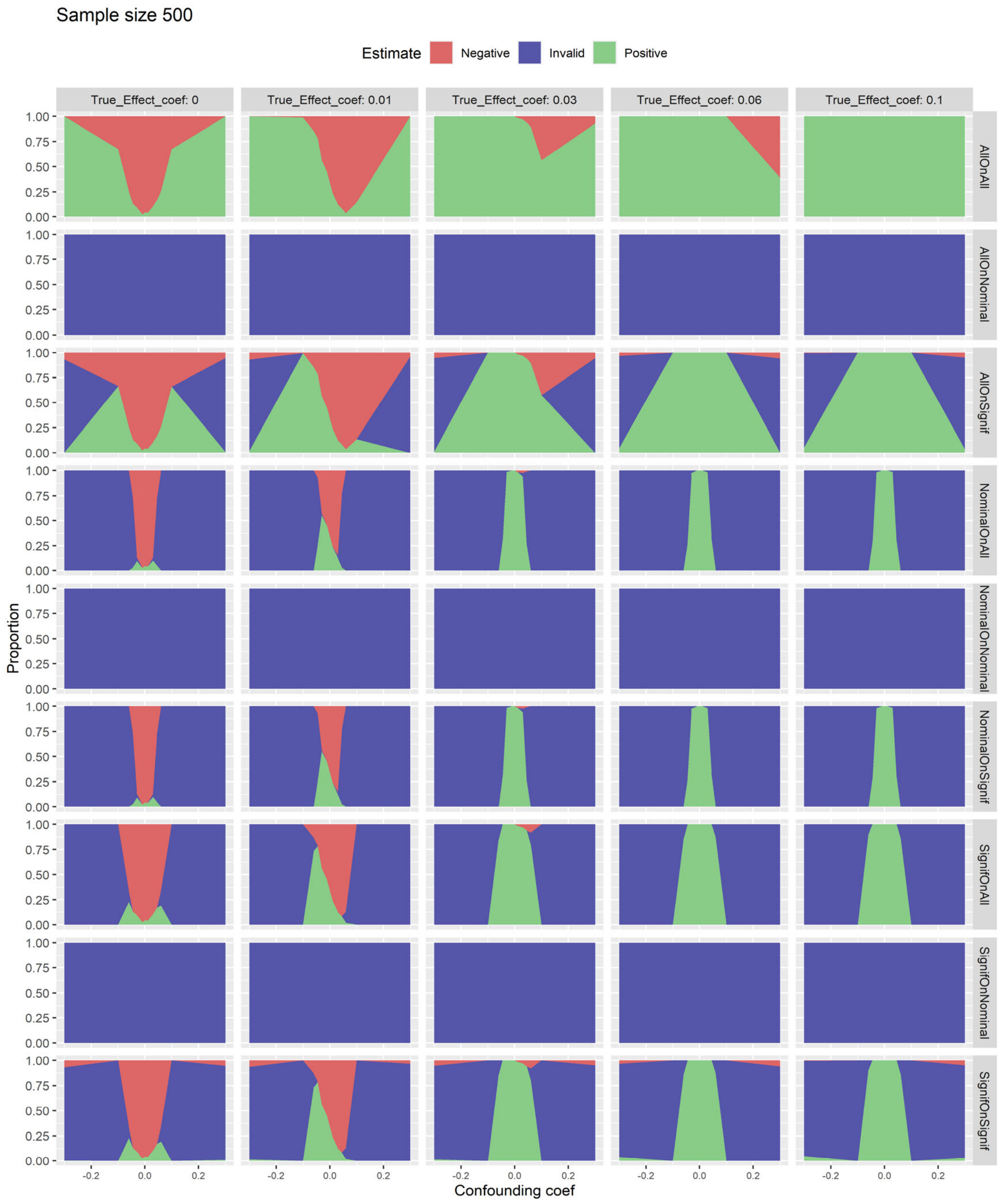

**S38. Three-state rule performance at the network level on simulation when covariate prevalence is low (10%) with sample size 1000.** Graphs show the proportion of studies that were positive (green, null hypothesis rejected), invalid (blue, study rejected), and negative (red, null hypothesis not rejected) with sample size 1000. The proportion is plotted against confounding coefficient $c_t$ from −0.3 to 0.3, and graphs from left to right show different values for effect coefficient $c_e$ from 0 to 0.1. The nine rows represent the nine rules listed in the text.

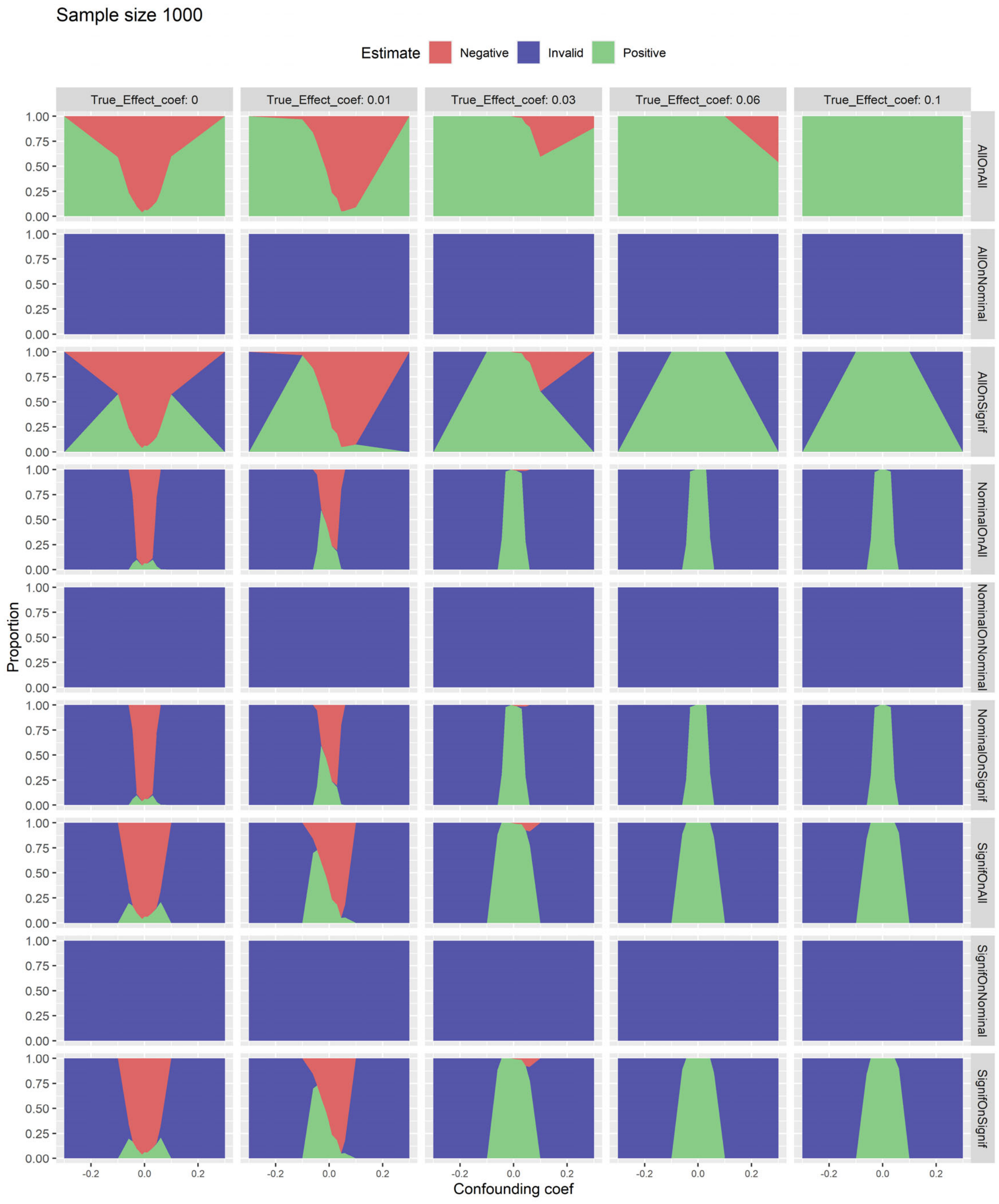

**S39. Three-state rule performance at the network level on simulation when covariate prevalence is low (10%) with sample size 2000.** Graphs show the proportion of studies that were positive (green, null hypothesis rejected), invalid (blue, study rejected), and negative (red, null hypothesis not rejected) with sample size 2000. The proportion is plotted against confounding coefficient $c_t$ from −0.3 to 0.3, and graphs from left to right show different values for effect coefficient $c_e$ from 0 to 0.1. The nine rows represent the nine rules listed in the text.

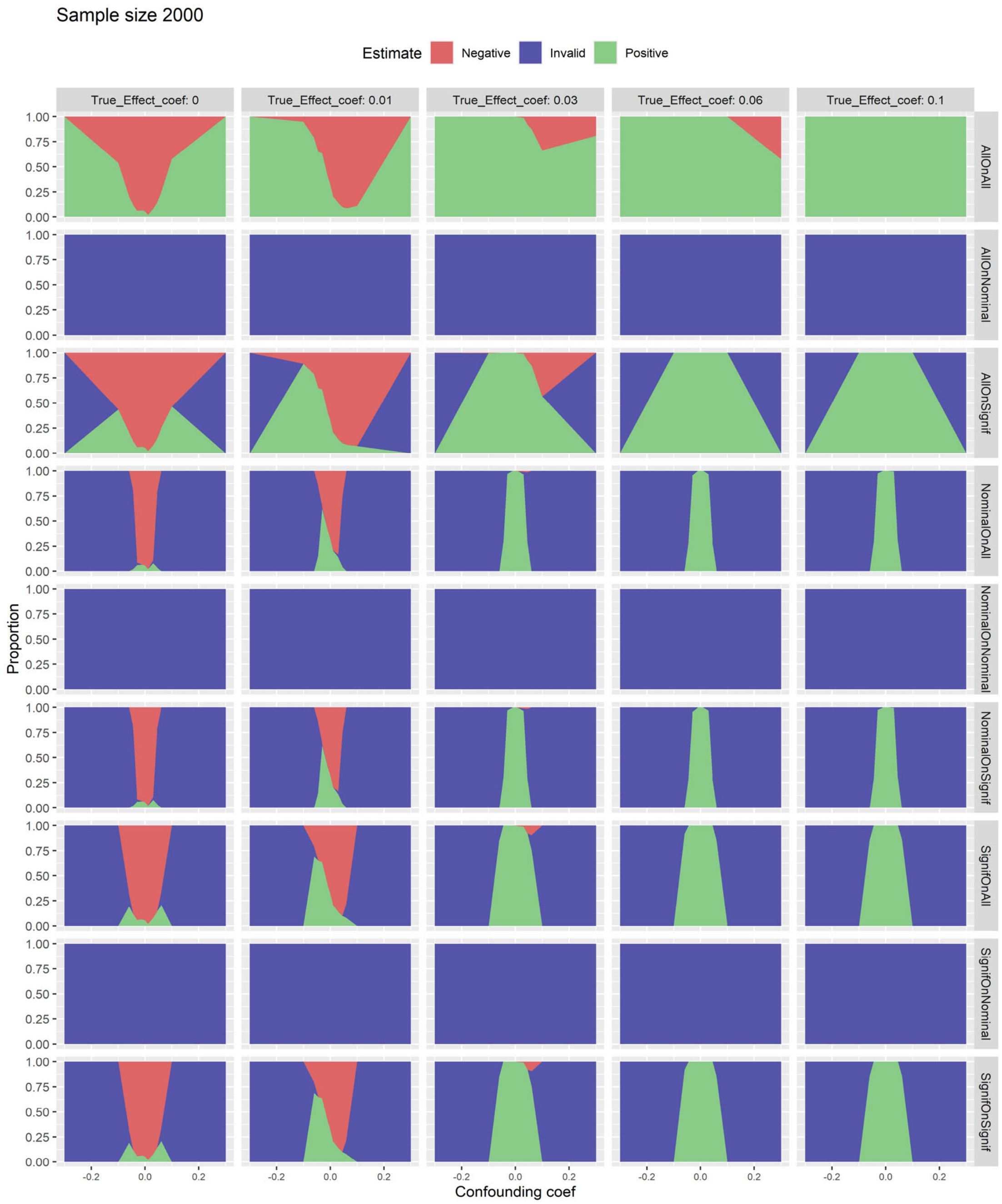

**S40. Three-state rule performance at the network level on simulation when covariate prevalence is low (10%) with sample size 4000.** Graphs show the proportion of studies that were positive (green, null hypothesis rejected), invalid (blue, study rejected), and negative (red, null hypothesis not rejected) with sample size 4000. The proportion is plotted against confounding coefficient $c_t$ from −0.3 to 0.3, and graphs from left to right show different values for effect coefficient $c_e$ from 0 to 0.1. The nine rows represent the nine rules listed in the text.

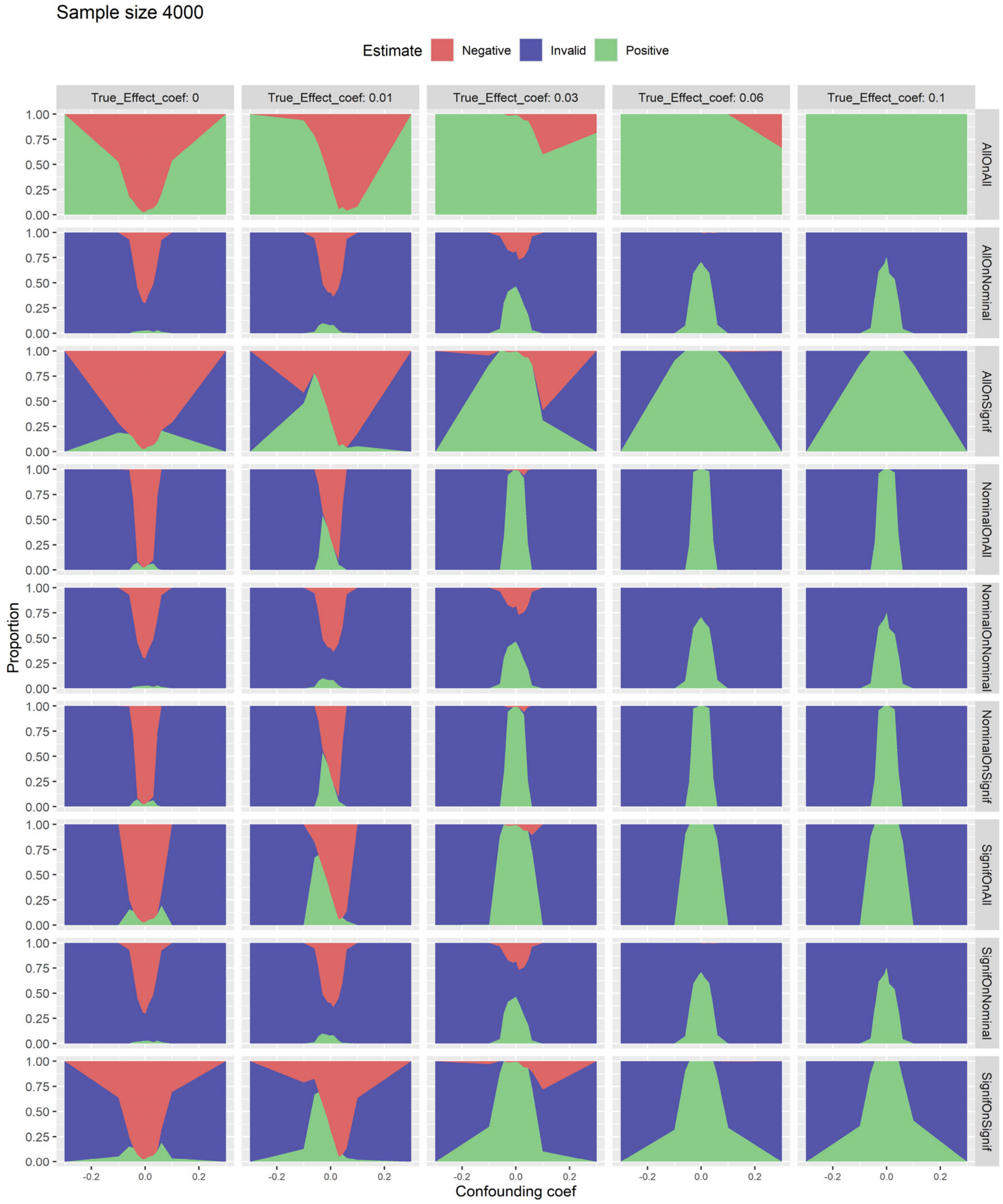

**S41. Three-state rule performance at the network level on simulation when confounding is heterogeneous ($c_t$ –0.3 to 0.3).** Graphs show the proportion of studies that were positive (green, null hypothesis rejected), invalid (blue, study rejected), and negative (red, null hypothesis not rejected). The proportion is plotted against sample size, and graphs from left to right show different values for effect coefficient $c_e$ from 0 to 0.1. The nine rows represent the nine rules listed in the text.

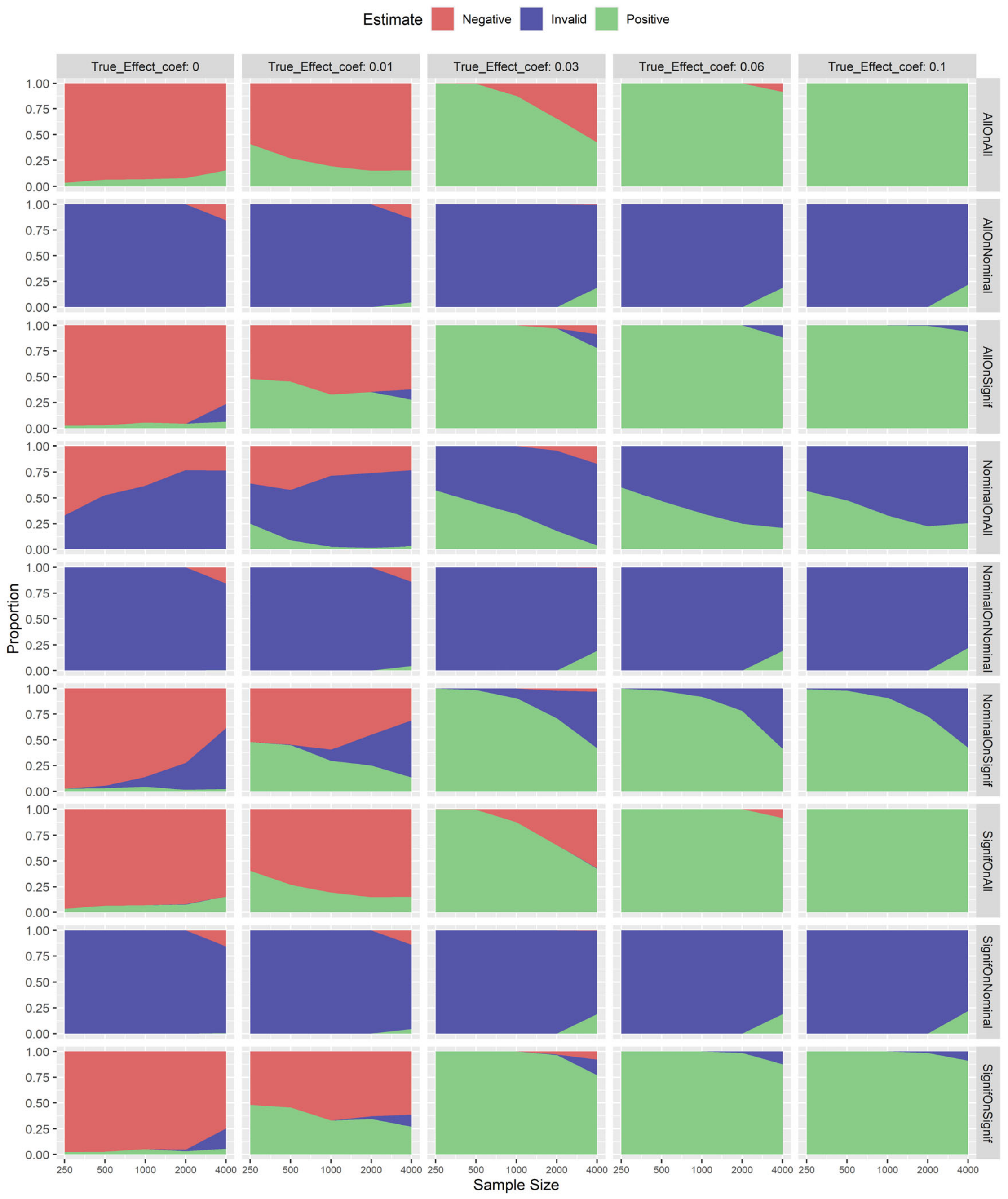

**S42. Three-state rule performance at the network level on simulation without Bonferroni correction with sample size 250.** Graphs show the proportion of studies that were positive (green, null hypothesis rejected), invalid (blue, study rejected), and negative (red, null hypothesis not rejected) with sample size 250. The proportion is plotted against confounding coefficient $c_t$ from −0.3 to 0.3, and graphs from left to right show different values for effect coefficient $c_e$ from 0 to 0.1. The nine rows represent the nine rules listed in the text.

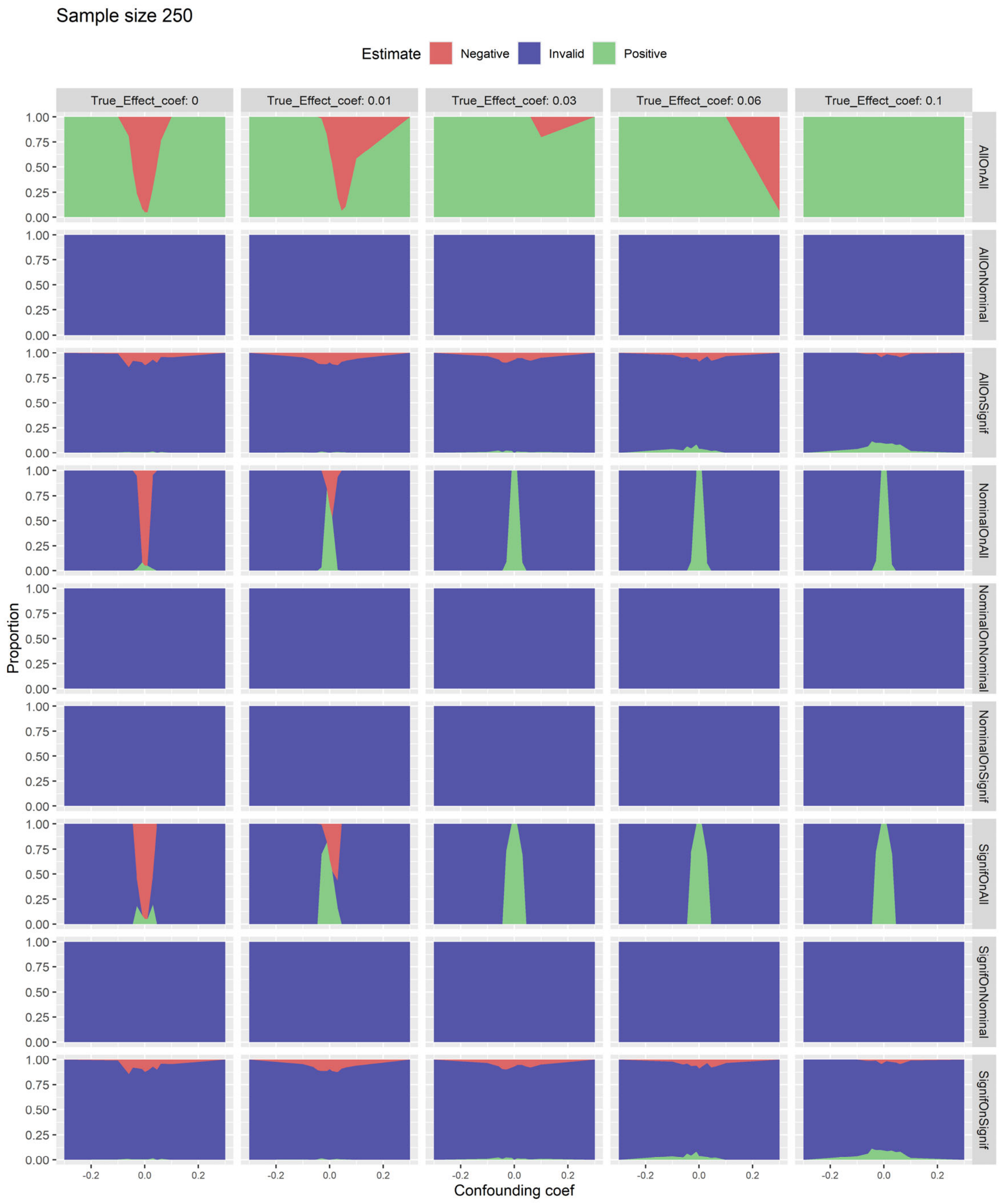

**S43. Three-state rule performance at the network level on simulation without Bonferroni correction with sample size 500.** Graphs show the proportion of studies that were positive (green, null hypothesis rejected), invalid (blue, study rejected), and negative (red, null hypothesis not rejected) with sample size 500. The proportion is plotted against confounding coefficient $c_t$ from −0.3 to 0.3, and graphs from left to right show different values for effect coefficient $c_e$ from 0 to 0.1. The nine rows represent the nine rules listed in the text.

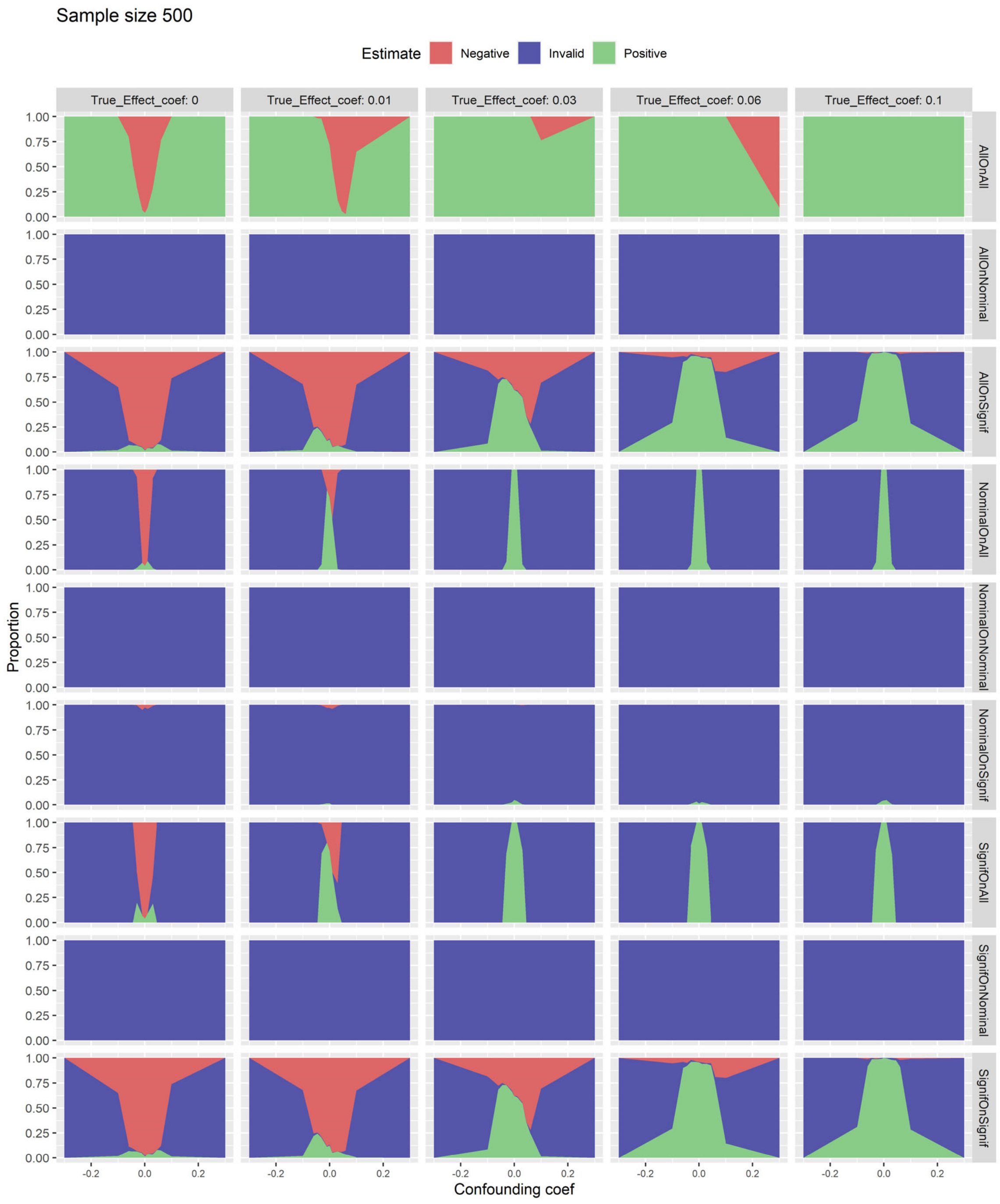

**S44. Three-state rule performance at the network level on simulation without Bonferroni correction with sample size 1000.** Graphs show the proportion of studies that were positive (green, null hypothesis rejected), invalid (blue, study rejected), and negative (red, null hypothesis not rejected) with sample size 1000. The proportion is plotted against confounding coefficient $c_t$ from $-0.3$ to 0.3, and graphs from left to right show different values for effect coefficient $c_e$ from 0 to 0.1. The nine rows represent the nine rules listed in the text.

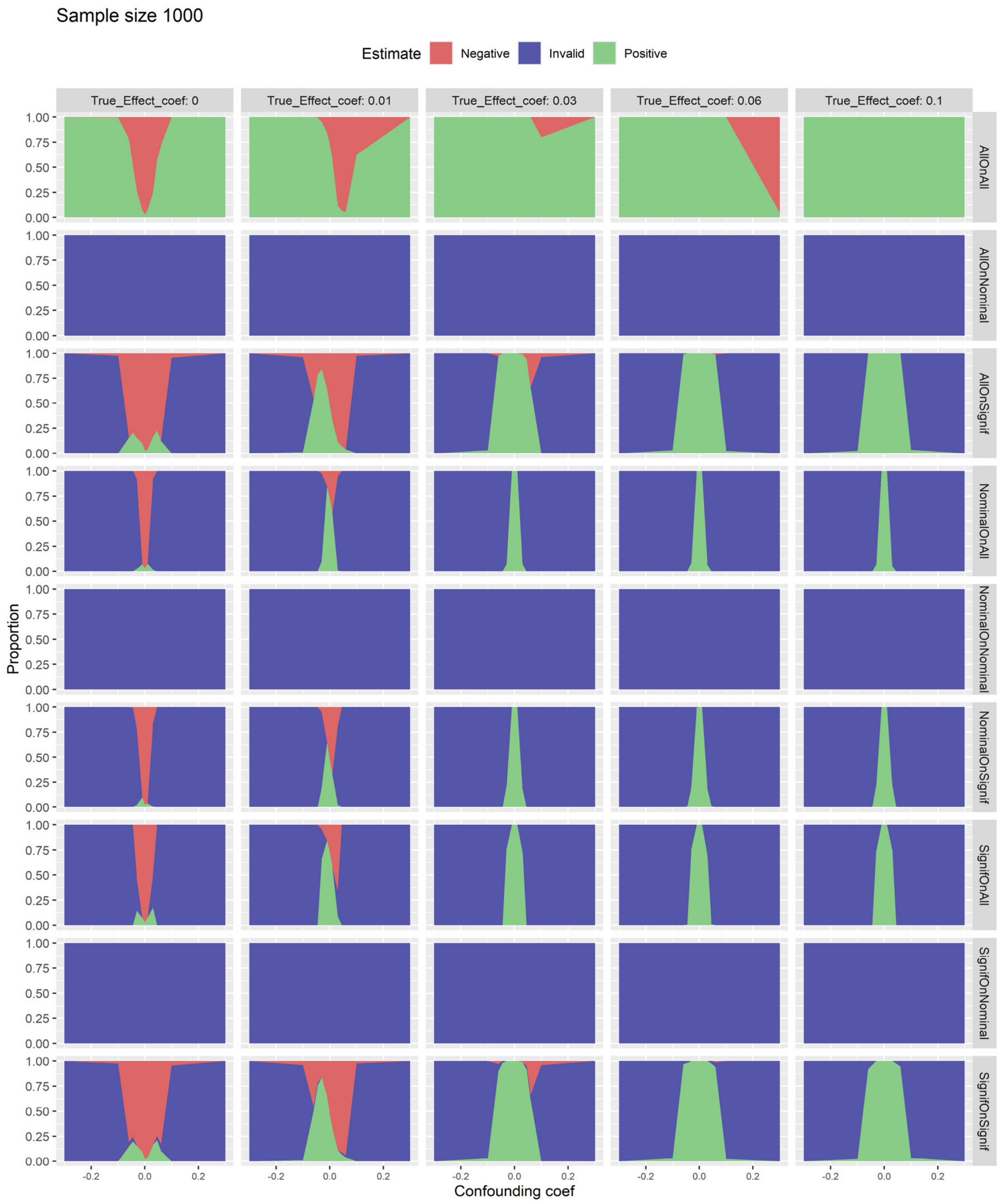

**S45. Three-state rule performance at the network level on simulation without Bonferroni correction with sample size 2000.** Graphs show the proportion of studies that were positive (green, null hypothesis rejected), invalid (blue, study rejected), and negative (red, null hypothesis not rejected) with sample size 2000. The proportion is plotted against confounding coefficient $c_t$ from −0.3 to 0.3, and graphs from left to right show different values for effect coefficient $c_e$ from 0 to 0.1. The nine rows represent the nine rules listed in the text.

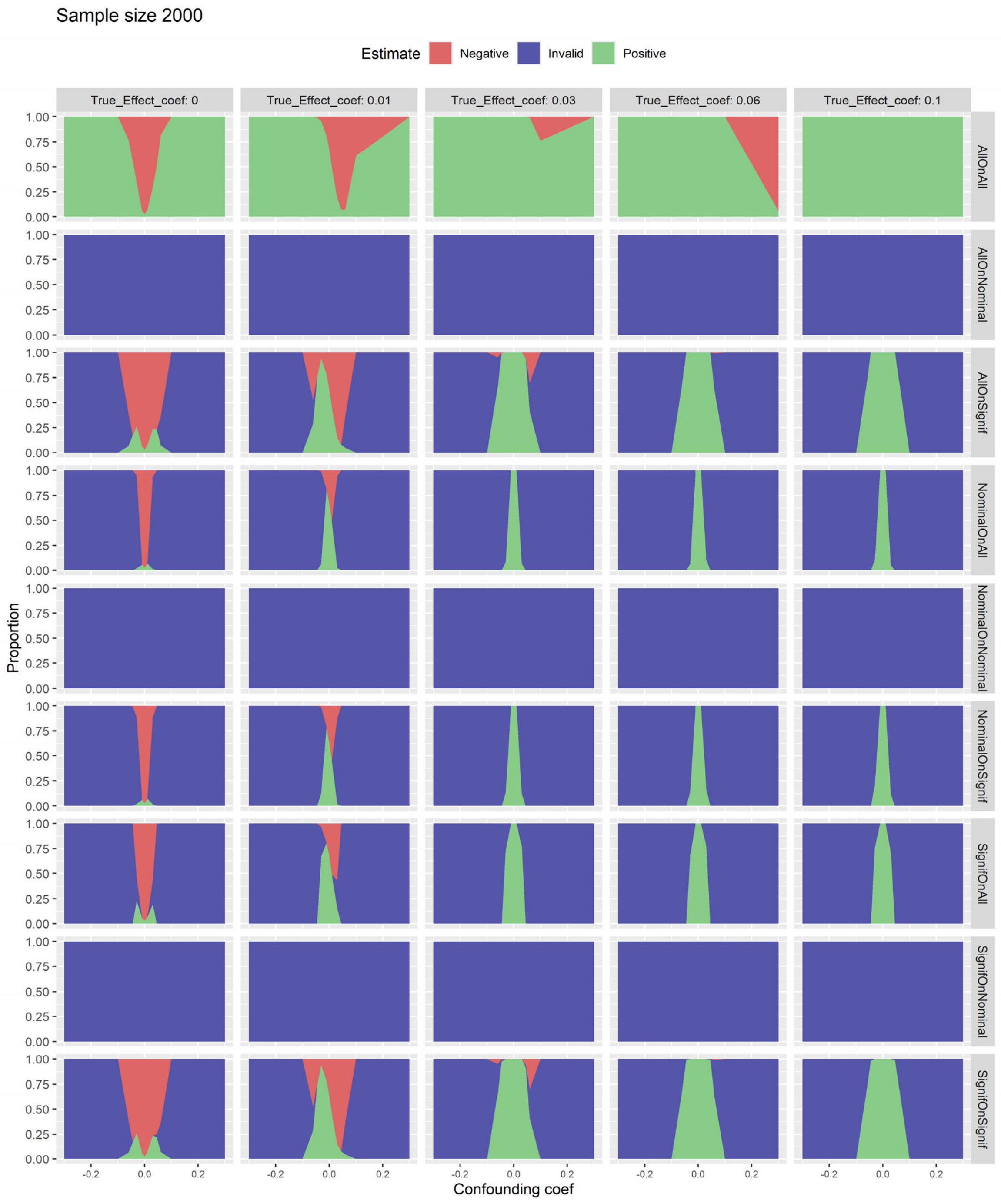

**S46. Three-state rule performance at the network level on simulation without Bonferroni correction with sample size 4000.** Graphs show the proportion of studies that were positive (green, null hypothesis rejected), invalid (blue, study rejected), and negative (red, null hypothesis not rejected) with sample size 4000. The proportion is plotted against confounding coefficient $c_t$ from $-0.3$ to 0.3, and graphs from left to right show different values for effect coefficient $c_e$ from 0 to 0.1. The nine rows represent the nine rules listed in the text.

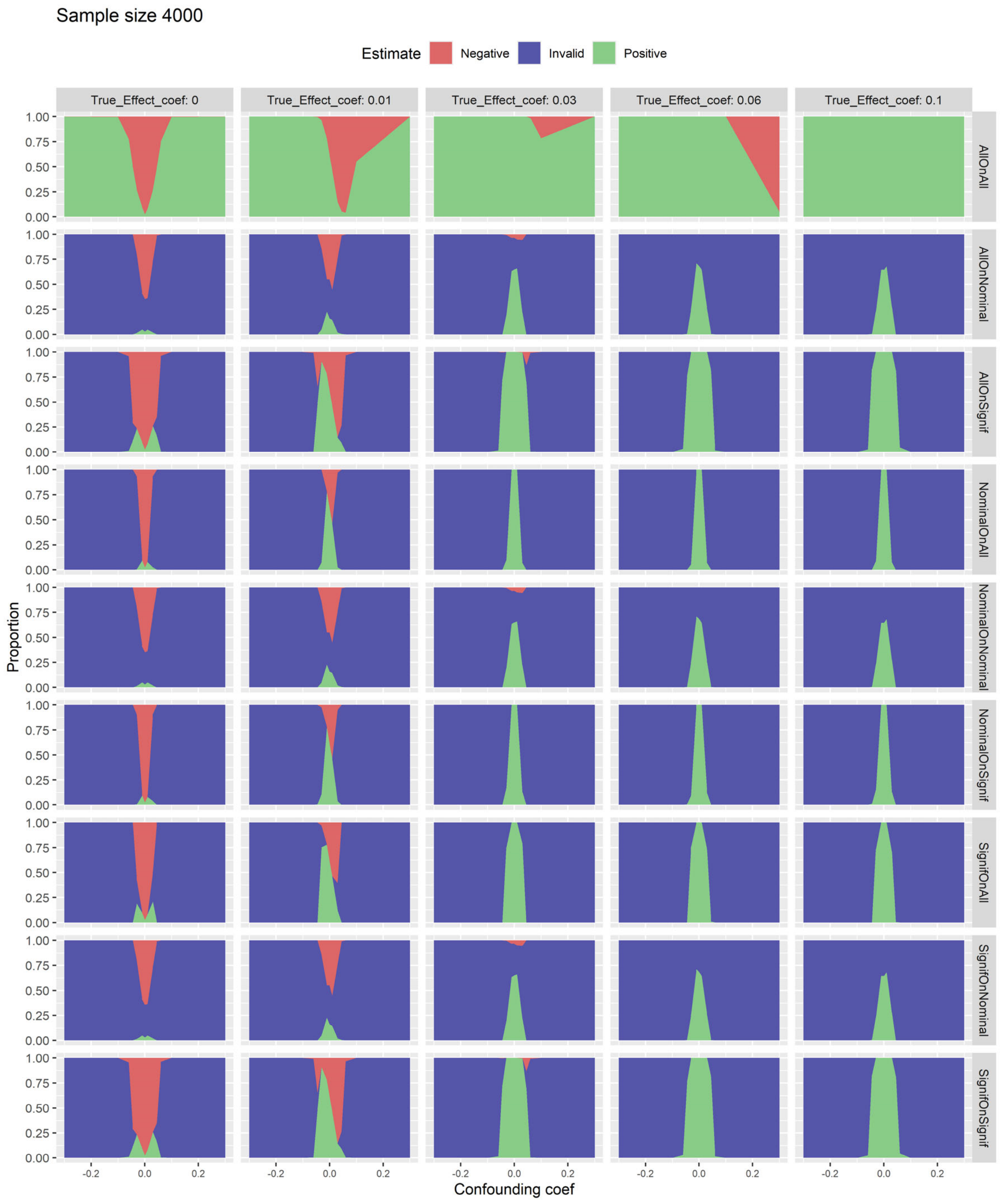

**S47. Three-state rule performance at the network level on simulation with 20 covariates with sample size 250.** Graphs show the proportion of studies that were positive (green, null hypothesis rejected), invalid (blue, study rejected), and negative (red, null hypothesis not rejected) with sample size 250. The proportion is plotted against confounding coefficient $c_t$ from −0.3 to 0.3, and graphs from left to right show different values for effect coefficient $c_e$ from 0 to 0.1. The nine rows represent the nine rules listed in the text.

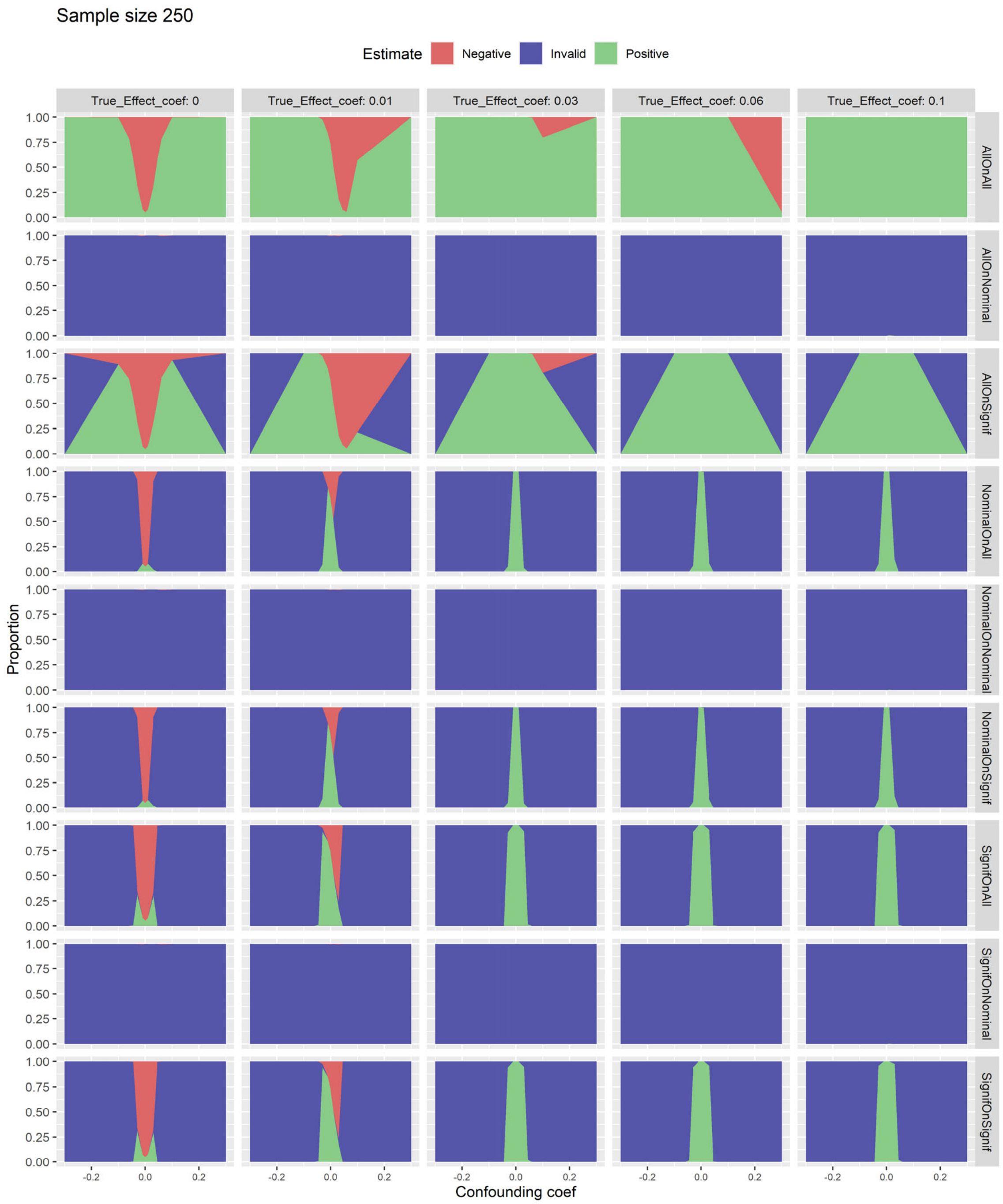

**S48. Three-state rule performance at the network level on simulation with 20 covariates with sample size 500.** Graphs show the proportion of studies that were positive (green, null hypothesis rejected), invalid (blue, study rejected), and negative (red, null hypothesis not rejected) with sample size 500. The proportion is plotted against confounding coefficient $c_t$ from −0.3 to 0.3, and graphs from left to right show different values for effect coefficient $c_e$ from 0 to 0.1. The nine rows represent the nine rules listed in the text.

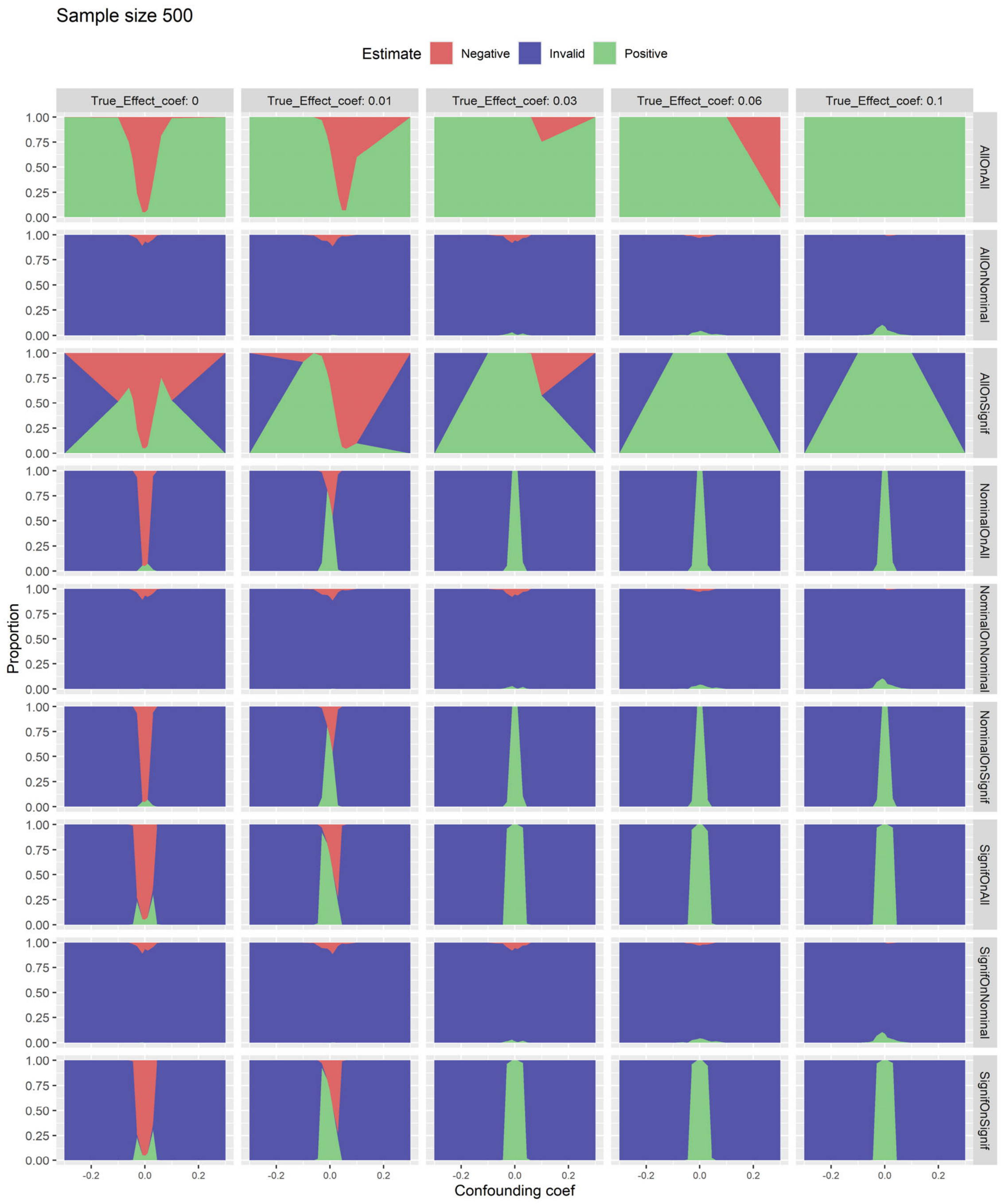

**S49. Three-state rule performance at the network level on simulation with 20 covariates with sample size 1000.** Graphs show the proportion of studies that were positive (green, null hypothesis rejected), invalid (blue, study rejected), and negative (red, null hypothesis not rejected) with sample size 1000. The proportion is plotted against confounding coefficient $c_t$ from −0.3 to 0.3, and graphs from left to right show different values for effect coefficient $c_e$ from 0 to 0.1. The nine rows represent the nine rules listed in the text.

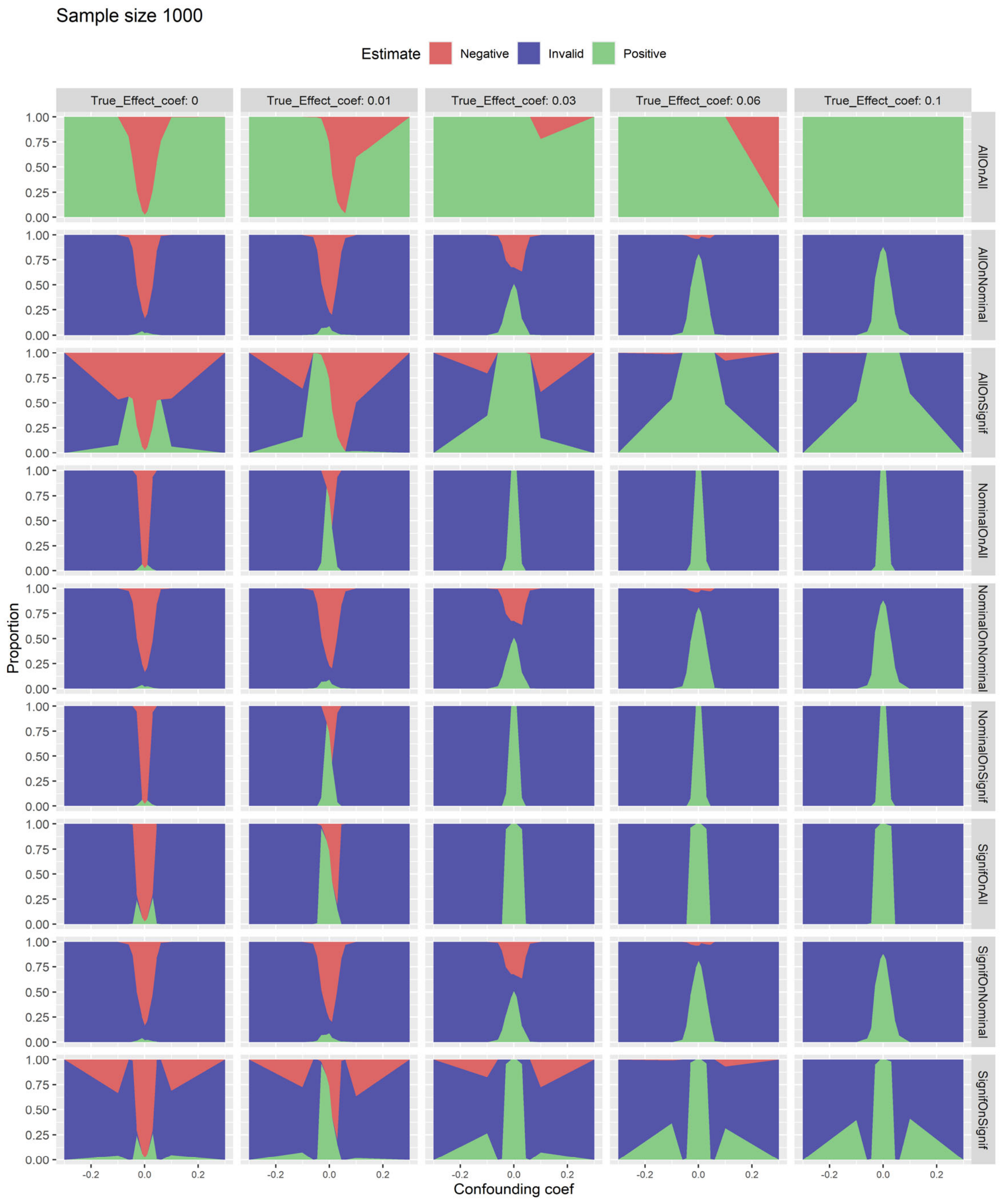

**S50. Three-state rule performance at the network level on simulation with 20 covariates with sample size 2000.** Graphs show the proportion of studies that were positive (green, null hypothesis rejected), invalid (blue, study rejected), and negative (red, null hypothesis not rejected) with sample size 2000. The proportion is plotted against confounding coefficient $c_t$ from $-0.3$ to 0.3, and graphs from left to right show different values for effect coefficient $c_e$ from 0 to 0.1. The nine rows represent the nine rules listed in the text.

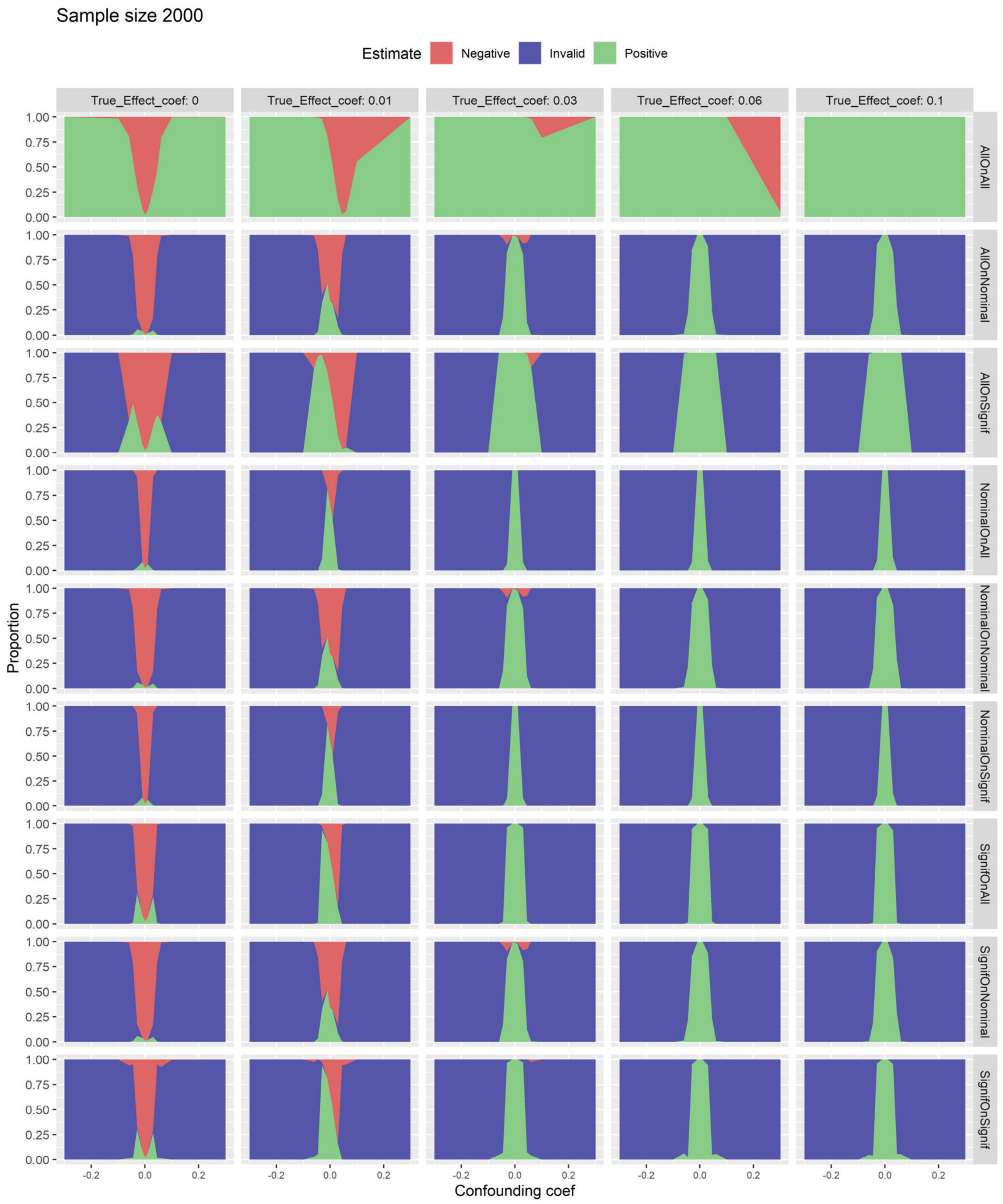

**S51. Three-state rule performance at the network level on simulation with 20 covariates with sample size 4000.** Graphs show the proportion of studies that were positive (green, null hypothesis rejected), invalid (blue, study rejected), and negative (red, null hypothesis not rejected) with sample size 4000. The proportion is plotted against confounding coefficient $c_t$ from –0.3 to 0.3, and graphs from left to right show different values for effect coefficient $c_e$ from 0 to 0.1. The nine rows represent the nine rules listed in the text.

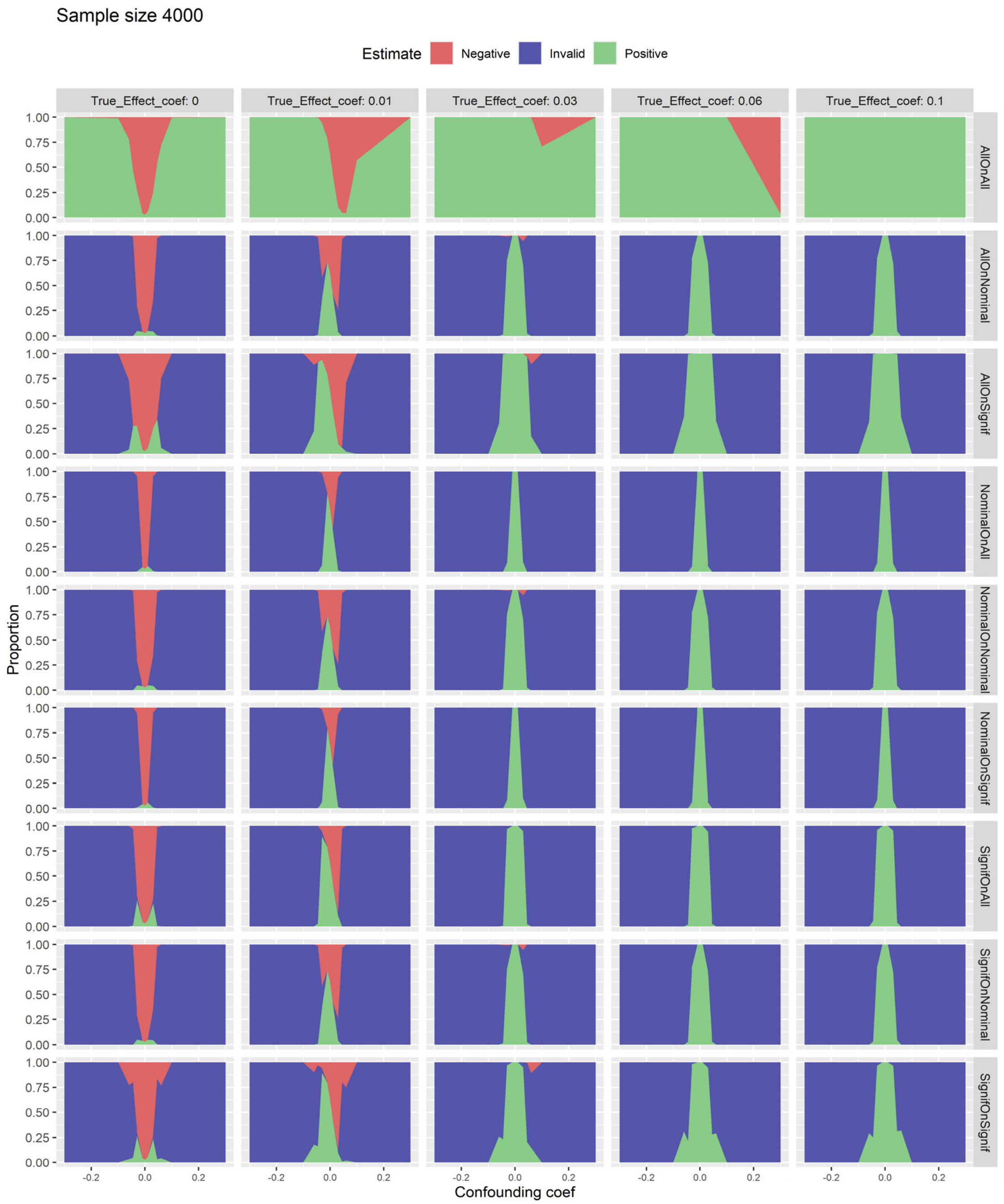
